# DNA methylation fingerprint for the diagnosis and monitoring of hepatocellular carcinoma from tissue and liquid biopsies

**DOI:** 10.1101/2021.06.01.21258144

**Authors:** Emanuel Gonçalves, Maria Reis, José B Pereira-Leal, Joana Cardoso

**Affiliations:** Ophiomics, Pólo Tecnológico de 8, R. Cupertino de Miranda 9, 1600-513 Lisboa, Portugal

**Keywords:** HCC, DNA Methylation, Liquid Biopsies, Early Detection, Monitoring

## Abstract

**Background:** Hepatocellular carcinoma (HCC) is amongst the cancers with highest mortality rates and is the most common malignancy of the liver. Early detection is vital to provide the best treatment possible and liquid biopsies combined with analysis of circulating tumour DNA methylation show great promise as a non-invasive approach for early cancer diagnosis and monitoring with low false negative rates.

**Methods:** To identify reliable diagnostic biomarkers of early HCC, we performed a systematic analysis of multiple hepatocellular studies and datasets comprising >1,500 genome-wide DNA methylation arrays, to define a methylation signature predictive of HCC in both tissue and cell-free DNA liquid biopsy samples.

**Results:** Our machine learning pipeline identified differentially methylated regions in HCC, some associated with transcriptional repression of genes related with cancer progression, that benchmarked positively against independent methylation signatures. Combining our signature of 38 DNA methylation regions, we derived a HCC detection score which confirmed the utility of our approach by identifying in an independent dataset 96% of HCC tissue samples with a precision of 98%, and most importantly successfully separated cfDNA of tumour samples from healthy controls. Notably, our risk score could identify cell-free DNA samples from patients with other tumours, including colorectal cancer.

**Conclusions:** Taken together, we propose a comprehensive HCC DNA methylation fingerprint and an associated risk score for the early diagnosis and early relapse detection of HCC from liquid biopsies.

## Introduction

Liver cancer is one of the deadliest types of cancer, with a 5-year overall survival rate lower than 20% and death rates increasing around 1.7% each year [1,2]. Hepatocellular carcinoma (HCC) is the most common malignancy of the liver accounting for nearly 90% of all cases [1,3–5]. Major risks of HCC include cirrhosis, viral infection with hepatitis B virus (HBV) or hepatitis C virus (HCV), alcoholic liver, non-alcoholic fatty liver disease and inherited traits such as metabolic diseases [1,6]. Current HCC diagnostic guidelines report the usage of invasive procedures, such as tissue biopsies, followed by histological and/or contrast-enhanced imaging [7]. This contributes to HCC being often detected in an advanced stage where it is estimated that 40% of the cases are multinodular or expanded beyond the liver leaving patients with limited therapeutic options [5,8]. Screening, surveillance and monitoring programmes are therefore vital to diagnose and detect HCC as early as possible to provide patients with the best treatment possible [9–11]. In addition, HCC patients surgically treated often experience relapses and early detection could bring better management of the disease and increase patient’s life quality and span [12].

Body fluids, for example plasma, serum and urine, contain circulating biomarkers that can be measured non-invasively and inexpensively for diagnosis and monitoring of HCC [5,13,14]. Among others, alpha-fetoprotein (AFP) is often proposed as a diagnostic biomarker present in serum or plasma of high-risk individuals for HCC [7,13,15], nonetheless official guidelines indicate that AFP has no diagnostic approved role [3,4]. High levels of AFP are considered diagnostic of HCC with almost perfect specificity, although sensitivity (recall) rates are frequently low, less than 45% [7]. Lower thresholds of AFP (20 ng/ml) comprises a balance between specificity and sensitivity with both ranging around 79% [7]. Of note, in patients with chronic liver disease, the population where screening methods are most needed, the precision of AFP is significantly reduced and insufficient for robust diagnosis [7,16,17]. This is particularly problematic since chronic liver diseases are the major risk factor for HCC, thus novel non-invasive and accurate clinical approaches are needed to improve cancer detection.

Liquid Biopsies (LBs) have recently emerged as a promising approach for early detection of tumours by characterising circulating tumour cells or circulating tumour free nucleic acids [18]. LBs contain cell-free DNA (cfDNA) material evocative of cells from the entire body, including varying levels of circulating tumour DNA (ctDNA) [19,20] that is estimated to range between 0.1% and 10% in cancer patients [21–23]. Measurement of genetic markers in ctDNA, such as mutations and methylation, can be used as a diagnostic and therapeutic tool [13,18,20,24–32] and provide complementary information to tissue samples, for example circumventing potential tissue heterogeneity which might result in sampling bias [33,34].

DNA methylation plays an important role in cancer initiation and progression through the repression of tumour suppressor genes by promoter hypermethylation and promoter hypomethylation of many oncogenes [35–38]. Importantly, DNA methylation changes characteristic of cancer cell formation are often observed in early stages of carcinogenesis [39–42]. Hence, ctDNA methylation holds great promise for early cancer detection and monitoring, with systematic studies showing it outperforms other genetic markers like mutations and copy number alterations [26,43]. For example, promoter methylation of the gene Septin 9 (SEPT9/ SEPTIN9) is a plasma derived biomarker for colorectal cancer and is being studied for HCC [27,44]. Several studies have focused on the identification of DNA methylation biomarkers for HCC [43,45–48], nonetheless these were limited to either tissue samples only, focused on the identification of small sets of single CpG sites, and/or mostly compared to healthy liver tissue samples. Relying on the accurate measurement of very specific and small sets of methylation biomarkers mostly derived tissue samples may hinder the clinical generalisation of these methylation signatures to LBs and other cohorts. Additionally, it is fundamental to ensure that signatures can distinguish HCC patients from a background of chronic liver diseases, where current non-invasive molecular markers perform worse [7,16,17].

Here, we perform a systematic discovery of a HCC methylation signature by compiling 1,551 genome-wide DNA methylation arrays from 13 studies [1,31,45,46,49–58], including both tissue and liquid biopsy samples from HCC, cirrhosis and healthy controls. We developed a machine learning pipeline to harness this resource to identify differentially methylated regions (DMRs) predictive of HCC in both tissue and liquid biopsies, from a background of cirrhotic samples. Our approach benchmarked favourably against 12 independent HCC methylation signatures and supported the development of a novel signature comprising 38 DMRs. Some of the identified regions were associated with transcriptional repression of several members of the Zinc Finger Proteins (ZFNs) family suggesting a potential role with cancer progression and early onset. Lastly, we combined the information of the novel DMR signature into a single score which successfully identified HCC tissue samples in an independent dataset (recall 96% and precision 98%), perfectly classified 13 healthy cfDNA samples, and identified 7 (out of 11) tumour cfDNA samples. Of note, the DMR signature score successfully identified cfDNA from diverse tumours, including colorectal and breast cancer, showing its potential as a diagnostic tool for multiple cancers. Overall, we present a systematic discovery and benchmark of methylation biomarkers for the early detection and monitoring of HCC using tissue and liquid biopsies and propose an improved signature and risk score with the potential to be used for non-invasive clinical diagnostics.

## Results

### DNA methylation dataset for the discovery of HCC biomarkers

To systematically discover DNA methylation biomarkers for the detection of HCC from tissue and plasma cfDNA samples we performed a comprehensive search of HCC-related studies and datasets characterising genome-wide DNA methylation changes (Figure 1a). We queried commonly used data repositories, GEO [59,60] and ArrayExpress [61], using the keywords Hepatocellular Carcinoma, cfDNA and ctDNA. To ensure an exhaustive analysis of methylation markers we focused on studies that provided high-throughput assays and specifically Illumina-based, Infinium 450K and EPIC assays, as these have been broadly adopted by large-scale studies. Additionally, to minimise potential undesired and technical batch effects while integrating multiple data sources for model training, only studies that provided raw unprocessed files were considered to allow the same processing pipeline to be applied to all samples [62–64]. Matching the criteria defined above we assembled 859 samples from 6 different studies [31,45,46,56–58] covering: HCC and cirrhotic samples from tissue and cfDNA, including cirrhotic tissue from multiple aetiologies; healthy controls from both liver tissue and cfDNA; other non-HCC diseased tissue (e.g. liver obesity and Alpha 1 antitrypsin deficiency); and cfDNA from non-HCC patients (e.g. sepsis and other cancer types) (Figure 1a and 1b and Supplementary Figure 1a and 1b). A total of 452,567 methylation sites (CpG sites) are measured and methylation levels represented using beta methylation values, ranging between 0, hypomethylated, and 1, hypermethylated. Additionally, we compiled a Validation dataset containing 692 tissue samples from 7 independent datasets [1,49–55] for which original data or publication was not accessible but processed beta methylation values was available (Figure 1a, Supplementary Figure 1c). This validation dataset comprises multiple studies with distinct experimental and analytical pipelines and is intended to be used as independent validation of the approaches adopted in this study. Overall, we assembled 1,551 whole-genome DNA methylation samples (Supplementary Table 1) representing an heterogenous and comprehensive resource to discover and benchmark DNA methylation biomarkers of HCC (Supplementary Figure 1d and 1e) from clinically relevant diseased backgrounds, such as cirrhosis.

**Figure 1.**
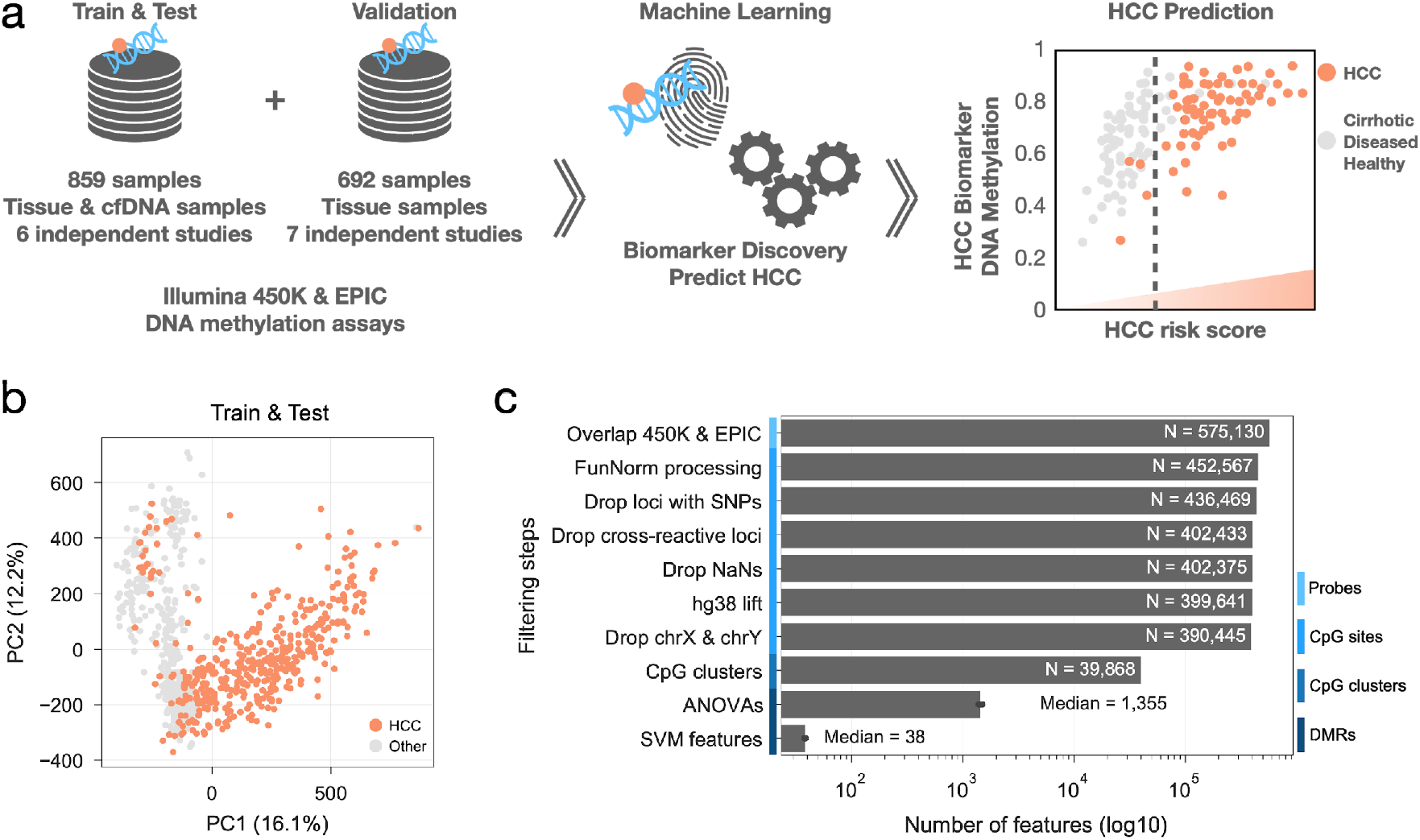
Data and workflow overview. a) diagram depicting the different datasets assembled to discover Hepatocellular Carcinoma (HCC) DNA methylation biomarkers using machine learning approaches and to construct a HCC risk score. b) principal component analysis (PCA) of the Train & Test DNA methylation dataset highlighting HCC samples. Principal component explained variances is shown within brackets. c) Feature, i.e. probes, CpG sites, CpG clusters and differential methylated regions (DMRs), reduction steps across different stages in the processing and feature discovery pipeline.

### Selection of high quality and informative DNA methylation regions

The assembled dataset measures >450,000 CpG sites which is several orders of magnitude greater than the number of samples, thus posing a number of problems for training informative models. To mitigate this and to focus on high quality and informative measurements we applied several filtering steps to reduce the number of CpG sites in 14% to 390,445 (Figure 1c). Secondly, while a single CpG site can be informative and have strong predictive power of HCC status, due to the much larger number of CpG sites compared to the number of samples this can lead to spurious associations that are unlikely to be functionally relevant and generalisable to other cohorts, i.e. overfit. Considering that HCC patient samples showed distinct patterns of multiple and clustered CpG sites with hypo and hyper methylation profiles [58], we searched for CpG clusters, spanning at least 3 CpG sites, such that two consecutive sites are at most 500 base-pairs (bp) apart. This defined a total of 39,868 CpG clusters with a median size of 700bp spanning all 22 autosomal chromosomes (Figure 1c, Supplementary Figure 2a). For each CpG cluster we took the mean methylation of all CpG sites contained in it. Taken together, we performed an unsupervised reduction of the number of features by excluding problematic CpG sites and to focus on genomic regions, instead of individual CpG sites, to reduce the impact of potential confounder effects and help discover more generalisable biomarkers of HCC.

### Discovery of methylation regions predictive of HCC

To identify HCC from a background of cirrhotic samples in tissue and cfDNA we set out to find methylation regions predictive of HCC by training linear support vector machine classifiers (LinearSVC) (Figure 2a). We applied a leave-one-out cross-validation strategy, where one sample at a time was left out for testing the prediction, while the other 858 samples were used as a training set. Considering there are many more tissue samples compared to cfDNA, this can create potential biases when training the LinearSVC (e.g. classes with more samples will weigh more on the importance of the features). To address this we balanced the number of samples of each class by randomly under-sampling the tissue samples to obtain 22 HCC (HCC-T) and 22 cirrhosis (C-T) samples, complemented with 22 HCC cfDNA (HCC-CF) and 22 cirrhosis cfDNA samples (C-CF). One balanced dataset per leave-one-out fold is generated ensuring that the sample left out for testing is not considered.

**Figure 2.**
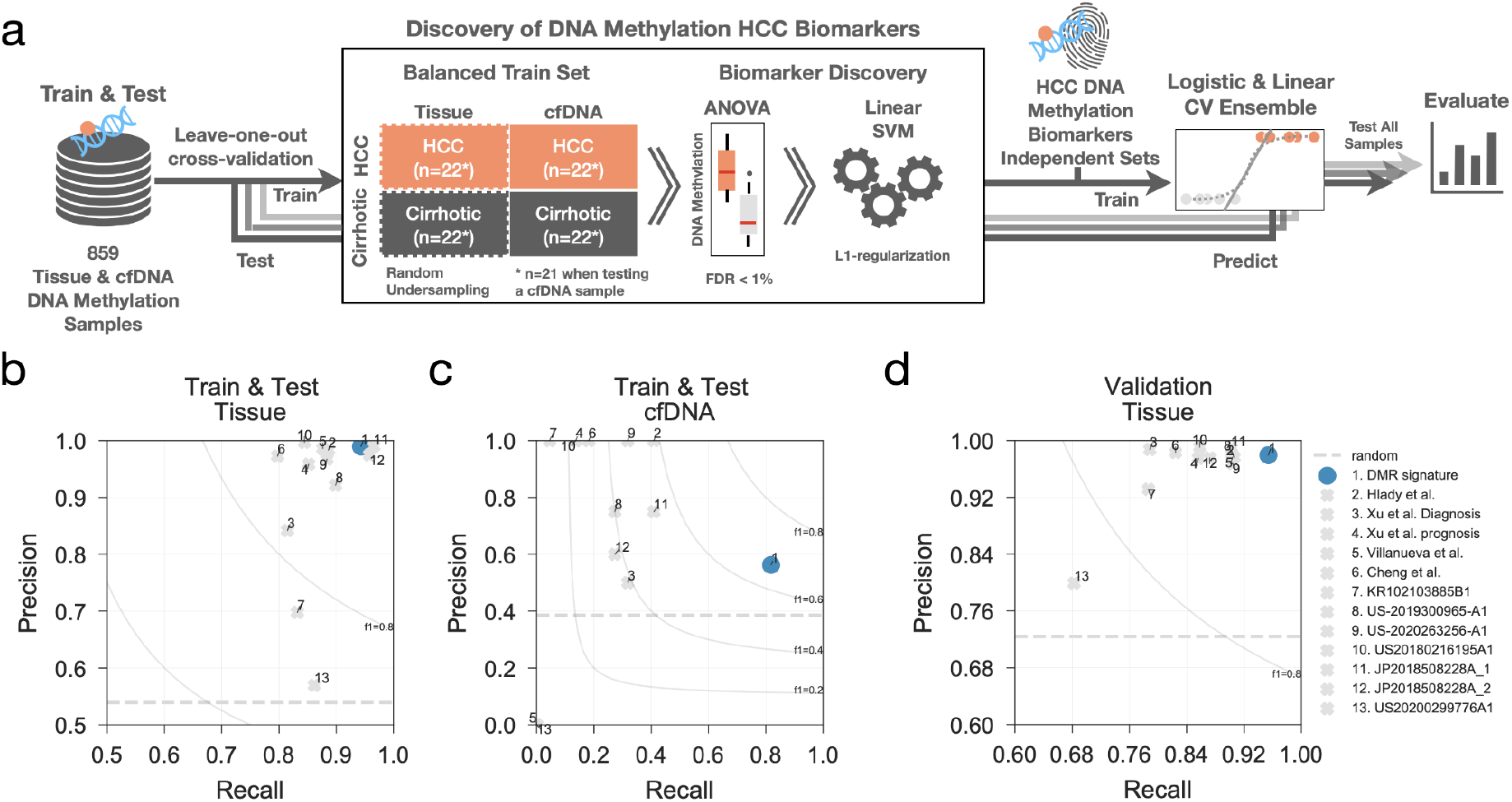
HCC biomarker discovery and benchmark pipeline. a) machine learning workflow to identify DNA methylation regions predictive of HCC samples using balanced training sets and support vector machines and then benchmark against other independent DNA methylation biomarkers using an ensemble of logistic and linear regression classifiers. b) precision and recall rates calculated over the leave-one-out test samples predicted using the logistic and ridge regression classifier ensemble. Similarly, precision and recall rates are calculated using the same ensemble but trained with CpG sites from independent HCC DNA methylation biomarkers and are compared. Here, only tissue samples of the Train & Test dataset are considered for the calculation of the precision and recall metrics. c) similar to b), instead precision and recall for cfDNA samples only are reported. d) precision and recall rates obtained predicting the independent Validation set using the same ensemble trained with the multiple HCC biomarker feature sets measured in the Train & Test dataset.

Differentially methylated and predictive regions are discovered using the balanced datasets in a two-step approach. Firstly, differentially methylated regions (DMR) are identified by removing potential cofounder effects, i.e. sex, age, global methylation and tumour purity. Considering that sex and age were not available for all samples, we estimated them from the DNA methylation arrays [62,65,66]. Global changes in methylation affect large swaths of CpG sites and thereby these do not represent optimal candidates for biomarkers due to their lack of specificity (Supplementary Figure 2b and 2c). Lastly, the varying tumour purity of TCGA samples, the biggest source of HCC tissue samples in our analysis, has been quantified and represents a technical limitation that can affect molecular measurements, including DNA methylation, and their interpretation [67]. Tumour purity estimation is only available for TCGA samples. We observed from the PCA analysis of Train & Test dataset that PC5 is significantly correlated with tumour purity (Pearson’s r=0.6, p-value=3.28e-37). Therefore we considered PC5 as a proxy of tumour purity impact in the DNA methylation measurements (Supplementary Figure 2d and 2e). A differential methylation analysis between HCC (HCC-T and HCC-CF) and cirrhotic (C-T and C-CF) samples was performed taking the previous variables as covariates in the linear model in order to discount their potential impact. Only significantly differentially methylated CpG clusters (likelihood-ratio test FDR < 1%) were selected for model training, thus reducing the number of features to a median of 1,355 DMRs, across all leave-one-out folds (Figure 1c). Secondly, DMRs are then used to train LinearSVC models for each cross-validation fold using a L1-regularization parameter to further reduce the number of DMRs to find the top predictive biomarkers of HCC. A median of 38 DMRs were selected per model (Figure 1c). Taken together, this identified 150 DMRs that are present in at least 5% (n=43) of all trained models (Supplementary Table 2) and the frequency of the DMRs in the optimal LinearSVC across the leave-one-out cross-validation is positively associated with their absolute mean effect size (Spearman rho=0.29 and p-value=1.9e-41, Supplementary Figure 2f).

In conclusion, the feature selection and model training steps performed in each cross-validated train set avoids information leak between train and test sets, addresses the problem of having many more features than samples and identifies the most predictive DNA methylation biomarkers of HCC.

### Evaluation, comparison and assembly of HCC methylation signature

Next, we set to define a DNA methylation signature predictive of HCC and compare it against independently defined sets. We estimated the optimal number of DMRs to consider in the methylation signature by sequentially testing the addition of DMRs into the feature set and tested the increment in precision and recall of the LinearSVC models (Supplementary Figure 3a). Recall and precision shows the steepest increase up to 10 DMRs, and from that point the test and validation datasets show small but consistent increments in performance. Together with the fact that frequency of each DMRs in the optimal models is positively correlated with its absolute mean effect size, we selected the top 38 most frequent DMRs in the leave-one-out cross-validation procedure (Supplementary Table 3). The selected DMRs encompass hyper and hypo methylation events in HCC that are largely consistent across both Train & Test and Validation datasets and unsupervised clustering separates most HCC from non-HCC samples (Supplementary Figure 3b).

We then benchmarked our DNA methylation signature against other similar approaches, assembling from the literature 12 sets of CpG sites proposed in 4 publications [1,31,47,68] and 7 patents [69–75]. Notably, the DNA methylation sets were largely non-overlapping (Supplementary Figure 4a) suggesting a disparity among HCC biomarkers and possibility indicating datasets-specific features which might not generalise well to other patient cohorts. To avoid potential methodological bias, we used an ensemble model using logistic and linear classification models, different from the support vector machine model we used previously to identify informative regions, and iteratively predicted the HCC status of the sample that is left-out for testing in a leave-one-out cross-validation (Figure 2a). The performance of all models was estimated using multiple metrics, i.e. recall, precision, accuracy, Mathew’s correlation coefficient (MCC) and balanced accuracy (Supplementary Figure 4b, 4c and 4d). It is important to note that most of the feature sets were derived using part of the DNA methylation datasets also utilised in this study, thus a complete independent validation of these feature sets was not possible, and it is expected that metrics will be overestimated. Overall precision and recall scores across the tissue are greater than 80% (Figure 2b) and all models had a poorer performance when predicting the subset of cfDNA samples, while precisions were less affected (Figure 2b and 2c). We then used the Validation tissue samples dataset as an independent benchmark, and observed that overall feature sets provided a mean precision of 96% and recall rates of 86% (Figure 2d and Supplementary Figure 5), where our signature obtained the highest recall (95%) while preserving precision (98%) (Figure 2d).

Collectively, our approach identifies a signature of hyper and hypo methylated regions that successfully distinguishes HCC samples from cirrhotic, healthy and other non-HCC samples, and benchmarks positively against other DNA methylation signatures, particularly showing low false negative rates, i.e. high recall, both in tissue and cfDNA samples.

### Molecular characterisation of methylation biomarkers

Having assembled a methylation signature of HCC, we then set out to molecular characterise it in more detail. The top 38 DMRs encompasses a total of 214 CpG sites out of which 118 and 74 showed significant hyper and hypo methylation in HCC, respectively (Figure 3a, Supplementary Table 3). Reassuringly, inspecting the top DMRs showed that the methylation of the CpG sites within each cluster is able to clearly separate between HCC and non-HCC samples in both tissue and cfDNA samples (Figure 3b and 3c). We further explored this by taking advantage of the availability of gene-expression datasets for 410 liver samples from the TCGA consortium [56,76], and systematically tested associations between the 38 DMRs and 15,341 gene expression profiles. We identified a total of 39 significant DMR-gene associations (linear regression log-likelihood ratio test FDR < 10%, Supplementary Table 4). Among the top associations are several positive associations between DMR Chr7:27144326-27145664 and multiple members of the homeobox transcription factors (HOXA6, HOXA3, HOXA5, HOXA7 and HOXA4) (Supplementary Figure 6a) which are all genomically close to the DMR and have been suggested to be involved in tumorigenesis and cell proliferation and migration [77,78]. While positive associations, i.e. increase in methylation associated with increased gene expression, might be related with potentially more complex regulatory mechanisms, negative associations might capture decreased gene expression through repression of transcription due to hypermethylation. We observed multiple negative associations with Zinc Finger Proteins (ZNF518B, ZNF502 and ZNF132) (Supplementary Figure 6b). The role of the Zinc Finger Proteins in cell adhesion and in cancer is well described [79,80] and could highlight some of the biological mechanisms underlying hypermethylation of these regions in HCC (Supplementary Figure 6c). In summary, the methylated DNA regions highlighted with our approach, are potential useful biomarkers for HCC and may also reveal important biological information, specifically ZNF518B and its associated DMR, Chr10:133445694-133446718, is among the most important features and has been previously described with possible implications in cancer cell invasion and metastatic potential [81].

**Figure 3.**
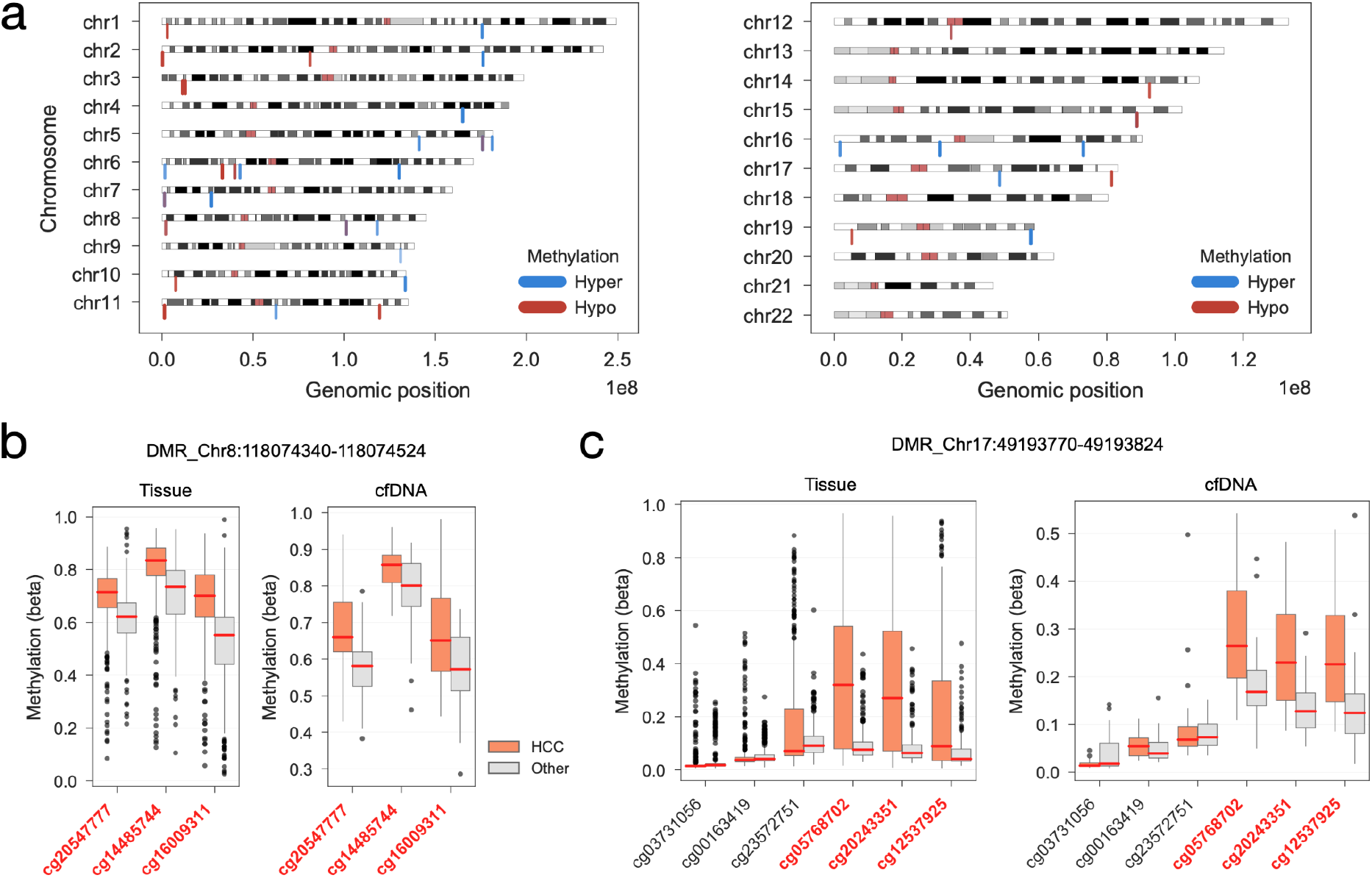
HCC DNA methylation biomarkers. a) genomic localisation of the significantly differentially methylated CpG sites contained in the top 38 DMRs. Blue represents hypermethylation and red hypomethylation in HCC. b) Top recurrent DMR in the optimal LinearSVC models. Distribution of DNA methylation (beta) of CpG sites contained within 1,000 base-pairs up/down-stream of the DMR. In red are labeled CpG sites that are contained in the DMR. DNA methylation is split and coloured by HCC and the rest. Left panel shows the methylation of all tissue samples in the Train & Test dataset, and right-hand side the DNA methylation of cfDNA HCC, cirrhotic and healthy samples. Above the plots are reported the DMR associated chromosome and genomic coordinates. c) similar to b), instead showing the distribution of a representative DMR that is highly predictive of HCC in both tissue and cfDNA samples.

### Diagnostic score based on HCC methylation signature

Lastly, we defined a single metric that could encompass the information from a whole DNA methylation signature to use as a diagnostic metric for early detection of HCC. First, we robustly estimated the coefficients of each DMR in the signature by randomly generating 1,000 balanced training datasets, as described before (Figure 2a), and training a regularised linear regression classifier (Supplementary Figure 7a). Sorted in descending order of their absolute coefficients, the top 8-10 DMRs in the signature contribute most to the recall of HCC in the Validation dataset using the Train and Test dataset for training, while the remaining DMRs provide smaller but consistent improvement (Supplementary Figure 7b). Secondly, we built an additive linear score (DMR signature score) where each 38 DMRs of the methylation signature is weighted by their signed mean coefficients, i.e. DMRs with high absolute mean coefficients have higher preponderance in the score. For all samples in the Test and Train and Validation dataset we calculated their DMR signature score and ranked them into how probable they are from being HCC (Supplementary Table 5). Similarly, we estimated a linear risk score for the other CpG site signatures, and observed that in the independent Validation dataset the score based on our DMRs signature outperformed and provided very accurate predictions of HCC (Supplementary Figure 7c). Furthermore, in samples from the Train & Test dataset that were held out from the training of the DMR signature and score could achieve a clear split between the HCC compared to non-HCC samples with a recall (sensitivity) of 86% and precision of 83% (Figure 4a and 4b).

**Figure 4.**
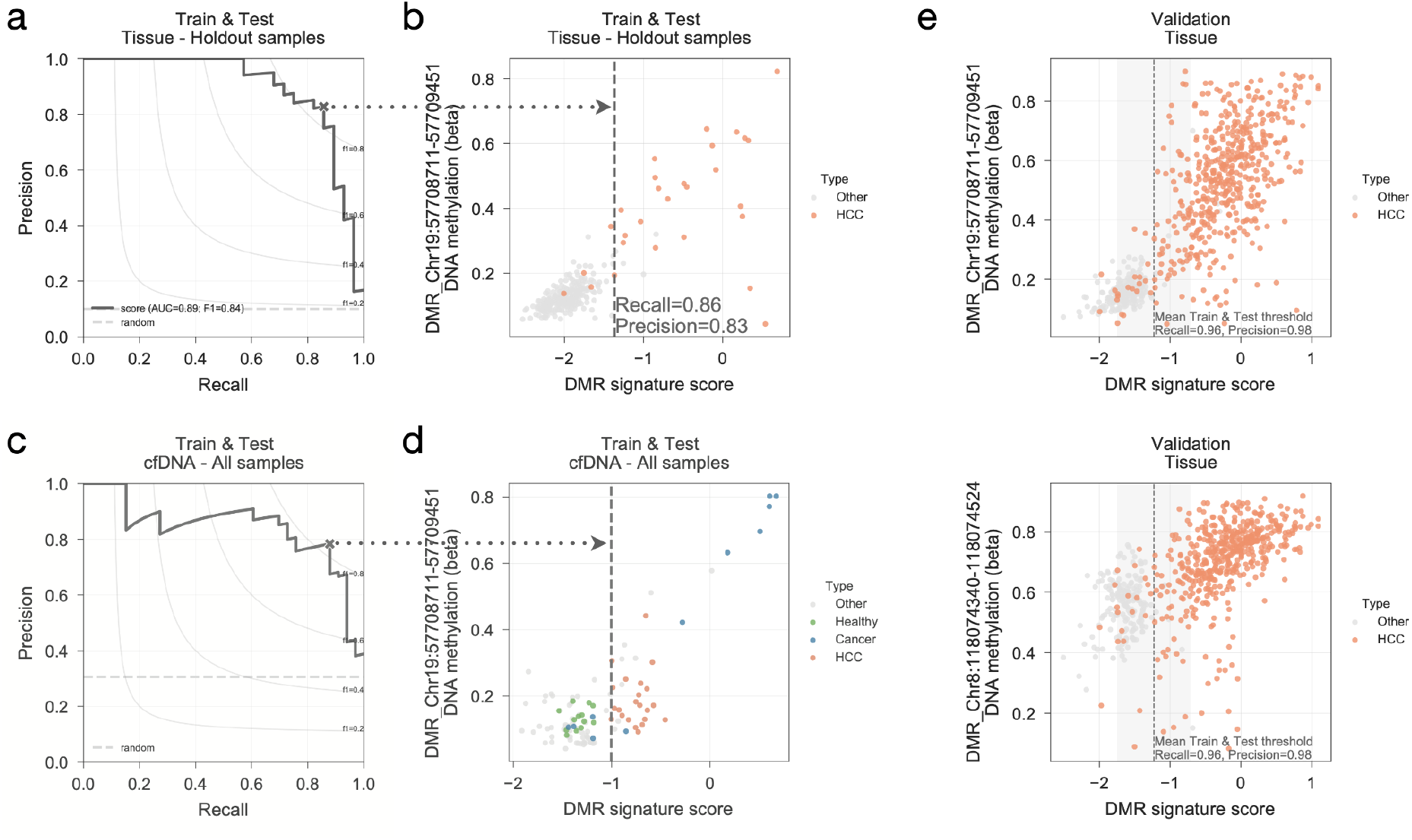
HCC DMR signature score. a) precision-recall curve of the DMR signature linear risk score ranking exclusively samples in the Train & Test dataset that were not used to identify and estimate the HCC biomarkers and weights. Maximum F1-score along the curve is represented with “x” and used to define the DMR signature score threshold at the given recall and precision. Random precision is drawn as a dashed horizontal line. b) DMR signature score of Train & Test samples not used for HCC biomarker discovery plotted against a representative top performing DMR. Vertical line represents the DMR signature score threshold found at the maximum F1-score in a) and the associated recall and precision rates are reported. c) precision-recall curve of all cfDNA samples of the Train & Test dataset including samples from patients with other types of cancer (labeled as “Cancer” and coloured blue). d) similar to b), DMR signature score threshold, vertical dashed line, is estimated from the maximum F1-score point along the precision-recall curve in c). e) DMR signature score calculated for the Validation set samples plotted against two highly predictive HCC DMRs and their methylation profiles. Precision and recall rates reported are those estimated in the Validation dataset using the DMR signature score threshold calculated with the Train & Test dataset.

We also looked in particular to the cfDNA samples which have noisier backgrounds in terms of methylation signals and are more relevant for non-invasive early-stage diagnostic approaches based on blood liquid biopsies. In addition to the HCC and cirrhotic cfDNA samples, we also considered cfDNA samples of healthy controls, sepsis and patients with cancers from other tissues, including lung, breast and colon [57]. Not surprisingly our metric could separate cfDNA HCC and cirrhotic samples, which are used for training of the signature and score. More interestingly, it perfectly splitted independent healthy control samples and could identify cfDNA samples from patients with other cancers (Figure 3c and 3d), supporting the capacity of our signature and associated score.

Altogether, the linear risk score represents a metric for the diagnosis of HCC that showed robust predictive power across many different datasets (Figure 4e) with heterogeneous backgrounds and most importantly both in tissue and liquid biopsies (Supplementary Figure 7d and 7e). While the recall and precision metrics reported here are limited to the amount of cfDNA datasets available these results suggest that DNA methylation from plasma cfDNA is a promising alternative to AFP-based approaches.

## Discussion

Hepatocellular carcinoma (HCC) diagnosis is challenging and often misses early detection which is vital to ensure curative options are available to the patient. Non-invasive diagnostic approaches based on serum biomarkers, such as alpha-fetoprotein (AFP), AFP isoforms and micro-RNAs, have shown sub-optimal sensitivity, leaving many patients undiagnosed. Tumour cell-free DNA (cfDNA) from blood liquid biopsies holds great promise to transform clinical oncology diagnosis [26,43,82,83] with several studies reporting highly specific methylation signatures for the diagnosis and prognosis of HCC [1,31,47,68]. Currently, most HCC methylation signatures are small sets of single CpG sites (median n = 7) and overall show poor agreement between them. This might indicate these signatures are potentially specific to the studies, which could hinder generalisation to other cohorts and the utility for liquid biopsies as these are noisier backgrounds with low available materials, thus affecting the detection of these very specific features. To address this, we assembled >1,500 genome-wide DNA methylation arrays from 13 independent datasets [1,31,45,46,49–58] making this one of the largest methylation compendium to study HCC to date. We harnessed this rich dataset by implementing a machine learning pipeline that searches, in an unbiased way, for significantly differentially methylated regions (DMRs) in HCC presenting several improvements. Firstly, considering regions spanning multiple CpG sites increases confidence as these can be more robustly measured in liquid biopsies in clinical settings. This procedure reduces the impact of eventual CpG site misdetection in the diagnosis and makes this more amenable for next-generation sequencing readouts, which measure all sites within the specified region. Secondly, training machine learning predictors with a training set equally representing tissue and liquid biopsies ensures the DMRs identified are representative of HCC tumours that can also be measured in ctDNA. Moreover, making this comparison against a cirrhotic background, instead of healthy liver samples, provides a more relevant clinical comparison. Very often patients who develop HCC also suffer from chronic liver disease and cirrhosis, and these are the backgrounds where existing non-invasive alternatives underperform. Lastly, to reduce potential analytical artefacts in the DMR biomarker discovery we processed the training dataset (859 samples from 6 different studies [31,45,46,56–58]) from raw data with the same pipeline and applied stringent filters to remove problematic measurements and account for potential confounders, such as sex, age, tumour purity and global methylation, often not considered by other studies. Additionally, we validated our approach using not only hold-out samples and cross-validated procedures, but also an assembled validation dataset (692 samples from 7 independent datasets [1,49–55]), which was never used for training and comprises differently and independently processed datasets, thus testing the robustness of our DMRs to diverse processing pipelines.

Our machine learning approach compared favourably against 12 HCC methylation signatures [1,31,47,68–75] across multiple datasets in both tissue and liquid biopsy samples. We harnessed this to derive a novel methylation fingerprint comprising 38 DMRs and combined it into a single diagnostic metric which detected HCC tissue samples in a validation dataset with 96% recall and 98% precision.

A limitation of our analysis is linked with the scarcity of cfDNA methylation samples. While this is ubiquitous across other independent studies, it limits the estimation and extrapolation of evaluation metrics, recall and precision, to other cohorts. To mitigate this, we thoroughly benchmarked our approach by assembling comprehensive and independent training and validation DNA methylation datasets. Specifically, we aimed to integrate as many liquid biopsy samples as possible, e.g. cfDNA analyses from healthy controls, sepsis and different tumours [57], and while not directly related with HCC these samples supported the utility of out approach, by for example showing it could correctly classify all healthy cfDNA samples. Of note, the DMR signature score also successfully identified 7 cfDNA samples (out of 11) from other tumours, including breast, lung and colorectal cancer.

This last point suggests that our ctDNA methylation signature and risk score have the potential for pan-cancer early diagnostics. Indeed other studies have shown that DNA methylation biomarkers can be used for the detection of different cancers, such as promoter methylation of the gene SEPT9 in colorectal cancer and HCC [26,43,57,84]. Gene expression analysis showed that several DMRs of our signature are significantly associated with transcriptional repression of multiple Zinc Finger Proteins (ZFNs) supporting a potential role of these regions in cancer progression and early onset [80,85]. Lastly, this approach can also be proposed to monitor HCC patients that have undergone therapies, such as surgical resection, radiofrequency ablation and chemoembolization, as a means of clinical follow-up to identify residual disease and guide treatment [12,86].

## Conclusions

In this study, we present an artificial intelligence pipeline that harnesses a comprehensive genome-wide DNA methylation resource to build a signature and a diagnostic score for HCC that benchmarks favourably against existing biomarkers. While further work to confirm the clinical utility of this approach is ongoing, it addresses important challenges of the design of reliable non-invasive diagnostic and monitoring approaches for HCC from liquid biopsies, to provide long sought-after alternatives to current suboptimal approaches.

## Methods

### DNA methylation datasets assembly and processing

DNA methylation samples from 6 different datasets [31,45,46,56–58] using Infinium HumanMethylation EPIC and 450K assays were processed using the R package minfi (v1.32.0) [62,64]. Datasets were integrated by considering the overlapping CpG probes between the two Infinium HumanMethylation assays (n=575,130). All datasets were merged into a single matrix containing signal intensities imported from the raw IDAT files and processed using the functional normalisation pipeline [63]. Lastly, the ratio between the methylation and unmethylated channels was calculated and exported as beta values (β) [EQ1] with an offset of 100 and rounded to 5 decimal places:

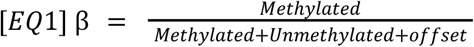

Altogether, we generated a single matrix of DNA methylation beta values spanning 452,567 CpG sites measured across 859 samples, integrating multiple studies processed from the raw signals using the same pipeline. For the downstream analyses several filtering steps were taken: (i) probes containing a single nucleotide polymorphism (SNP) in the CpG site or in the single nucleotide extension at a minor allele frequency (MAF) lower than 0.01 were excluded from downstream analysis; (ii) using maxprobes R package (v0.0.2,https://github.com/markgene/maxprobes) cross-reactive probes of the Illumina methylation arrays were removed [87–90]; (iii) CpG sites with missing values were discarded; (iv) we utilised an updated probe annotation mapped to the hg38 reference build and probes with no available alignments were not considered; and (v) to focus on biomarkers that are sex agnostic CpG sites mapping to sex chromosomes X and Y were removed from downstream analyses. The final filtered DNA methylation matrix covered a total of 390,445 CpG sites without any missing value across all samples.

### DNA methylation regions, CpG clusters

To identify DNA methylation regions, CpG clusters, we utilised a similar approach to the one described in Jaffe et al. [91]. Using the clusterMaker function from Bump Hunter R package (v1.30.0) [62,91] we identified CpG clusters with a maximum of 500 base-pairs (bp) distance between any 2 genomically consecutive CpG sites. Then we overlapped the CpG clusters with the filtered CpG sites defined previously and only considered CpG clusters with at least 3 CpG sites with measurements. A final CpG cluster matrix was defined by taking the mean of all filtered CpG sites within each cluster region, generating a DNA methylation matrix spanning 39,868 CpG clusters.

### Dimension reduction analyses

Dimension reduction analysis was performed using Principal Component Analysis (PCA) implemented in scikit-learn Python module (v0.24.0) [92].

### Balancing training samples sets

Considering the number of samples in each class, i.e. HCC, cirrhotic, cfDNA and tissue, the Train and & Test is highly unbalanced and this can generate artefacts that can limit an unbiased discovery of HCC biomarkers (Supplementary Figure 1a). Thus, we balanced the number of samples in each type for the training of the machine learning models. Since the limiting number of samples are from cfDNA samples, all samples available for HCC (n=22) and cirrhotic (n=22) from cfDNA are used for training. Then an equal number of samples (n=22) for HCC and cirrhotic are randomly sampled from the tissue samples, specifically Primary Tumour – Liver for HCC class, and Cirrhosis + HBV, Cirrhosis + HCV, Cirrhosis + AATD and Cirrhosis + EtOH for the cirrhotic class. Some cirrhotic tissue samples from the same dataset showed very distinct profiles diverging from other cirrhotic samples, thus we excluded them from the generation of the balanced dataset by considering only those cirrhotic samples from the GSE60753 dataset [58] with a Principal Component (PC) 2 lower than 200 (Supplementary Figure 8a and 8b). Taken together, a total of 88 samples, evenly separated by HCC and cirrhotic and cfDNA and tissue, are used for model training (Figure 2a). Within the leave-one-out cross-validation procedure, see below, in the cases where the test sample is a cfDNA sample this sample is not used for training and the total number of samples in each class is therefore reduced to 21, hence a total of 84 evenly distributed samples are used instead.

### Discovery of HCC biomarkers using support vector machine classifiers

The systematic search of DNA methylation biomarkers of HCC and benchmark against other independent sets of biomarkers [1,31,47,68–75] was performed within a leave-one-out cross-validation procedure across the 859 samples contained in the Train & Test dataset. In this procedure one sample at a time is left out for testing and the rest are used to build a balanced dataset (undersampling of the HCC and cirrhotic tissue samples) to identify differentially methylated regions (DMRs) predictive of HCC.

Firstly, with the balanced train dataset we defined DMRs using a multivariate linear regression model, LinearRegression class from scikit-learn (v0.24.0), that takes as dependent variables the mean methylation values of the 39,868 CpG clusters contained in the balanced dataset (Samples x CpG clusters) and as independent variable (Samples x 1) the binary classification if a sample is HCC (1) or not (0). Additionally, multiple potential confounding factors, covariates, are included in the model as independent variables: (i) binary variable representing sex (female), since this information in incomplete, we accurately estimated the sample sex using the methylation profiles and the R package minfi (v1.32.0) [62,64]; (ii) patient age, this is also largely unavailable and therefore we used [65,66] the R package watermelon [93] (v1.0.0) to estimate methylation age of the sample using their methylation profile and considered the Hannum [65] and Horvath [66] approaches; (iii) sample global methylation, to mitigate potential biases mediated by the sample overall methylation levels we calculated the sample mean methylation levels and considered it as another independent variable; (iv) tumour purity, this information is only available for the TCGA samples [56,76], CPE purity [67], and the varying levels of tumour purity affect the molecular measurements and thereby we included Train & Test PC5 in the model as a proxy to tumour purity estimations (Spearman’s rho 0.59, p-value 9.6e-37); and lastly (v) we included an intercept term. The full model is fitted and a beta coefficient is estimated for each independent variable. To statistically assess those CpG clusters that are significantly differentially methylated in HCC we also trained a smaller model (null hypothesis) that excludes the HCC status to test the hypothesis that the CpG cluster methylation status provides a significant increase in the classification power of HCC over the covariates. This is estimated using the log-likelihood ratio test for every CpG cluster and the p-values are then adjusted for multiple-hypothesis testing using the Benjamini-Hochberg False Discovery Rate (FDR). We complement this with a ANOVA differential CpG cluster methylation analysis performed with the *f_classif* function from the scikit-learn (v0.24.0) [92] module and statistical assessment using the F-scores associated p-values after adjusting for multiple hypothesis with FDR. Lastly, DMRs are defined as those CpG clusters with a ratio test and ANOVA FDR lower than 1%. This identified a median of 1,355 DMRs across the leave-one-out procedure.

Secondly, having identified DMRs in HCC we then estimate the most important DMRs to predict HCC by training linear support vector machines (LinearSVC) using a L1 regularization, with penalty parameter (C) set to 1.5, to reduce the number of DMRs considered in the model. DMRs with non-zero weights in the trained model are then defined as the most predictive DMRs to classify HCC samples. A median of 38 HCC predictive DMRs are identified per model across the 859 folds of the leave-one-out procedure, where 150 unique DMRs are found in at least 5% of all trained models (n=43).

### Benchmarking DMRs against other DNA methylation signatures

Within each fold of the leave-one-out cross-validation, the top predictive DMRs (DMRs with non-zero coefficients in the LinearSVC model) are used to train an independent ensemble model. The ensemble model includes Logistic and Ridge Linear classifiers both independently cross-validated to estimate the regularization parameter C and alpha, respectively. This is implemented using the VotingClassifier class from scikit-learn (v0.24.0) [92] using a soft voting modality, i.e. taking the argmax of the estimated probabilities to be HCC, and the LogisticRegressionCV and RidgeClassifierCV for the cross-validated logistic and linear classifiers, respectively. For training the ensemble all samples from the Train & Test dataset are used, apart from the one left out for testing. In contrast to the DMR discovery, training of the ensemble is not restricted to the balanced sample set. Lastly, the trained ensemble model is used to make a prediction of the HCC status of the test sample using the soft-voting.

A similar procedure is performed for the 12 independent HCC DNA methylation signatures, training an ensemble model per signature restricted to the CpG sites contained in the signature, and then a prediction is made about the HCC status of the test sample. All predictions on the test sample are stored and multiple evaluation metrics are calculated compared to the true label: confusion matrices, recall, precision, sensitivity, balanced accuracy, and Mathew correlation coefficients (MCC).

### Sequential feature selection

A forward greedy sequential feature selection procedure to iteratively find the optimal number of DMRs from the Train and Test dataset was performed using the SequentialFeatureSelector function implemented in the python module scikit-learn (v0.24.0) [92]. For this analysis only DMRs that were found in more than 5% of the leave-one-out optimal models (n = 43 models) were considered (n = 150 DMRs). Considering that 38 is the median optimal number of features in the leave-one-out cross-validated models (Figure 1c) and that the frequency of the DMRs is positively correlated with its absolute effect size (Supplementary Figure 2d), we train and tested LinearSVC models with a ranging number of CpG clusters from 1 to 38. For each LinearSVC we utilised a balanced dataset (see methods) for training and repeated this 30 times for each number of DMRs. In each model, predictions for the train, test and validation samples were performed and evaluated with precision and recall metrics (Supplementary Figure 3a).

### Linear regression models between gene expression and methylation

To identify potential associations between DMRs methylation and gene expression we utilised transcriptomics measurements [56,76] available for the liver TCGA samples contained in the Train & Test dataset (n=410). Within this subset we systematically tested linear associations between methylation profiles of the top 38 DMRs and 15,341 gene expression profiles using linear regression models implemented in the python module limix (v3.0.4) [94]. We defined the following linear mixed model [EQ2]:

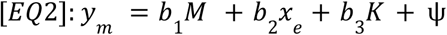

where *y* represents a vector of a single DMR methylation profile, *M* represents a matrix of covariates, *x* a gene expression vector of a single gene, *K* is the random effects term represented by the Kinship matrix of all the samples estimated using a linear kernel, andψis the noise term. The covariate matrix, *M*, contains several factors that might confound associations, similar to before: (i) global methylation; (ii) predicted patient sex; (iii) predicted patient age using both Hannum and Horvath methods; (iv) tumour purity, Train & Test PC5 used as a proxy; and (v) an intercept term. Gene expression measurements were standardized by subtracting the mean and dividing by the standard deviation. For each DMR and gene association a [EQ2] linear mixed model was fitted by minimising the residual sum of squares to estimate the parameters *b*_1_, *b*_2_ and *b*_3_. Statistical significance was assessed by performing a log likelihood ratio test between the full model [EQ2] and the null model which excludes the gene expression term (*b*_2_ *x*_*e*_), p-value was derived using a chi-square distribution with one degree of freedom and correction for multiple testing using FDR. A total of 582,958 DMR and gene expression associations were tested and 39 were found to be significant at a FDR < 10%.

### HCC DMR signature score

HCC linear risk score (DMR signature score) is a weighted sum of the methylation of the top 38 DMRs recurrently present with non-zero weights in the linear support vector machines (LinearSVCs) trained with the balanced sample sets in the leave-one-out cross-validation. The preponderance (weight) of each DMR is estimated using 1,000 permutations of the balanced dataset which are used to train a Ridge classifier with an alpha parameter set to 1. This ensured a regularisation of the model’s feature coefficients, while preserving them non-zero. The mean and standard deviation of each DMR is then calculated across all 1,000 iterations. The mean coefficients are then used in a weighted additive score where features with larger absolute coefficients have larger preponderance in the linear DMR signature score. Based on this feature set and weights a score is calculated for each sample using the sample-specific DMR methylation values. Recall and precision curves are generated using the risk score and the HCC status of the samples. Optimal threshold and precision and recall rates are estimated based on the best F1 metric possible along the curves. A similar approach is taken for the other 12 independent DNA methylation signatures, where CpG sites are used as features instead.

## Data Availability

Data and source code used to perform the analyses described in this study are provided as supplementary materials.

## Acknowledgements

We thank all the authors that made their data publicly available which allowed this study, particularly Prof. Keith D. Robertson and Dr. Ryan Hlady for kindly sharing raw data.

## Author Information

### Contributions

EG, JBPR and JC designed the study. EG performed all statistical analysis and data visualisation and MR, JBPR and JC advised. EG, MR, JBPR and JC wrote the manuscript. All authors read and approved the manuscript.

### Corresponding authors

Correspondence should be addressed to Joana Cardoso.

## Supplementary Figures

**Supplementary Figure 1.**
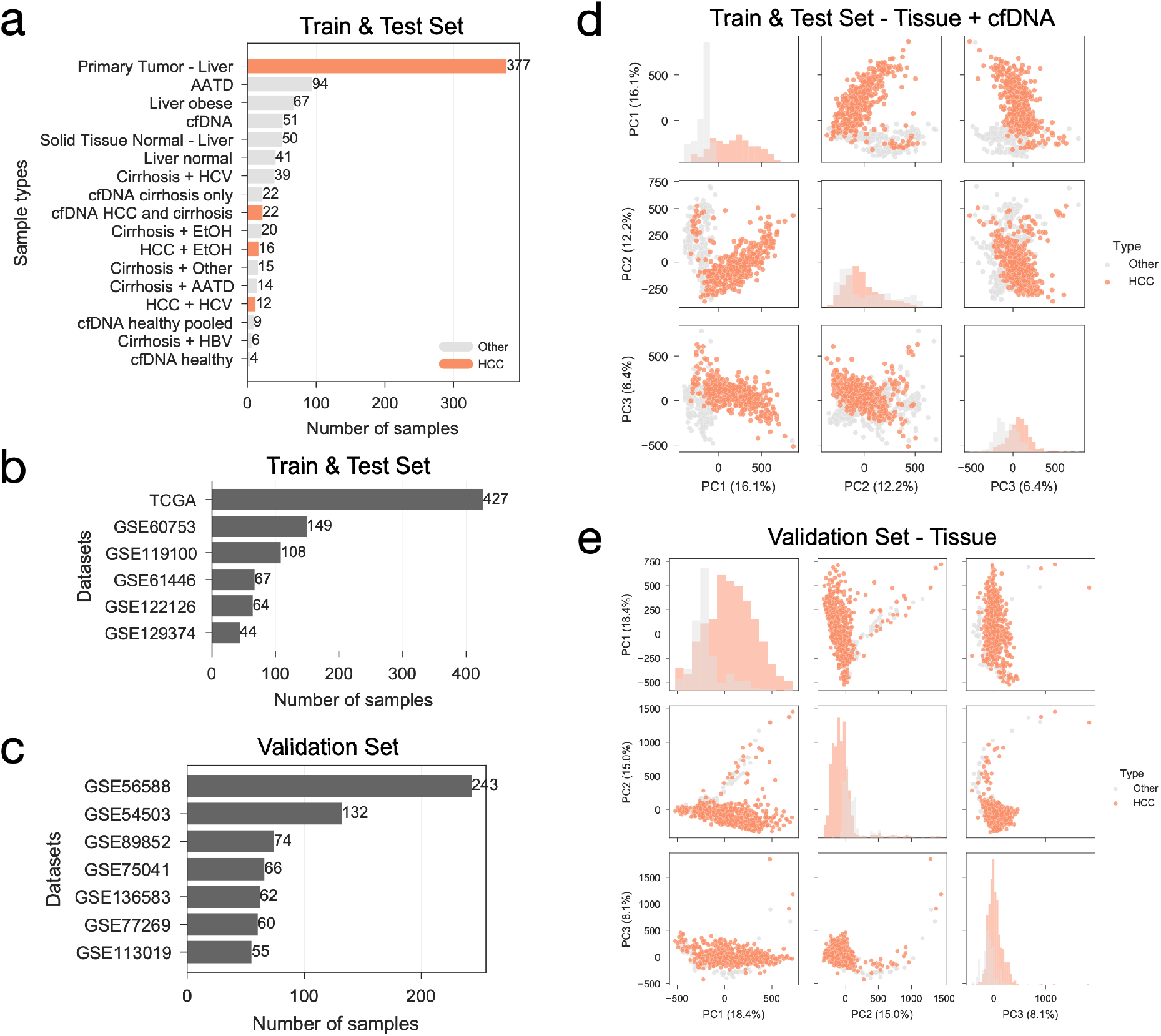
Overview of the DNA methylation datasets assembled. a) number of samples across different types, i.e. HCC, healthy, cirrhotic and other diseased liver samples. b) number of samples per study constituting the Train & Test dataset. c) similar to b), number of samples per study constituting the Validation dataset. d) principal component analysis (PCA) of the Train & Test dataset, plotting the first 3 principal components against each other (off-diagonal) and the distribution (diagonal). HCC samples are highlighted from the rest. e) similar to d), PCA of the Validation dataset.

**Supplementary Figure 2.**
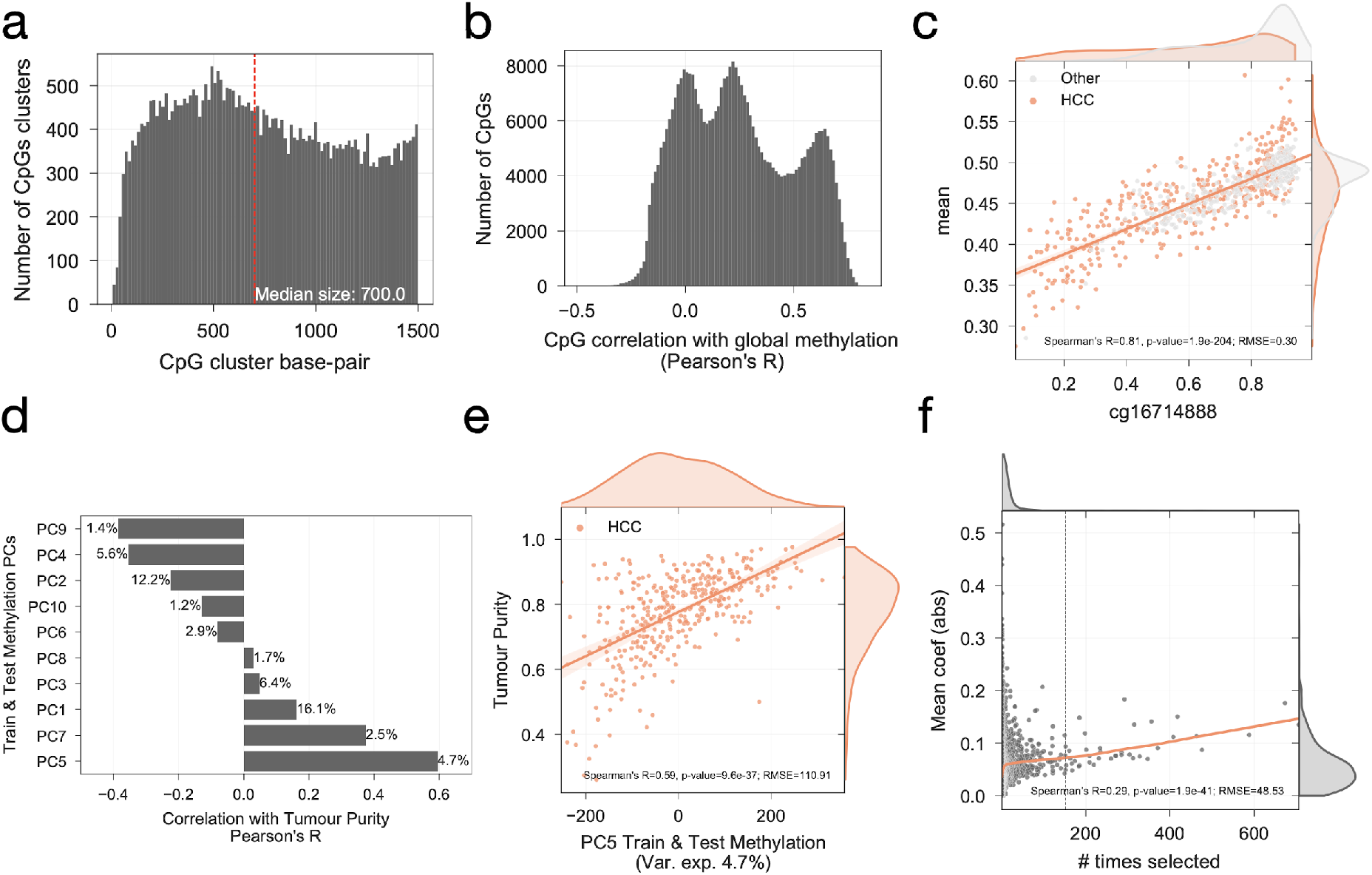
Information of DNA methylation features. a) distribution of the length of the CpG clusters. Median size of all clusters is represented as a vertical line. b) distribution of Pearson’s correlation coefficient (R) between all CpG sites contained in the Train & Test dataset and the overall sample mean methylation. c) top correlated CpG site with mean DNA methylation. HCC samples from the Train & Test dataset are highlighted in orange from the rest. d) Pearson’s correlation coefficients between the first 10 principal components (PCs) of the Train & Test methylation dataset with estimated tumour purity [67] for the HCC samples from the TCGA dataset [56]. PCs explained variance is reported next to the corresponding bar. e) Scatter and linear regression of top correlated Train & Test methylation PC, PC5, with TCGA HCC samples estimated tumour purity. f) number of times a DMR is present in the optimal LinearSVC model in the leave-one-out cross-validation procedure plotted against the mean absolute coefficient. Dashed vertical line represents the frequency cut-off of the top 38 DMRs.

**Supplementary Figure 3.**
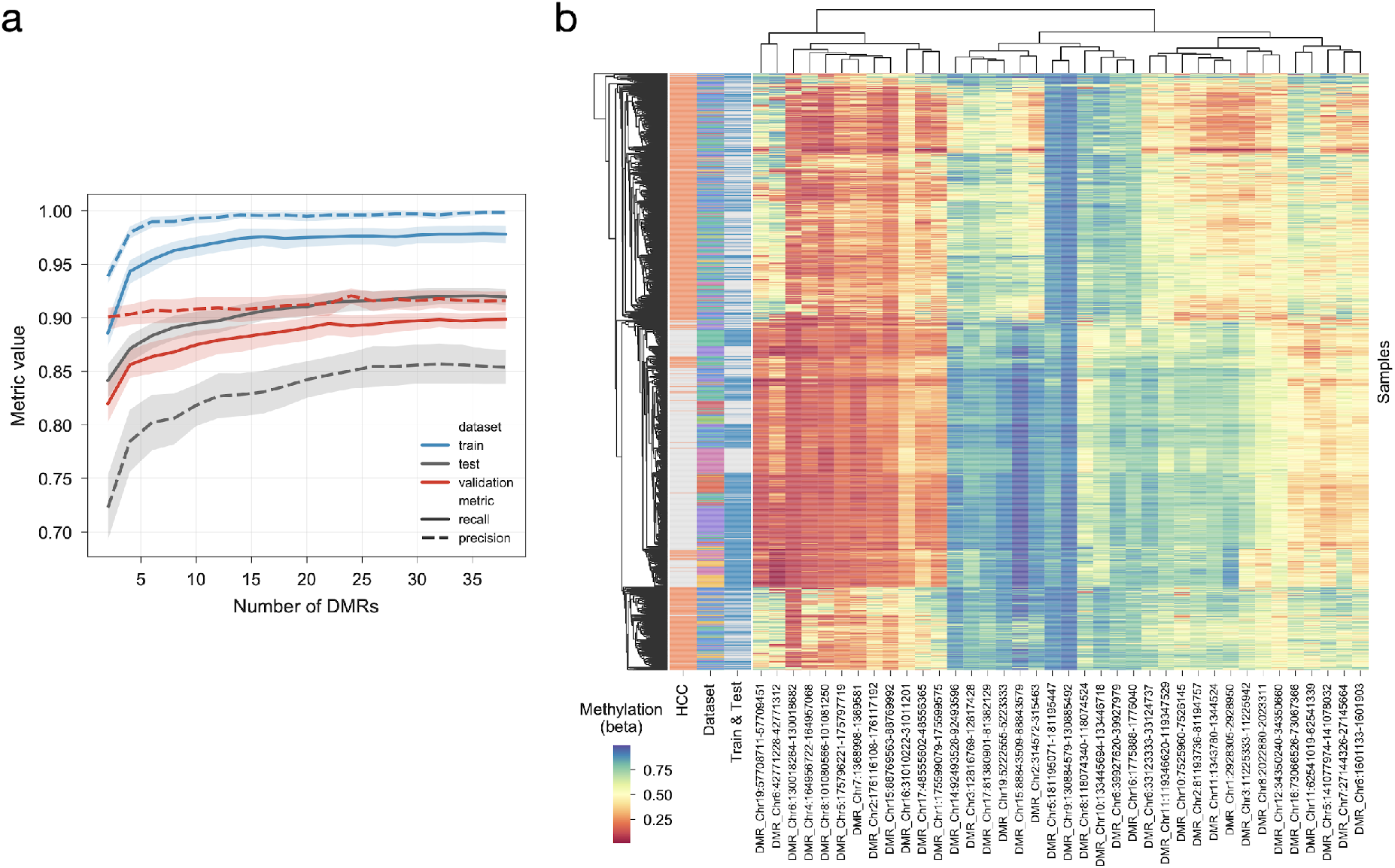
Top DNA methylation HCC biomarkers. a) greedy sequential DMR selection of the best DMR is selected to be added to the LinearSVC model. For each number of DMRs, 30 balanced train sets were generated and benchmarked. Models were trained with balanced train sets and used to predict the train, the test and the validation datasets. The number of features to be selected ranges from 1 to 38, where the latter represents the median number of features in the LinearSVC models. Error margins represent the 95th confidence interval. b) DNA methylation heatmap of top 38 DMRs across Train & Test and Validation datasets. Row colour annotations identify, in order: HCC samples (red) from the rest (gray); the different datasets used; and the samples that correspond to the Train & Test dataset (blue) from the Validation dataset (gray).

**Supplementary Figure 4.**
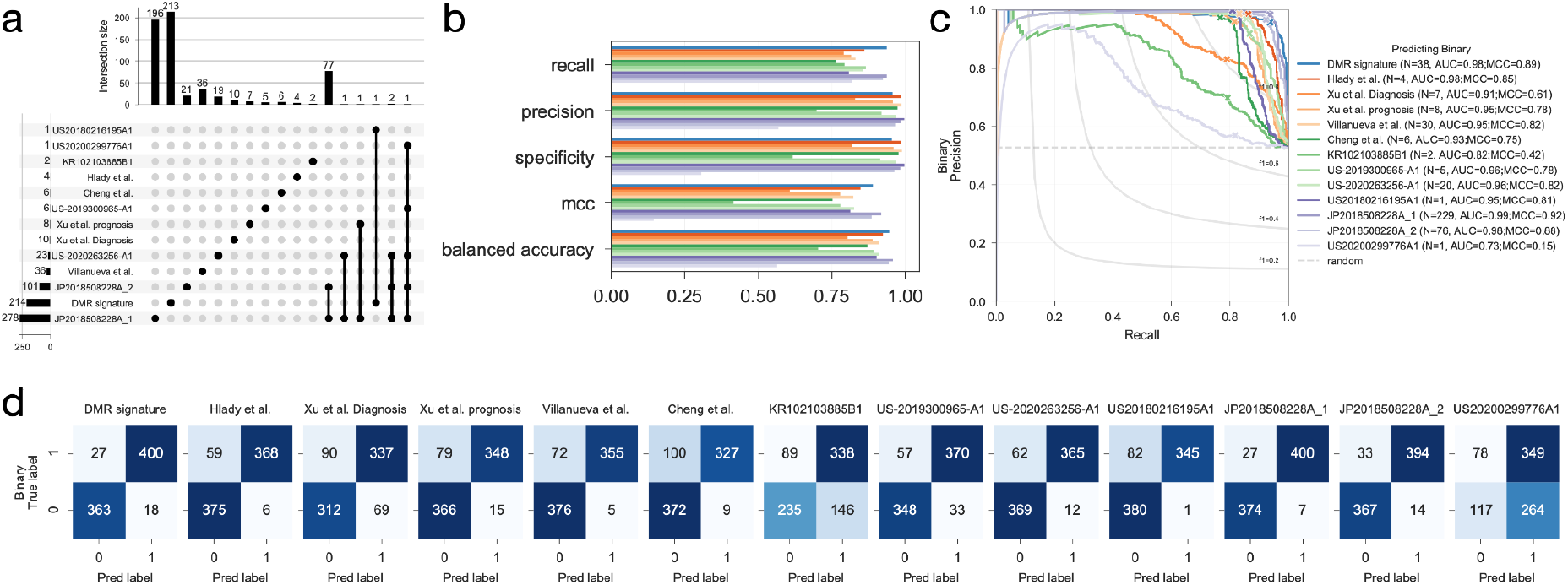
Train & Test dataset trained models performance. a) UpSet plot showing the intersection of the multiple independent HCC DNA methylation biomarker sets. Horizontal bars represent the total number of unique features in the respective signature. Vertical bars represent the number of features unique to the dataset, or in case of multiple signatures the number of overlapping features. The signatures selected to draw the barplots are identified with black circles. b) evaluation of the leave-one-out cross-validation procedure in the Train & Test dataset using the Recall, Precision, Specificity, Mathew’s Correlation Coefficient (MCC) and the balanced accuracy metrics. Each HCC DNA methylation feature set is coloured differently as in c). c) Precision-recall curves obtained by each feature set using the confidence scores of each sample belonging to the HCC class. Confidence scores are proportional to the sample distance to the hyperplane. Optimal F1-scores along the curves are marked with a “x”. Number of features in each dataset overlapping with the Train & Test dataset is specified in the label, as well as the area under the precision-recall curve and the MCC. Random precision is represented as a dashed horizontal line. d) confusion matrices of the predictions of each HCC DNA methylation biomarker set.

**Supplementary Figure 5.**
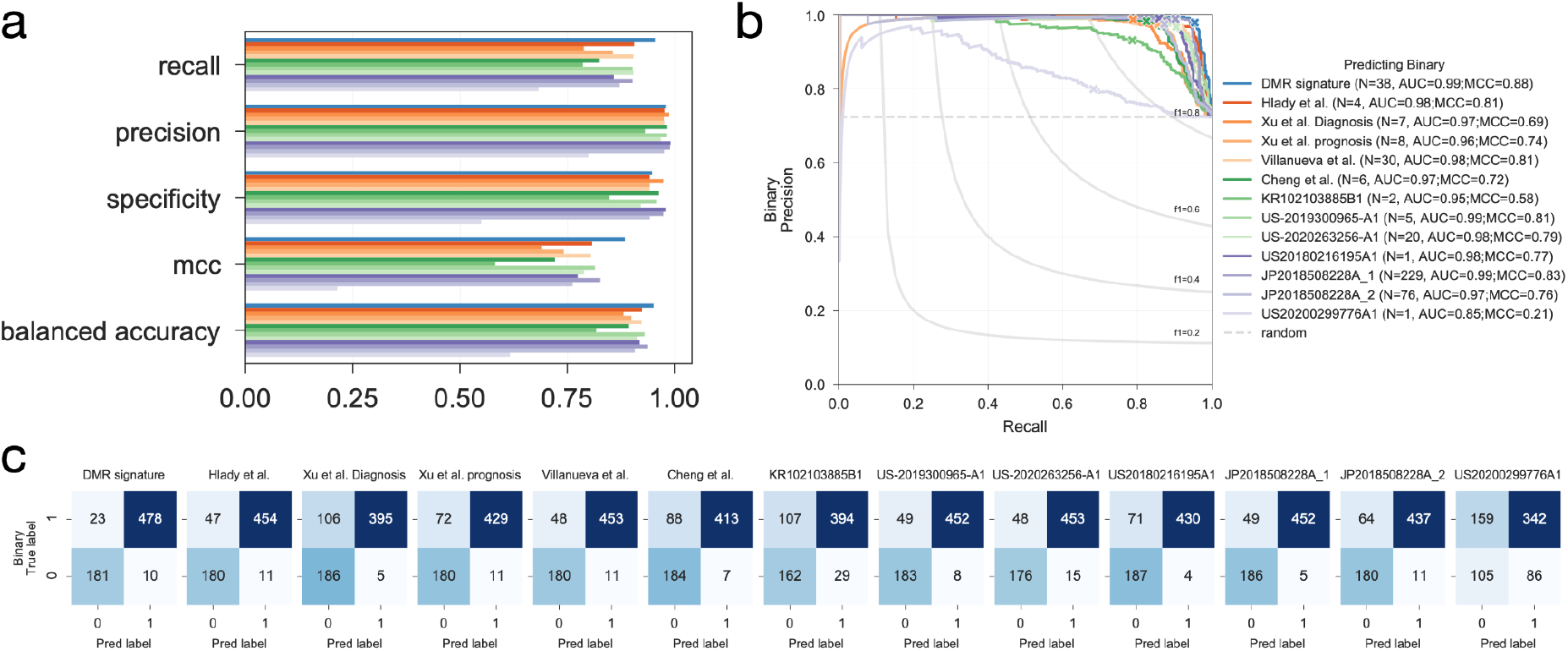
Validation dataset prediction performance. a) evaluation of the Validation dataset using an ensemble of logistic and linear ridge classification models trained with the Train & Test dataset. Recall, Precision, Specificity, Mathew’s Correlation Coefficient and balanced accuracy metrics are presented. b) Precision-recall curves obtained by each feature set using the confidence scores of each sample belonging to the HCC class. Confidence scores are proportional to the sample distance to the hyperplane. Optimal F1-scores along the curves are marked with a “x”. Number of features in each dataset overlapping with the Validation dataset is specified in the label, as well as the area under the precision-recall curve and the MCC. Random precision is represented as a dashed horizontal line. c) confusion matrices of the predictions of each HCC DNA methylation biomarker set.

**Supplementary Figure 6.**
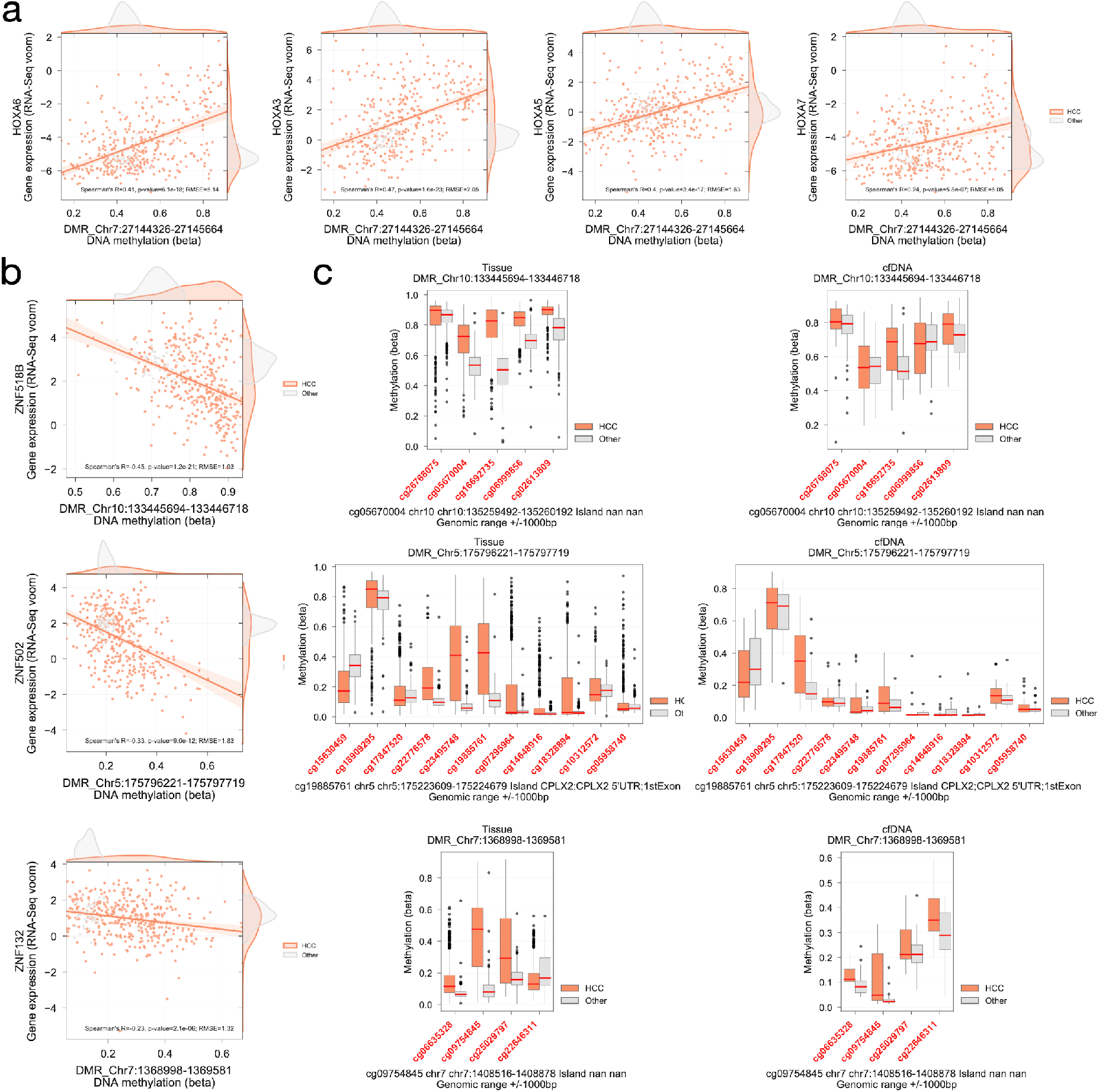
DNA methylation distribution of representative top CpG clusters predictive of HCC biomarkers. a) significant associations between DMR Chr7:27144326-27145664 and gene expression of multiple HOXA members. b) similar to a), but instead showing significant associations between multiple Zinc Finger Proteins and DMRs Chr10:133445694-133446718, Chr5:175796221-175797719 and Chr7:1368998-1369581. c) DNA methylation (beta) of CpG sites contained in a range of 1,000 base-pairs of the DMRs presented in b). Genomic information of the location of the DMR to genes and CpG islands is provided below. Left panel shows the distribution of the DNA methylation in the tissue samples of the Test & Train datasets, while the right panel shows the distribution in the cfDNA samples of the same CpG sites. CpG sites are sorted in ascending order according to their genomic location. In red are CpG sites that are contained in the DMR.

**Supplementary Figure 7.**
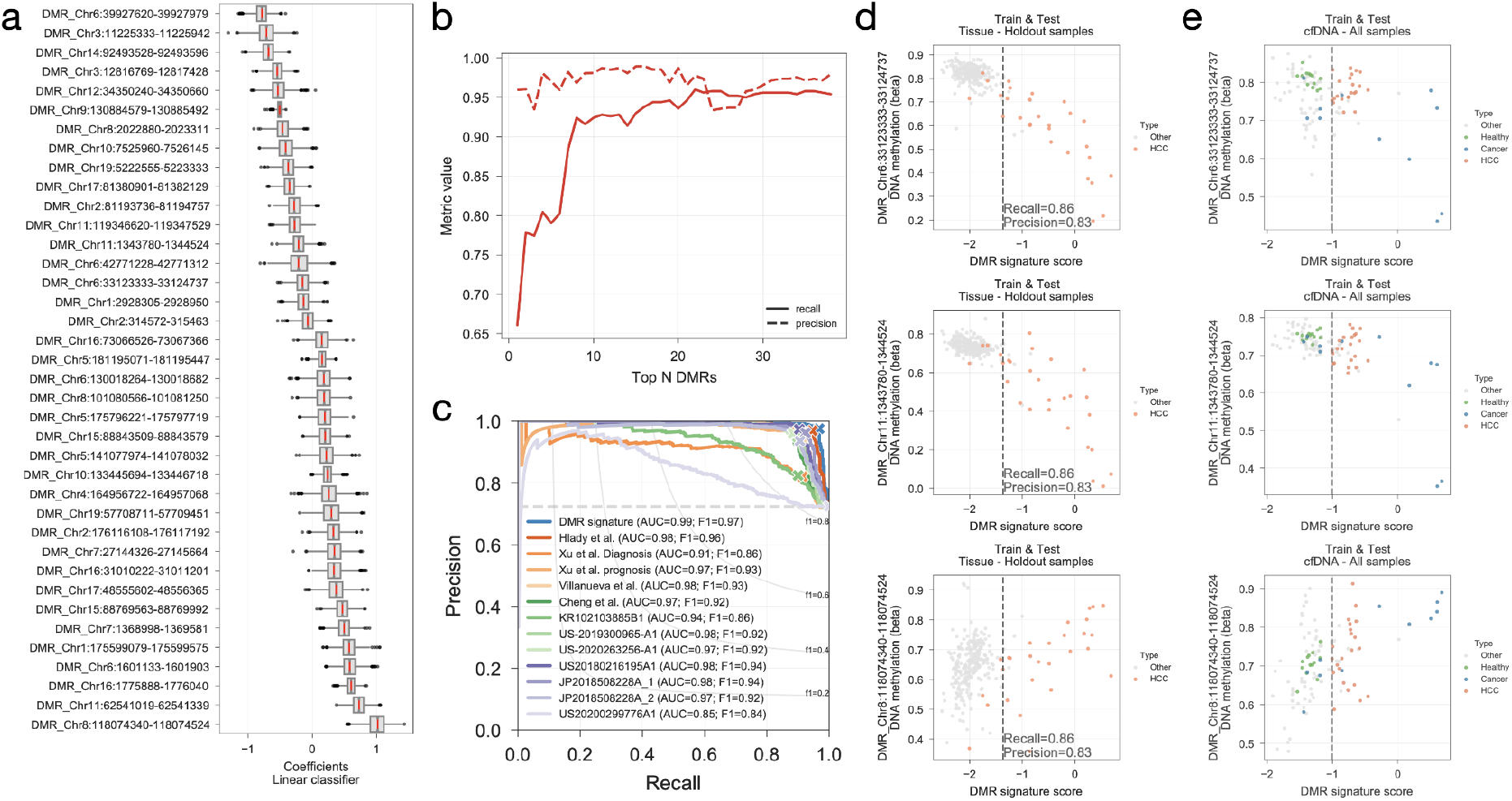
HCC DNA methylation risk score. a) distributions of DMR coefficients across the 1,000 permutations of the balanced datasets. b) Recall and precision metrics of the predicted HCC labels in the Validation dataset using an iterative addition of the top N DMRs ranked descendingly according to their absolute mean coefficients reported in a). Train & Test dataset is used to train a cross-validated ensemble of logistic and linear classifiers to predict HCC samples in the Validation dataset. c) Precision-recall curves of the Validation samples calculated using linear risk score estimated from the mean coefficients obtained in the 1,000 permutation analysis. d) HCC DMR signature score calculated for all the samples in the Train & test dataset which were not used for the identification of the DMR signature and score nor their weights. DMR signature score plotted against three representative HCC DNA methylation biomarkers. HCC classification threshold is represented by a dashed vertical line and precision and recall rates are reported. e) Similar to d), instead only cfDNA samples are utilised and cfDNA samples from patients with other cancers (marked as blue and labeled as “Cancer”) are also considered as a positive event. cfDNA samples from healthy controls are marked in green (“Healthy”).

**Supplementary Figure 8.**
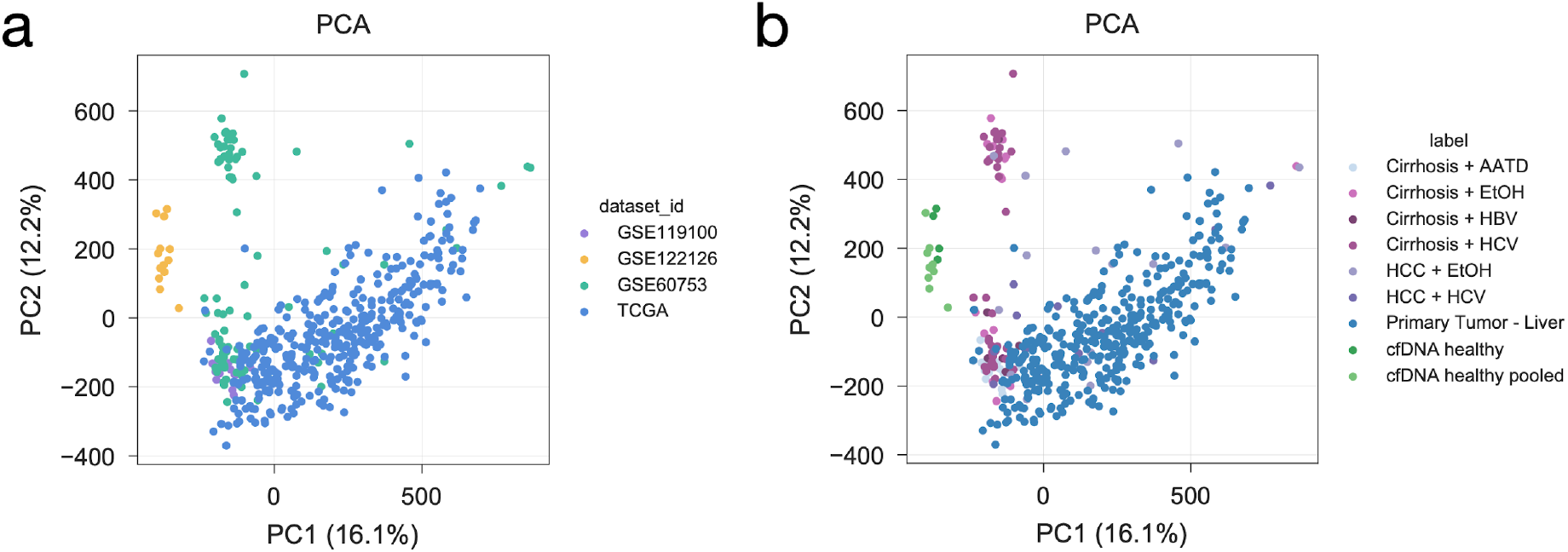
Train and Test PCA analysis. First and second principal component of the Train & Test dataset samples coloured by a) original dataset, and b) sample type.

## Supplementary Information

**Supplementary Table 1**. List of samples and original studies that comprise the Train & Test and Validation datasets.

**Supplementary Table 2**. DMRs present in the optimal LinearSVC across the leave-one-out cross-validation procedure, frequency and effect sizes are reported.

**Supplementary Table 3**. Genomic information for all CpG sites mapping to the top 38 DMRs.

**Supplementary Table 4**. Significant CpG cluster - Gene expression associations.

**Supplementary Table 5**. HCC linear risk scores for the Test & Train and Validation datasets.

**Supplementary File 1**. Analyses source code.

## References

1. Villanueva A, Portela A, Sayols S, Battiston C, Hoshida Y, Méndez-González J, et al. DNA methylation-based prognosis and epidrivers in hepatocellular carcinoma. Hepatology. 2015;61:1945–56.

2. National Cancer Institute. SEER Cancer Stat Facts: Liver and Intrahepatic Bile Duct Cancer [Internet]. seer.cancer.gov. [cited 2021 Mar 10]. Available from: https://seer.cancer.gov/statfacts/html/livibd.html

3. Vogel A, Cervantes A, Chau I, Daniele B, Llovet JM, Meyer T, et al. Hepatocellular carcinoma: ESMO Clinical Practice Guidelines for diagnosis, treatment and follow-up. Ann Oncol. 2018;29:iv238–55.

4. European Association for the Study of the Liver. Electronic address, European Association for the Study of the Liver. EASL Clinical Practice Guidelines: Management of hepatocellular carcinoma. J Hepatol. 2018;69:182–236.

5. Llovet JM, Kelley RK, Villanueva A, Singal AG, Pikarsky E, Roayaie S, et al. Hepatocellular carcinoma. Nat Rev Dis Primers. Springer Science and Business Media LLC; 2021;7:6.

6. Lambert M-P, Paliwal A, Vaissière T, Chemin I, Zoulim F, Tommasino M, et al. Aberrant DNA methylation distinguishes hepatocellular carcinoma associated with HBV and HCV infection and alcohol intake. J Hepatol. 2011;54:705–15.

7. Bialecki ES, Di Bisceglie AM. Diagnosis of hepatocellular carcinoma. HPB. 2005;7:26–34.

8. Brar G, Greten TF, Graubard BI. Hepatocellular Carcinoma Survival by Etiology: A SEER-Medicare Database Analysis. Hepatology [Internet]. Wiley Online Library; 2020; Available from: https://aasldpubs.onlinelibrary.wiley.com/doi/abs/10.1002/hep4.1564

9. Giannini EG, Cucchetti A, Erroi V, Garuti F, Odaldi F, Trevisani F. Surveillance for early diagnosis of hepatocellular carcinoma: how best to do it? World J Gastroenterol. 2013;19:8808–21.

10. Yang JD, Hainaut P, Gores GJ, Amadou A, Plymoth A, Roberts LR. A global view of hepatocellular carcinoma: trends, risk, prevention and management. Nat Rev Gastroenterol Hepatol. Nature Publishing Group; 2019;16:589–604.

11. Ayuso C, Rimola J, Vilana R, Burrel M, Darnell A, García-Criado Á, et al. Diagnosis and staging of hepatocellular carcinoma (HCC): current guidelines. Eur J Radiol. 2018;101:72–81.

12. Marrero JA, Kulik LM, Sirlin CB, Zhu AX, Finn RS, Abecassis MM, et al. Diagnosis, Staging, and Management of Hepatocellular Carcinoma: 2018 Practice Guidance by the American Association for the Study of Liver Diseases. Hepatology. 2018;68:723–50.

13. Qu C, Wang Y, Wang P, Chen K, Wang M, Zeng H, et al. Detection of early-stage hepatocellular carcinoma in asymptomatic HBsAg-seropositive individuals by liquid biopsy. Proc Natl Acad Sci U S A. 2019;116:6308–12.

14. Banini BA, Sanyal AJ. The use of cell free DNA in the diagnosis of HCC. Hepatoma Res [Internet]. 2019;5. Available from: http://dx.doi.org/10.20517/2394-5079.2019.30

15. Degroote H, Callebout E, Iesari S, Dekervel J, Schreiber J, Pirenne J, et al. Extended criteria for liver transplantation in hepatocellular carcinoma. A retrospective, multicentric validation study in Belgium. Surg Oncol. 2020;33:231–8.

16. Tzartzeva K, Obi J, Rich NE, Parikh ND, Marrero JA, Yopp A, et al. Surveillance imaging and alpha fetoprotein for early detection of hepatocellular carcinoma in patients with cirrhosis: A meta-analysis. Gastroenterology. Elsevier BV; 2018;154:1706–18.e1.

17. Trevisani F, D’Intino PE, Morselli-Labate AM, Mazzella G, Accogli E, Caraceni P, et al. Serum alpha-fetoprotein for diagnosis of hepatocellular carcinoma in patients with chronic liver disease: influence of HBsAg and anti-HCV status. J Hepatol. 2001;34:570–5.

18. Diaz LA Jr, Bardelli A. Liquid biopsies: genotyping circulating tumor DNA. J Clin Oncol. 2014;32:579–86.

19. Siravegna G, Marsoni S, Siena S, Bardelli A. Integrating liquid biopsies into the management of cancer. Nat Rev Clin Oncol. 2017;14:531–48.

20. Wan JCM, Massie C, Garcia-Corbacho J, Mouliere F, Brenton JD, Caldas C, et al. Liquid biopsies come of age: towards implementation of circulating tumour DNA. Nat Rev Cancer. 2017;17:223–38.

21. Bettegowda C, Sausen M, Leary RJ, Kinde I, Wang Y, Agrawal N, et al. Detection of circulating tumor DNA in early- and late-stage human malignancies. Sci Transl Med. 2014;6:224ra24.

22. Diehl F, Li M, Dressman D, He Y, Shen D, Szabo S, et al. Detection and quantification of mutations in the plasma of patients with colorectal tumors. Proc Natl Acad Sci U S A. 2005;102:16368–73.

23. Dawson S-J, Tsui DWY, Murtaza M, Biggs H, Rueda OM, Chin S-F, et al. Analysis of circulating tumor DNA to monitor metastatic breast cancer. N Engl J Med. 2013;368:1199–209.

24. Li J, Zhao S, Lee M, Yin Y, Li J, Zhou Y, et al. Reliable tumor detection by whole-genome methylation sequencing of cell-free DNA in cerebrospinal fluid of pediatric medulloblastoma. Sci Adv [Internet]. 2020;6. Available from: http://dx.doi.org/10.1126/sciadv.abb5427

25. Murtaza M, Dawson S-J, Tsui DWY, Gale D, Forshew T, Piskorz AM, et al. Non-invasive analysis of acquired resistance to cancer therapy by sequencing of plasma DNA. Nature. 2013;497:108–12.

26. Liu MC, Klein E, Hubbell E, Maddala T, Aravanis AM, Beausang JF, et al. Plasma cell-free DNA (cfDNA) assays for early multi-cancer detection: The circulating cell-free genome atlas (CCGA) study. Ann Oncol. Elsevier; 2018;29:viii14.

27. Oussalah A, Rischer S, Bensenane M, Conroy G, Filhine-Tresarrieu P, Debard R, et al. Plasma mSEPT9: A Novel Circulating Cell-free DNA-Based Epigenetic Biomarker to Diagnose Hepatocellular Carcinoma. EBioMedicine. 2018;30:138–47.

28. Cescon DW, Bratman SV, Chan SM, Siu LL. Circulating tumor DNA and liquid biopsy in oncology. Nature Cancer. 2020;1:276–90.

29. Ignatiadis M, Sledge GW, Jeffrey SS. Liquid biopsy enters the clinic - implementation issues and future challenges. Nat Rev Clin Oncol [Internet]. 2021; Available from: http://dx.doi.org/10.1038/s41571-020-00457-x

30. Shen SY, Singhania R, Fehringer G, Chakravarthy A, Roehrl MHA, Chadwick D, et al. Sensitive tumour detection and classification using plasma cell-free DNA methylomes. Nature. 2018;563:579–83.

31. Hlady RA, Zhao X, Pan X, Yang JD, Ahmed F, Antwi SO, et al. Genome-wide discovery and validation of diagnostic DNA methylation-based biomarkers for hepatocellular cancer detection in circulating cell free DNA. Theranostics. 2019;9:7239–50.

32. Turner NC, Kingston B, Kilburn LS, Kernaghan S, Wardley AM, Macpherson IR, et al. Circulating tumour DNA analysis to direct therapy in advanced breast cancer (plasmaMATCH): a multicentre, multicohort, phase 2a, platform trial. Lancet Oncol. 2020;21:1296–308.

33. Chabon JJ, Simmons AD, Lovejoy AF, Esfahani MS, Newman AM, Haringsma HJ, et al. Circulating tumour DNA profiling reveals heterogeneity of EGFR inhibitor resistance mechanisms in lung cancer patients [Internet]. Nature Communications. 2016. Available from: http://dx.doi.org/10.1038/ncomms11815

34. Abbosh C, Birkbak NJ, Wilson GA, Jamal-Hanjani M, Constantin T, Salari R, et al. Phylogenetic ctDNA analysis depicts early-stage lung cancer evolution. Nature. 2017;545:446–51.

35. Hattori N, Ushijima T. Epigenetic impact of infection on carcinogenesis: mechanisms and applications. Genome Med. 2016;8:10.

36. Koch A, Joosten SC, Feng Z, de Ruijter TC, Draht MX, Melotte V, et al. Analysis of DNA methylation in cancer: location revisited. Nat Rev Clin Oncol. 2018;15:459–66.

37. Thienpont B, Steinbacher J, Zhao H, D’Anna F, Kuchnio A, Ploumakis A, et al. Tumour hypoxia causes DNA hypermethylation by reducing TET activity. Nature. 2016;537:63–8.

38. Irizarry RA, Ladd-Acosta C, Wen B, Wu Z, Montano C, Onyango P, et al. The human colon cancer methylome shows similar hypo- and hypermethylation at conserved tissue-specific CpG island shores. Nat Genet. 2009;41:178–86.

39. Teschendorff AE, Widschwendter M. Differential variability improves the identification of cancer risk markers in DNA methylation studies profiling precursor cancer lesions. Bioinformatics. 2012;28:1487–94.

40. Teschendorff AE, Jones A, Fiegl H, Sargent A, Zhuang JJ, Kitchener HC, et al. Epigenetic variability in cells of normal cytology is associated with the risk of future morphological transformation. Genome Med. 2012;4:24.

41. Zhuang J, Jones A, Lee S-H, Ng E, Fiegl H, Zikan M, et al. The dynamics and prognostic potential of DNA methylation changes at stem cell gene loci in women’s cancer. PLoS Genet. 2012;8:e1002517.

42. Hughes LAE, Melotte V, de Schrijver J, de Maat M, Smit Vthbm, Bovée JVMG, et al. The CpG island methylator phenotype: what’s in a name? Cancer Res. 2013;73:5858–68.

43. Liu MC, Oxnard GR, Klein EA, Swanton C, Seiden MV, Cummings SR, et al. Sensitive and specific multi-cancer detection and localization using methylation signatures in cell-free DNA. Ann Oncol. 2020;31:745–59.

44. deVos T, Tetzner R, Model F, Weiss G, Schuster M, Distler J, et al. Circulating methylated SEPT9 DNA in plasma is a biomarker for colorectal cancer. Clin Chem. 2009;55:1337–46.

45. Bonder MJ, Kasela S, Kals M, Tamm R, Lokk K, Barragan I, et al. Genetic and epigenetic regulation of gene expression in fetal and adult human livers. BMC Genomics. 2014;15:860.

46. Wang L, Marek GW 3rd, Hlady RA, Wagner RT, Zhao X, Clark VC, et al. Alpha-1 Antitrypsin Deficiency Liver Disease, Mutational Homogeneity Modulated by Epigenetic Heterogeneity With Links to Obesity. Hepatology. 2019;70:51–66.

47. Cheng J, Wei D, Ji Y, Chen L, Yang L, Li G, et al. Integrative analysis of DNA methylation and gene expression reveals hepatocellular carcinoma-specific diagnostic biomarkers. Genome Med. genomemedicine.biomedcentral …; 2018;10:42.

48. Li R, Shui L, Jia J, Wu C. Construction and Validation of Novel Diagnostic and Prognostic DNA Methylation Signatures for Hepatocellular Carcinoma. Front Genet. 2020;11:906.

49. Kuramoto J, Arai E, Tian Y, Funahashi N, Hiramoto M, Nammo T, et al. Genome-wide DNA methylation analysis during non-alcoholic steatohepatitis-related multistage hepatocarcinogenesis: comparison with hepatitis virus-related carcinogenesis. Carcinogenesis. 2017;38:261–70.

50. Tian Y, Arai E, Makiuchi S, Tsuda N, Kuramoto J, Ohara K, et al. Aberrant DNA methylation results in altered gene expression in non-alcoholic steatohepatitis-related hepatocellular carcinomas. J Cancer Res Clin Oncol. Springer Science and Business Media LLC; 2020;146:2461–77.

51. Yang Y, Chen L, Gu J, Zhang H, Yuan J, Lian Q, et al. Recurrently deregulated lncRNAs in hepatocellular carcinoma. Nat Commun. 2017;8:14421.

52. Qiu J, Peng B, Tang Y, Qian Y, Guo P, Li M, et al. CpG methylation signature predicts recurrence in Early-stage hepatocellular carcinoma: Results from a multicenter study. J Clin Oncol. American Society of Clinical Oncology (ASCO); 2017;35:734–42.

53. Shen J, Wang S, Zhang Y-J, Wu H-C, Kibriya MG, Jasmine F, et al. Exploring genome-wide DNA methylation profiles altered in hepatocellular carcinoma using Infinium HumanMethylation 450 BeadChips. Epigenetics. Informa UK Limited; 2013;8:34–43.

54. Shen J, Wang S, Siegel AB, Remotti H, Wang Q, Sirosh I, et al. Genome-wide expression of MicroRNAs is regulated by DNA methylation in hepatocarcinogenesis. Gastroenterol Res Pract. Hindawi Limited; 2015;2015:230642.

55. Shimada S, Mogushi K, Akiyama Y, Furuyama T, Watanabe S, Ogura T, et al. Comprehensive molecular and immunological characterization of hepatocellular carcinoma. EBioMedicine. 2019;40:457–70.

56. Cancer Genome Atlas Research Network. Electronic address, Cancer Genome Atlas Research Network. Comprehensive and Integrative Genomic Characterization of Hepatocellular Carcinoma. Cell. Elsevier BV; 2017;169:1327–41.e23.

57. Moss J, Magenheim J, Neiman D, Zemmour H, Loyfer N, Korach A, et al. Comprehensive human cell-type methylation atlas reveals origins of circulating cell-free DNA in health and disease. Nat Commun. 2018;9:5068.

58. Hlady RA, Tiedemann RL, Puszyk W, Zendejas I, Roberts LR, Choi J-H, et al. Epigenetic signatures of alcohol abuse and hepatitis infection during human hepatocarcinogenesis. Oncotarget. 2014;5:9425–43.

59. Edgar R, Domrachev M, Lash AE. Gene Expression Omnibus: NCBI gene expression and hybridization array data repository. Nucleic Acids Res. 2002;30:207–10.

60. Barrett T, Wilhite SE, Ledoux P, Evangelista C, Kim IF, Tomashevsky M, et al. NCBI GEO: archive for functional genomics data sets—update. Nucleic Acids Res. Oxford University Press; 2013;41:D991–5.

61. Athar A, Füllgrabe A, George N, Iqbal H, Huerta L, Ali A, et al. ArrayExpress update – from bulk to single-cell expression data. Nucleic Acids Res. Oxford University Press (OUP); 2019;47:D711–5.

62. Aryee MJ, Jaffe AE, Corrada-Bravo H, Ladd-Acosta C, Feinberg AP, Hansen KD, et al. Minfi: a flexible and comprehensive Bioconductor package for the analysis of Infinium DNA methylation microarrays. Bioinformatics. Oxford University Press (OUP); 2014;30:1363–9.

63. Fortin J-P, Labbe A, Lemire M, Zanke BW, Hudson TJ, Fertig EJ, et al. Functional normalization of 450k methylation array data improves replication in large cancer studies. Genome Biol. Springer Science and Business Media LLC; 2014;15:503.

64. Fortin J-P, Triche TJ Jr, Hansen KD. Preprocessing, normalization and integration of the Illumina HumanMethylationEPIC array with minfi. Bioinformatics. Oxford University Press (OUP); 2016;btw691.

65. Hannum G, Guinney J, Zhao L, Zhang L, Hughes G, Sadda S, et al. Genome-wide methylation profiles reveal quantitative views of human aging rates. Mol Cell. 2013;49:359–67.

66. Horvath S. DNA methylation age of human tissues and cell types. Genome Biol. 2013;14:R115.

67. Aran D, Sirota M, Butte AJ. Systematic pan-cancer analysis of tumour purity. Nat Commun. 2015;6:8971.

68. Xu R-H, Wei W, Krawczyk M, Wang W, Luo H, Flagg K, et al. Circulating tumour DNA methylation markers for diagnosis and prognosis of hepatocellular carcinoma. Nat Mater. nature.com; 2017;16:1155–61.

69. 김영준,&김다원,&하정실. Method for Identifying Whether Biological Sample is Derived from Liver Tissue [Internet]. Patent. 2020 [cited 2021 Jan 28]. Available from: https://patentimages.storage.googleapis.com/cb/94/7f/9c5a922f5bb020/KR102103885B1.pd f

70. Zhang K, Hou R, Zheng L. Liver cancer methylation markers and uses thereof [Internet]. US Patent. 2019 [cited 2021 Jan 28]. Available from: https://patentimages.storage.googleapis.com/e4/51/a6/677243789f7101/US20190300965A1.pdf

71. 张康,&侯睿,&郑良宏. Methylation markers for diagnosis of hepatocellular carcinoma and lung cancer [Internet]. Patent. 2019 [cited 2021 Jan 28]. Available from: https://patentimages.storage.googleapis.com/7a/89/7c/fcbb677b5ca5c8/CN110603329A.pdf

72. Zhang K, Hou R, Zheng L. Methylation markers for diagnosing hepatocellular carcinoma and lung cancer [Internet]. US Patent. 2020 [cited 2021 Jan 28]. Available from: https://patentimages.storage.googleapis.com/ee/37/45/2922038f188d5b/US20200263256A1.pdf

73. Elnitski LL, Margolin G. Cancer detection methods [Internet]. US Patent. 2018 [cited 2021 Jan 28]. Available from: https://patentimages.storage.googleapis.com/06/6d/bc/132737d19291fc/US20180216195A1.pdf

74. チャン,カン,&ホウ,ルイ,&チェン,リャンホン. Method and system for determining cancer status [Internet]. Patent. 2018 [cited 2021 Jan 28]. Available from: https://patentimages.storage.googleapis.com/33/13/b1/b9f6f046512a23/JP2018508228A.pdf

75. Zhang K, Hou R, Zheng L. Solid tumor methylation markers and uses thereof [Internet]. US Patent. 2020 [cited 2021 Jan 28]. Available from: https://patentimages.storage.googleapis.com/2f/72/d6/01f7b19bf41bbb/US20200299776A1.pdf

76. Rahman M, Jackson LK, Johnson WE, Li DY, Bild AH, Piccolo SR. Alternative preprocessing of RNA-Sequencing data in The Cancer Genome Atlas leads to improved analysis results. Bioinformatics. 2015;31:3666–72.

77. Novak P, Jensen T, Oshiro MM, Wozniak RJ, Nouzova M, Watts GS, et al. Epigenetic inactivation of the HOXA gene cluster in breast cancer. Cancer Res. 2006;66:10664–70.

78. Wang F, Yang H, Deng Z, Su Y, Fang Q, Yin Z. HOX Antisense lincRNA HOXA-AS2 Promotes Tumorigenesis of Hepatocellular Carcinoma. Cell Physiol Biochem. 2016;40:287–96.

79. Hajra KM, Chen DY-S, Fearon ER. The SLUG zinc-finger protein represses E-cadherin in breast cancer. Cancer Res. 2002;62:1613–8.

80. Cassandri M, Smirnov A, Novelli F, Pitolli C, Agostini M, Malewicz M, et al. Zinc-finger proteins in health and disease. Cell Death Discov. 2017;3:17071.

81. Gimeno-Valiente F, Riffo-Campos ÁL, Vallet-Sánchez A, Siscar-Lewin S, Gambardella V, Tarazona N, et al. ZNF518B gene up-regulation promotes dissemination of tumour cells and is governed by epigenetic mechanisms in colorectal cancer. Sci Rep. 2019;9:9339.

82. Sheridan C. Investors keep the faith in cancer liquid biopsies. Nat Biotechnol. 2019;37:972–4.

83. Deveson IW, Gong B, Lai K, LoCoco JS, Richmond TA, Schageman J, et al. Evaluating the analytical validity of circulating tumor DNA sequencing assays for precision oncology. Nat Biotechnol [Internet]. 2021; Available from: http://dx.doi.org/10.1038/s41587-021-00857-z

84. Kotoh Y, Suehiro Y, Saeki I, Hoshida T, Maeda M, Iwamoto T, et al. Novel Liquid Biopsy Test Based on a Sensitive Methylated SEPT9 Assay for Diagnosing Hepatocellular Carcinoma. Hepatol Commun. 2020;4:461–70.

85. Jen J, Wang Y-C. Zinc finger proteins in cancer progression. J Biomed Sci. 2016;23:53.

86. Siravegna G, Mussolin B, Venesio T, Marsoni S, Seoane J, Dive C, et al. How liquid biopsies can change clinical practice in oncology. Ann Oncol. 2019;30:1580–90.

87. Pidsley R, Zotenko E, Peters TJ, Lawrence MG, Risbridger GP, Molloy P, et al. Critical evaluation of the Illumina MethylationEPIC BeadChip microarray for whole-genome DNA methylation profiling. Genome Biol. 2016;17:208.

88. McCartney DL, Walker RM, Morris SW, McIntosh AM, Porteous DJ, Evans KL. Identification of polymorphic and off-target probe binding sites on the Illumina Infinium MethylationEPIC BeadChip. Genom Data. 2016;9:22–4.

89. Chen Y-A, Lemire M, Choufani S, Butcher DT, Grafodatskaya D, Zanke BW, et al. Discovery of cross-reactive probes and polymorphic CpGs in the Illumina Infinium HumanMethylation450 microarray. Epigenetics. Informa UK Limited; 2013;8:203–9.

90. Benton MC, Johnstone A, Eccles D, Harmon B, Hayes MT, Lea RA, et al. An analysis of DNA methylation in human adipose tissue reveals differential modification of obesity genes before and after gastric bypass and weight loss. Genome Biol. Springer Science and Business Media LLC; 2015;16:8.

91. Jaffe AE, Murakami P, Lee H, Leek JT, Fallin MD, Feinberg AP, et al. Bump hunting to identify differentially methylated regions in epigenetic epidemiology studies. Int J Epidemiol. Oxford University Press (OUP); 2012;41:200–9.

92. Pedregosa F, Varoquaux G, Gramfort A, Michel V, Thirion B, Grisel O, et al. Scikit-learn: Machine Learning in Python. J Mach Learn Res. JMLR.org; 2011;12:2825–30.

93. Pidsley R Y Wong CC, Volta M, Lunnon K, Mill J, Schalkwyk LC. A data-driven approach to preprocessing Illumina 450K methylation array data. BMC Genomics. 2013;14:293.

94. Lippert C, Casale FP, Rakitsch B, Stegle O. LIMIX: genetic analysis of multiple traits [Internet]. bioRxiv. 2014 [cited 2017 Dec 14]. p. 003905. Available from: http://www.biorxiv.org/content/early/2014/05/22/003905

