## Supplementary File 1 for "DNA methylation fingerprint for the diagnosis and monitoring of hepatocellular carcinoma from tissue and liquid biopsies": BinaryMergedGridSearch.py

```
import sys

sys.path.extend([
    "/home/dockeruser/jupyter_notebooks/LiquidBiopsy",
    "/home/dockeruser/jupyter_notebooks/LiquidBiopsy/LiquidBiopsy",
])
print(f"Python {sys.version} on {sys.platform}")

import numpy as np
import pandas as pd
import seaborn as sns
import matplotlib.pyplot as plt
from LiquidBiopsy import LOG
from sklearn.svm import LinearSVC
from sklearn.model_selection import GridSearchCV
from MLiquidBiopsy import MLiquidBiopsy, MLiquidBiopsyPlot


PLOT_PREFIX = "Binary_Merged"

PAL_LABEL = pd.Series(sns.color_palette("Set1", n_colors=2).as_hex(), index=["Cirrhosis", "HCC"])

LABELS_TRAIN = [
    "Primary Tumor - Liver",
    "cfDNA HCC and cirrhosis",
    "Cirrhosis + EtOH",
    "Cirrhosis + HCV",
    "cfDNA cirrhosis only",
]

LABELS_TEST = [
    "Cirrhosis + HBV",
    "Cirrhosis + Other",
    "Cirrhosis + AATD",
    "HCC + EtOH",
    "HCC + HCV",
]


def tissue_multi_class(stype):
    if stype in [
        "Primary Tumor - Liver",
        "cfDNA HCC and cirrhosis",
        "HCC + EtOH",
        "HCC + HCV",
    ]:
        return 1

    elif stype in [
        "Cirrhosis + EtOH",
        "Cirrhosis + HBV",
        "Cirrhosis + HCV",
        "Cirrhosis + AATD",
        "Cirrhosis + Other",
        "cfDNA cirrhosis only",
    ]:
        return 0

    else:
        return np.nan


def dim_red(df, name):
    methy_pcs, methy_umap, methy_vexp = MLiquidBiopsy.dimension_reduction(df, ss_all)

    methy_pcs.to_csv(f"reports/{PLOT_PREFIX}_{name}_PCs.csv")
    methy_umap.to_csv(f"reports/{PLOT_PREFIX}_{name}_UMAP.csv")

    MLiquidBiopsyPlot.dimension_reduction_plot(
        "PC1", "PC2", "label", methy_pcs, vexp=methy_vexp, palette=pal_label
    )
    plt.savefig(f"reports/{PLOT_PREFIX}_{name}_PCA.pdf", bbox_inches="tight", dpi=600)
    plt.close("all")

    MLiquidBiopsyPlot.dimension_reduction_plot(
        "Dim1", "Dim2", "label", methy_umap, method="UMAP", palette=pal_label
    )
    plt.savefig(f"reports/{PLOT_PREFIX}_{name}_UMAP.pdf", bbox_inches="tight", dpi=600)
    plt.close("all")


def matrices(df, ss):
    Y_train = ss.query(f"label in @LABELS_TRAIN")["label"].apply(tissue_multi_class)
    X_train = df[Y_train.index].copy().T

    Y_test = ss.query(f"label in @LABELS_TEST")["label"].apply(tissue_multi_class)
    X_test = df[Y_test.index].copy().T

    return X_train, Y_train, X_test, Y_test


def matrices_train(df, ss):
    X_train, Y_train, X_test, Y_test = matrices(df, ss)

    clf = MLiquidBiopsy.get_classifier(ctype="LinearSVC", C=1).fit(X_train, Y_train)

    clf_eval_train = MLiquidBiopsy.eval_pred(Y_train, clf.predict(X_train), as_str=False)
    clf_eval_train["etype"] = "train"

    clf_eval_test = MLiquidBiopsy.eval_pred(Y_test, clf.predict(X_test), as_str=False)
    clf_eval_test["etype"] = "test"

    return clf, clf_eval_train, clf_eval_test


if __name__ == "__main__":
    """
    Imports
    """
    # Samplesheet
    ss_all = pd.read_csv(
        f"~/datasets/DNAMethylationPipeline_samplesheet.csv.gz", index_col=0
    )
    ss_all = ss_all[ss_all["label"].isin(LABELS_TRAIN + LABELS_TEST)]
    pal_label = ss_all.groupby("label")["palette"].first()

    # Cols manifest
    manifest_cols = [
        "CHR_hg38", "Start_hg38", "End_hg38",
        "Strand_hg38", "SourceSeq", "UCSC_CpG_Islands_Name",
        "Relation_to_UCSC_CpG_Island", "UCSC_RefGene_Name", "UCSC_RefGene_Group"
    ]

    # Probe annotation information
    pannot = MLiquidBiopsy.read_probe_annot()

    # Methylation
    methy_b = pd.read_csv(f"~/datasets/DNAMethylationPipeline_MergedSubset_methy_b.csv")
    methy_b = methy_b[[c for c in methy_b if c in ss_all.index]]
    methy_b = methy_b.drop(columns=["8784233035_R03C02"])

    ss_all = ss_all[ss_all.index.isin(methy_b.columns)]

    # Drop NaNs
    methy_b = methy_b.dropna()

    # Drop probes not in hg38 lift annotation
    methy_b = methy_b.loc[methy_b.index.isin(pannot.index)]

    # Drop sex chromosome
    psex = MLiquidBiopsy.probes_sex_chr()
    methy_b = methy_b.drop(index=psex, errors="ignore")

    # Grid Search
    Y_train = ss_all["label"].apply(tissue_multi_class)
    X_train = methy_b[Y_train.index].copy().T

    ml = GridSearchCV(
        estimator=LinearSVC(max_iter=1e8),
        param_grid=dict(
            tol=[1e-6],
            penalty=["l1"],
            dual=[False],
            C=[0.1, 0.25, 0.5, 0.75, 1, 2, 5],
            fit_intercept=[True],
            class_weight=[None],
        ),
        verbose=1,
    ).fit(X_train, Y_train)
    LOG.info(ml.best_estimator_)
```
