## Supplementary File 1 for "DNA methylation fingerprint for the diagnosis and monitoring of hepatocellular carcinoma from tissue and liquid biopsies": BinaryMergedTraining.py

```
import numpy as np
import pandas as pd
import seaborn as sns
import matplotlib.pyplot as plt
from LiquidBiopsy import LOG
from crispy import CrispyPlot
from crispy.GIPlot import GIPlot
from sklearn.svm import LinearSVC
from matplotlib_venn import venn3, venn3_circles
from sklearn.model_selection import StratifiedKFold, GridSearchCV, LeaveOneOut
from sklearn.metrics import precision_score, recall_score
from MLiquidBiopsy import MLiquidBiopsy, MLiquidBiopsyPlot
from sklearn.feature_selection import SequentialFeatureSelector


PLOT_PREFIX = "Binary_Merged"

PAL_LABEL = pd.Series(sns.color_palette("Set1", n_colors=2).as_hex(), index=["Cirrhosis", "HCC"])

LABELS_TRAIN = [
    "Primary Tumor - Liver",
    "cfDNA HCC and cirrhosis",
    "Cirrhosis + EtOH",
    "Cirrhosis + HCV",
    "cfDNA cirrhosis only",
]

LABELS_TEST = [
    "Cirrhosis + HBV",
    "Cirrhosis + Other",
    "Cirrhosis + AATD",
    "HCC + EtOH",
    "HCC + HCV",
]


def tissue_multi_class(stype):
    if stype in [
        "Primary Tumor - Liver",
        "cfDNA HCC and cirrhosis",
        "HCC + EtOH",
        "HCC + HCV",
    ]:
        return 1

    elif stype in [
        "Cirrhosis + EtOH",
        "Cirrhosis + HBV",
        "Cirrhosis + HCV",
        "Cirrhosis + AATD",
        "Cirrhosis + Other",
        "cfDNA cirrhosis only",
    ]:
        return 0

    else:
        return np.nan


def dim_red(df, name):
    methy_pcs, methy_umap, methy_vexp = MLiquidBiopsy.dimension_reduction(df, ss_all)

    methy_pcs.to_csv(f"reports/{PLOT_PREFIX}_{name}_PCs.csv")
    methy_umap.to_csv(f"reports/{PLOT_PREFIX}_{name}_UMAP.csv")

    MLiquidBiopsyPlot.dimension_reduction_plot(
        "PC1", "PC2", "label", methy_pcs, vexp=methy_vexp, palette=pal_label
    )
    plt.savefig(f"reports/{PLOT_PREFIX}_{name}_PCA.pdf", bbox_inches="tight", dpi=600)
    plt.close("all")

    MLiquidBiopsyPlot.dimension_reduction_plot(
        "Dim1", "Dim2", "label", methy_umap, method="UMAP", palette=pal_label
    )
    plt.savefig(f"reports/{PLOT_PREFIX}_{name}_UMAP.pdf", bbox_inches="tight", dpi=600)
    plt.close("all")


def matrices(df, ss):
    Y_train = ss.query(f"label in @LABELS_TRAIN")["label"].apply(tissue_multi_class)
    X_train = df[Y_train.index].copy().T

    Y_test = ss.query(f"label in @LABELS_TEST")["label"].apply(tissue_multi_class)
    X_test = df[Y_test.index].copy().T

    return X_train, Y_train, X_test, Y_test


def matrices_train(df, ss, method="LinearSVC"):
    X_train, Y_train, X_test, Y_test = matrices(df, ss)

    clf = MLiquidBiopsy.get_classifier(ctype=method, C=0.5).fit(X_train, Y_train)

    clf_eval_train = MLiquidBiopsy.eval_pred(Y_train, clf.predict(X_train), as_str=False)
    clf_eval_train["etype"] = "train"

    clf_eval_test = MLiquidBiopsy.eval_pred(Y_test, clf.predict(X_test), as_str=False)
    clf_eval_test["etype"] = "test"

    return clf, clf_eval_train, clf_eval_test


if __name__ == "__main__":
    """
    Imports
    """
    # Samplesheet
    ss_all = pd.read_csv(
        f"~/datasets/DNAMethylationPipeline_samplesheet.csv.gz", index_col=0
    )
    ss_all = ss_all[ss_all["label"].isin(LABELS_TRAIN + LABELS_TEST)]
    pal_label = ss_all.groupby("label")["palette"].first()

    # Cols manifest
    manifest_cols = [
        "CHR_hg38", "Start_hg38", "End_hg38",
        "Strand_hg38", "SourceSeq", "UCSC_CpG_Islands_Name",
        "Relation_to_UCSC_CpG_Island", "UCSC_RefGene_Name", "UCSC_RefGene_Group"
    ]

    # Probe annotation information
    pannot = MLiquidBiopsy.read_probe_annot()

    # Methylation
    methy_b = pd.read_csv(f"~/datasets/DNAMethylationPipeline_MergedSubset_methy_b.csv")
    methy_b = methy_b[[c for c in methy_b if c in ss_all.index]]
    methy_b = methy_b.drop(columns=["8784233035_R03C02"])

    ss_all = ss_all[ss_all.index.isin(methy_b.columns)]

    # Drop NaNs
    methy_b = methy_b.dropna()

    # Drop probes not in hg38 lift annotation
    methy_b = methy_b.loc[methy_b.index.isin(pannot.index)]

    # Drop sex chromosome
    psex = MLiquidBiopsy.probes_sex_chr()
    methy_b = methy_b.drop(index=psex, errors="ignore")

    # Dimension reduction
    dim_red(methy_b.T, "methy")

    # Global methylation
    methy_b_mean = methy_b.T.mean(1)
    methy_b_mean_corr = methy_b.T.corrwith(methy_b_mean)

    _, ax = plt.subplots(1, 1, figsize=(2, 1.5), sharex="all", sharey="all")

    sns.distplot(
        methy_b_mean_corr,
        hist=True,
        kde=False,
        bins=50,
        hist_kws=dict(linewidth=0, alpha=1),
        color=CrispyPlot.PAL_DTRACE[2],
        ax=ax,
    )

    ax.grid(True, ls="-", lw=0.1, alpha=1.0, zorder=0)
    ax.set_xlabel("CpG correlation with global methylation")
    ax.set_ylabel("Number of CpGs")
    plt.savefig(
        f"reports/{PLOT_PREFIX}_global_methylation_corr_distplot.pdf", bbox_inches="tight", dpi=600
    )
    plt.close("all")

    c_thres_eval = []
    for c_thres in np.arange(0.1, 0.75, 0.05):
        methy_b_c_thres = methy_b.loc[methy_b_mean_corr[methy_b_mean_corr.abs() < c_thres].index]

        _, eval_train, eval_test = matrices_train(methy_b_c_thres, ss_all, method="Logistic")

        eval_train["c_thres"] = c_thres
        eval_train["n_features"] = methy_b_c_thres.shape[0]
        c_thres_eval.append(eval_train)

        eval_test["c_thres"] = c_thres
        eval_test["n_features"] = methy_b_c_thres.shape[0]
        c_thres_eval.append(eval_test)
    c_thres_eval = pd.DataFrame(c_thres_eval)
    c_thres_eval.to_csv(f"reports/{PLOT_PREFIX}_global_methylation_corr_thres_eval_Logistic.csv", index=False)

    pal = dict(train=CrispyPlot.PAL_DTRACE[1], test=CrispyPlot.PAL_DTRACE[2])

    plot_df = c_thres_eval.copy()
    plot_df["name"] = [f"{n:.0f} (R < {r:.2f})" for n, r in plot_df[["n_features", "c_thres"]].values]

    _, axs = plt.subplots(2, 1, figsize=(4, 3), sharey="all", sharex="all")

    for i, y_var in enumerate(["recall", "precision"]):
        sns.barplot(
            x="name",
            y=y_var,
            hue="etype",
            data=plot_df,
            hue_order=["train", "test"],
            lw=0,
            palette=pal,
            saturation=1,
            ax=axs[i],
        )

        axs[i].grid(True, ls="-", lw=0.1, alpha=1.0, zorder=0)

        axs[i].set_xlabel("Number of Features" if i == 1 else "")
        axs[i].set_ylabel(y_var)
        axs[i].set_title("Global methylation filtering" if i == 0 else "")

        axs[i].legend(
            frameon=False,
            title="",
            prop={"size": 5},
        )

    plt.xticks(rotation=45, ha='right')
    plt.subplots_adjust(hspace=0.1, wspace=0.1)
    plt.savefig(
        f"reports/{PLOT_PREFIX}_global_methylation_corr_thres_eval_Logistic.pdf", bbox_inches="tight", dpi=600
    )
    plt.close("all")

    methy_b_ = methy_b.loc[methy_b_mean_corr[methy_b_mean_corr.abs() < 0.35].index]

    # Cluster methylation
    n_thres_eval = []
    for n_thres in np.arange(1, 15):
        n_thres_methy_b_cluster, _ = MLiquidBiopsy.probes_cluster(methy_b_, nmin=n_thres)

        _, eval_train, eval_test = matrices_train(n_thres_methy_b_cluster, ss_all, method="Logistic")

        eval_train["n_min"] = n_thres
        eval_train["n_features"] = n_thres_methy_b_cluster.shape[0]
        n_thres_eval.append(eval_train)

        eval_test["n_min"] = n_thres
        eval_test["n_features"] = n_thres_methy_b_cluster.shape[0]
        n_thres_eval.append(eval_test)

    n_thres_eval = pd.DataFrame(n_thres_eval)
    n_thres_eval.to_csv(f"reports/{PLOT_PREFIX}_n_cluster_thres_eval_Logistic.csv", index=False)

    pal = dict(train=CrispyPlot.PAL_DTRACE[1], test=CrispyPlot.PAL_DTRACE[2])

    plot_df = n_thres_eval.copy()
    plot_df["name"] = [f"{n:.0f} (n CpGs >= {r:.0f})" for n, r in plot_df[["n_features", "n_min"]].values]

    _, axs = plt.subplots(2, 1, figsize=(4.5, 3), sharey="all", sharex="all")

    for i, y_var in enumerate(["recall", "precision"]):
        sns.barplot(
            x="name",
            y=y_var,
            hue="etype",
            data=plot_df,
            hue_order=["train", "test"],
            lw=0,
            palette=pal,
            saturation=1,
            ax=axs[i],
        )

        axs[i].grid(True, ls="-", lw=0.1, alpha=1.0, zorder=0)

        axs[i].set_xlabel("Min number of CpGs per cluster" if i == 1 else "")
        axs[i].set_ylabel(y_var)
        axs[i].set_title("CpG clusters" if i == 0 else "")

        axs[i].legend(
            frameon=False,
            title="",
            prop={"size": 5},
        )

    plt.xticks(rotation=45, ha='right')
    plt.subplots_adjust(hspace=0.1, wspace=0.1)
    plt.savefig(
        f"reports/{PLOT_PREFIX}_n_cluster_thres_eval_Logistic.pdf", bbox_inches="tight", dpi=600
    )
    plt.close("all")

    methy_b_cluster, cluster_map = MLiquidBiopsy.probes_cluster(methy_b_, nmin=5)
    cluster_map_cpg = cluster_map.reset_index().groupby("cluster")["IlmnID"].agg(set)

    """
    Classifier
    """
    y_features, y_eval, y_pred, y_nfeatures = [], [], [], []

    for r_idx in range(100):
        # Undersample
        X = pd.concat(
            [
                methy_b_cluster[ss_all.query("label == 'Primary Tumor - Liver'").sample(22).index].T,
                methy_b_cluster[ss_all.query("label == 'cfDNA HCC and cirrhosis'").sample(22).index].T,
                methy_b_cluster[ss_all.query("label == 'Cirrhosis + HCV'").sample(11).index].T,
                methy_b_cluster[ss_all.query("label == 'Cirrhosis + EtOH'").sample(11).index].T,
                methy_b_cluster[ss_all.query("label == 'cfDNA cirrhosis only'").sample(22).index].T,
            ]
        )
        Y = ss_all.loc[X.index, "label"].apply(tissue_multi_class)

        # Objects to store results
        Y_pred = {}

        for cv_idx, (idx_train, idx_test) in enumerate(LeaveOneOut().split(X, Y)):
            # ML CV matrices
            cv_x_train, cv_y_train = X.iloc[idx_train], Y.iloc[idx_train]
            cv_x_test, cv_y_test = X.iloc[idx_test], Y.iloc[idx_test]

            # ANOVA
            dtanova = MLiquidBiopsy.anova(cv_x_train, cv_y_train)
            ftype_cpgs = set(dtanova.query(f"fdr < 0.01").index)
            y_nfeatures.append(ftype_cpgs)
            LOG.info(f"{r_idx} {cv_idx}: #(CpGs)={len(ftype_cpgs)}")

            if len(ftype_cpgs) < 10:
                continue

            cv_x_train = cv_x_train.reindex(index=cv_y_train.index, columns=ftype_cpgs)
            cv_x_test = cv_x_test.reindex(index=cv_y_test.index, columns=ftype_cpgs)

            # Classifier
            clf = MLiquidBiopsy.get_classifier(ctype="LinearSVC", C=1).fit(cv_x_train, cv_y_train)

            # Predict
            Y_pred[cv_y_test.index[0]] = clf.predict(cv_x_test)[0]

            # Coefficients
            clf_coefs = pd.Series(clf.coef_[0], cv_x_train.columns)
            clf_coefs = clf_coefs[clf_coefs != 0]

        Y_pred = pd.Series(Y_pred)[Y.index]

        clf_eval_test = MLiquidBiopsy.eval_pred(Y, Y_pred, as_str=False)
        print(clf_eval_test)

        # Eval
        clf_coefs = pd.Series((clf.coef_ != 0).sum(1), index=clf.classes_)
        clf_eval_train = MLiquidBiopsy.eval_pred(cv_y_train, clf.predict(cv_x_train), as_str=False)
        clf_eval_test = MLiquidBiopsy.eval_pred(cv_y_test, clf.predict(cv_x_test), as_str=False)

        LOG.info(f"{r_idx} {cv_idx}: Non-zero features={clf_coefs}")
        LOG.info(f"{r_idx} {cv_idx}: Train={'; '.join([f'{m}={s:.2f}' for m, s in clf_eval_train.items()])}")
        LOG.info(f"{r_idx} {cv_idx}: Test={'; '.join([f'{m}={s:.2f}' for m, s in clf_eval_test.items()])}")

        # Store results
        clf_features = pd.DataFrame(clf.coef_, index=clf.classes_, columns=ftype_cpgs).T
        clf_features = (clf_features.unstack().replace(0, np.nan).dropna().reset_index())
        clf_features.columns = ["ftype", "feature", "coef"]
        y_features.append(clf_features)

        clf_eval_test["etype"] = "test"
        y_eval.append(clf_eval_test)

        clf_eval_train["etype"] = "train"
        y_eval.append(clf_eval_train)

        cv_y_test_pred = pd.Series(clf.predict(cv_x_train), index=cv_x_train.index)
        y_pred.append(cv_y_test_pred)

    y_features = pd.concat(y_features, ignore_index=True)
    y_features.to_csv(f"reports/PredMultiClass_Merged_y_features.csv", index=False)

    y_eval = pd.DataFrame(y_eval)
    y_eval.to_csv(f"reports/PredMultiClass_Merged_y_eval.csv", index=False)
```
