## Supplementary File 1 for "DNA methylation fingerprint for the diagnosis and monitoring of hepatocellular carcinoma from tissue and liquid biopsies": BinaryTrainingPipeline.py

```
import numpy as np
import pandas as pd
import seaborn as sns
import matplotlib.pyplot as plt
from LiquidBiopsy import LOG
from crispy import CrispyPlot
from crispy.GIPlot import GIPlot
from upsetplot import from_contents
from crispy.LMModels import LMModels
from upsetplot import plot as upsetplot
from sklearn.utils.extmath import softmax
from sklearn.ensemble import VotingClassifier
from BinaryTrainingPlot import BinaryTrainingPlot
from sklearn.metrics import precision_score, recall_score
from MLiquidBiopsy import MLiquidBiopsy, MLiquidBiopsyPlot
from sklearn.feature_selection import SequentialFeatureSelector
from sklearn.model_selection import LeaveOneOut
from sklearn.svm import LinearSVC
from sklearn.linear_model import (
    PassiveAggressiveClassifier,
    LogisticRegressionCV,
    RidgeClassifierCV,
    Perceptron,
    RidgeClassifier,
)


class RidgeClassifierCVwithProba(RidgeClassifierCV):
    def predict_proba(self, X):
        d = self.decision_function(X)
        d_2d = np.c_[-d, d]
        return softmax(d_2d)


class PerceptronwithProba(Perceptron):
    def predict_proba(self, X):
        d = self.decision_function(X)
        d_2d = np.c_[-d, d]
        return softmax(d_2d)


class PassiveAggressiveClassifierwithProba(PassiveAggressiveClassifier):
    def predict_proba(self, X):
        d = self.decision_function(X)
        d_2d = np.c_[-d, d]
        return softmax(d_2d)


PAL_LABEL = pd.Series(
    sns.color_palette("Set1", n_colors=2).as_hex(), index=["Cirrhosis", "HCC"]
)

LABELS = [
    "Primary Tumor - Liver",
    "HCC + EtOH",
    "HCC + HCV",
    "cfDNA HCC and cirrhosis",
    "cfDNA cirrhosis only",
    "Cirrhosis + EtOH",
    "Cirrhosis + HCV",
    "Cirrhosis + HBV",
    "Cirrhosis + Other",
    "Cirrhosis + AATD",
    "AATD",
    "Liver obese",
    "Solid Tissue Normal - Liver",
    "Liver normal",
    "cfDNA",
    "cfDNA healthy pooled",
    "cfDNA healthy",
]

LABELS_CFDNA = [
    "cfDNA HCC and cirrhosis",
    "cfDNA cirrhosis only",
    "cfDNA healthy",
    "cfDNA healthy pooled",
    # "cfDNA",
]


def build_matrices(x_train, y_train, ss, labels_hepatocc=None, labels_cirrhosis=None):
    if labels_hepatocc is None:
        labels_hepatocc = [
            "Primary Tumor - Liver",
        ]

    if labels_cirrhosis is None:
        labels_cirrhosis = [
            "Cirrhosis + HBV",
            "Cirrhosis + AATD",
            "Cirrhosis + HCV",
            "Cirrhosis + EtOH",
        ]

    ss = ss.query(f"dataset_id != 'GSE60753' | PC2 < 200")

    nmin_samples = min(
        ss.query("label == 'cfDNA HCC and cirrhosis'").sample(frac=1.0).shape[0],
        ss.query("label == 'cfDNA cirrhosis only'").sample(frac=1.0).shape[0],
    )

    x_train_ = pd.concat(
        [
            x_train.loc[
                ss.query("label in @labels_hepatocc").sample(nmin_samples).index
            ],
            x_train.loc[
                ss.query("label == 'cfDNA HCC and cirrhosis'")
                .sample(nmin_samples)
                .index
            ],
            x_train.loc[
                ss.query("label in @labels_cirrhosis").sample(nmin_samples).index
            ],
            x_train.loc[
                ss.query("label == 'cfDNA cirrhosis only'").sample(nmin_samples).index
            ],
        ]
    ).copy()

    y_train_ = y_train[x_train_.index].copy()

    return x_train_, y_train_


def train_coefs(x, y, ss, fdr_thres=0.01):
    # Undersample
    cv_x_train_under, cv_y_train_under = build_matrices(x, y, ss.loc[y.index])

    # ANOVA
    extra_covs = ss_all[["GlobalMethylation", "SamplePurityPC5"]]

    dtanova = MLiquidBiopsy.anova_cov(
        cv_x_train_under,
        cv_y_train_under,
        ss,
        dataset_as_cov=False,
        extra_covs=extra_covs,
    )
    ftype_cpgs = set(
        dtanova.query(f"fdr < {fdr_thres} & ANOVA_fdr < {fdr_thres}").index
    )
    LOG.info(f"#(CpGs)={len(ftype_cpgs)}")

    cv_x_train_under = cv_x_train_under.reindex(
        index=cv_y_train_under.index, columns=ftype_cpgs
    )

    # Classifier
    clf_under = LinearSVC(
        penalty="l1",
        C=1.5,
        dual=False,
        max_iter=1e5,
    ).fit(cv_x_train_under, cv_y_train_under)

    # Coefficients
    clf_coefs = pd.Series(clf_under.coef_[0], cv_x_train_under.columns)
    clf_coefs = clf_coefs[clf_coefs != 0]
    LOG.info(f"Non-zero CpGs={len(clf_coefs)}")

    return clf_coefs, len(ftype_cpgs)


def run_ml(X, Y, cv, ss, fdr_thres=0.01):
    pred = {f: [] for f in features}
    prob = {f: [] for f in features}

    coefs, anovas = [], []

    X_std = pd.concat(
        [
            LMModels.transform_matrix(
                X.loc[ss.query(f"label not in @LABELS_CFDNA").index], add_nan_mask=False
            ),
            LMModels.transform_matrix(
                X.loc[ss.query(f"label in @LABELS_CFDNA").index], add_nan_mask=False
            ),
        ]
    ).loc[Y.index]
    ss = ss.loc[Y.index]

    cv = list(enumerate(cv.split(X_std, Y)))
    # cv = [(i, (train, test)) for i, (train, test) in cv if ss.loc[Y.iloc[test].index[0], "label"] in LABELS_CFDNA]
    for idx_cv, (idx_train, idx_test) in cv:
        # ML CV matrices
        cv_x_train, cv_y_train, cv_ss_train = (
            X_std.iloc[idx_train],
            Y.iloc[idx_train],
            ss.iloc[idx_train],
        )
        cv_x_test = X_std.iloc[idx_test]
        LOG.info(f"CV {idx_cv}: {Y.iloc[idx_test].index[0]}")

        # Classifiers
        for ftype in features:
            if ftype in MLiquidBiopsy.TOP_CPGS:
                # Benchmark sets
                ftype_cpgs = MLiquidBiopsy.TOP_CPGS[ftype].intersection(methy_b.index)
                ftype_x_train = (
                    methy_b.reindex(index=ftype_cpgs, columns=cv_y_train.index)
                    .dropna()
                    .T.copy()
                )
                ftype_x_test = (
                    methy_b.reindex(index=ftype_cpgs, columns=cv_x_test.index)
                    .dropna()
                    .T.copy()
                )

            else:
                # DMR signature features
                clf_coefs, t_anovas = train_coefs(
                    cv_x_train, cv_y_train, cv_ss_train, fdr_thres
                )
                coefs.append(clf_coefs)
                anovas.append(t_anovas)

                # Subset
                ftype_x_train = cv_x_train.reindex(
                    index=cv_y_train.index, columns=clf_coefs.index
                )
                ftype_x_test = cv_x_test.reindex(
                    index=cv_x_test.index, columns=clf_coefs.index
                )

            # All - Classifier
            clfs = [
                ("Ridge", RidgeClassifierCVwithProba(alphas=[0.1, 0.5, 1, 2, 4, 10])),
                ("Logistic", LogisticRegressionCV(max_iter=1e6, tol=1e-6)),
                # ("Gaussian", GaussianNB()),
                # ("Perceptron", PerceptronwithProba(alpha=1, max_iter=1e6, tol=1e-6)),
                # ("PassiveAggressive", PassiveAggressiveClassifierwithProba(C=1., max_iter=1e6, tol=1e-6)),
                # ("SVC", SVC(C=1, kernel="linear", probability=True)),
            ]
            clf = VotingClassifier(estimators=clfs, voting="soft").fit(
                ftype_x_train, cv_y_train
            )

            pred[ftype].append(
                pd.Series(clf.predict(ftype_x_test), index=cv_x_test.index)
            )
            prob[ftype].append(
                pd.Series(clf.predict_proba(ftype_x_test)[:, 1], index=cv_x_test.index)
            )

    pred = pd.DataFrame({k: pd.concat(v) for k, v in pred.items()})
    prob = pd.DataFrame({k: pd.concat(v) for k, v in prob.items()})

    coefs = pd.concat(coefs, axis=1)

    return pred, prob, coefs, anovas


if __name__ == "__main__":
    """
    Imports
    """
    # Samplesheet
    methy_b, ss_all, pal_label, pal_type = MLiquidBiopsy.import_dataset(
        f"~/datasets/DNAMethylationPipeline_MergedSubset_methy_b.csv",
        subset_labels=LABELS,
    )
    BinaryTrainingPlot.plot_number_samples(ss_all, pal_type)
    BinaryTrainingPlot.plot_number_samples_per_dataset(ss_all, name="methy")

    # Probe annotation information
    pannot = MLiquidBiopsy.read_probe_annot()

    # Dataset palette
    pal_dataset = ss_all.groupby("dataset_id")["dataset_color"].first()

    # Feature palette
    features = ["DMR signature"] + list(MLiquidBiopsy.TOP_CPGS)
    markers = ["o"] + ["x"] * len(list(MLiquidBiopsy.TOP_CPGS))
    linestyles = ["-"] + ["--"] * len(list(MLiquidBiopsy.TOP_CPGS))
    features_pal = BinaryTrainingPlot.palette_feature()

    """
    Feature reduction
    """
    N_FEATURES_REDUCTION = dict()
    N_FEATURES_REDUCTION["Overlap 450K & EPIC"] = [575130]
    N_FEATURES_REDUCTION["FunNorm processing"] = [452567]
    N_FEATURES_REDUCTION["Drop loci with SNPs"] = [436469]
    N_FEATURES_REDUCTION["Drop cross-reactive loci"] = [402433]

    # Drop NaNs
    methy_b = methy_b.dropna()
    N_FEATURES_REDUCTION["Drop NaNs"] = [methy_b.shape[0]]
    # methy_b.to_csv(f"tables/LOO_methylation_matrix.csv.gz", compression="gzip")

    # Dimension reduction
    pcs, dims, vexp = BinaryTrainingPlot.dim_red(
        methy_b.T, "methy", ss_all, pal_label, n_components=10
    )
    BinaryTrainingPlot.pairplot_pca(pcs, vexp, ss_all)
    plt.savefig(
        f"reports/LOO_methy_PCA_pairplot.pdf",
        bbox_inches="tight",
        dpi=600,
    )
    plt.close("all")

    BinaryTrainingPlot.plot_pca(
        pcs,
        [("PC1", "PC2")],
        ss_all,
        vexp,
        pal_type,
    )

    specific_labels = [
        "Primary Tumor - Liver",
        "HCC + EtOH",
        "HCC + HCV",
        "cfDNA healthy",
        "cfDNA healthy pooled",
        "Cirrhosis + HBV",
        "Cirrhosis + AATD",
        "Cirrhosis + HCV",
        "Cirrhosis + EtOH",
    ]

    df = pcs.query(f"label in @specific_labels")
    df = df.query(f"dataset_id != 'GSE60753' | PC2 < 200")

    MLiquidBiopsyPlot.dimension_reduction_plot(
        "PC1", "PC2", "dataset_id", df, vexp=vexp, palette=pal_dataset
    )
    plt.savefig(
        f"reports/LOO_PCA_specific_labels_dataset.pdf", bbox_inches="tight", dpi=600
    )
    plt.close("all")

    MLiquidBiopsyPlot.dimension_reduction_plot(
        "PC1", "PC2", "label", df, vexp=vexp, palette=pal_label
    )
    plt.savefig(
        f"reports/LOO_PCA_specific_labels_label.pdf", bbox_inches="tight", dpi=600
    )
    plt.close("all")

    MLiquidBiopsyPlot.dimension_reduction_plot(
        "Dim1", "Dim2", "dataset_id", dims, method="UMAP", palette=pal_dataset
    )
    plt.savefig(f"reports/LOO_methy_UMAP_dataset.pdf", bbox_inches="tight", dpi=600)
    plt.close("all")

    # Drop probes not in hg38 lift annotation
    methy_b_ = methy_b.loc[methy_b.index.isin(pannot.index)].copy()
    N_FEATURES_REDUCTION["hg38 lift"] = [methy_b_.shape[0]]

    # Drop sex chromosome
    psex = MLiquidBiopsy.probes_sex_chr()
    methy_b_ = methy_b_.drop(index=psex, errors="ignore")
    N_FEATURES_REDUCTION["Drop chrX & chrY"] = [methy_b_.shape[0]]

    # Global methylation
    methy_b_mean = methy_b_.T.mean(1)
    methy_b_mean_corr = methy_b_.T.corrwith(methy_b_mean)
    BinaryTrainingPlot.plot_global_corr_hist(methy_b_mean_corr)

    ss_all["GlobalMethylation"] = methy_b_mean[ss_all.index].values

    for cpg in ["cg16714888"]:
        plot_df = pd.concat(
            [
                methy_b.loc[cpg],
                methy_b_mean.rename("mean"),
                ss_all,
            ],
            axis=1,
        )

        GIPlot.gi_regression(cpg, "mean", plot_df, hue="Type", palette=pal_type)
        plt.savefig(
            f"reports/LOO_global_methylation_corrplot_{cpg}.pdf",
            bbox_inches="tight",
            dpi=600,
        )
        plt.close("all")

    # Tumour purity
    tumour_purity = ss_all["purity_CPE"].dropna()
    methy_b_mean_corr_purity = methy_b_[tumour_purity.index].T.corrwith(tumour_purity)
    methy_b_mean_corr_pc5 = methy_b_[pcs.index].T.corrwith(pcs["PC5"])

    ss_all["SamplePurityPC5"] = pcs.loc[ss_all.index, "PC5"].values

    BinaryTrainingPlot.purity_corr_pcs_barplot(pcs, vexp)
    BinaryTrainingPlot.purity_regression(
        "PC5", pcs, vexp, methy_b_mean_corr_pc5, pal_type
    )

    # Cluster methylation
    methy_b_cluster, cluster_map = MLiquidBiopsy.probes_cluster(methy_b_, nmin=3)
    cluster_map_cpg = cluster_map.reset_index().groupby("cluster")["IlmnID"].agg(set)
    N_FEATURES_REDUCTION["CpG clusters"] = [methy_b_cluster.shape[0]]

    BinaryTrainingPlot.plot_clusters_length(pannot, methy_b_, methy_b_cluster)
    # methy_b_cluster.round(5).to_csv(
    #     f"tables/LOO_methylation_matrix_cpg_clusters.csv.gz", compression="gzip"
    # )

    dmrs = pannot.loc[cluster_map.index]
    dmrs = pd.concat(
        [
            dmrs.groupby("cluster")["Start_hg38"].min(),
            dmrs.groupby("cluster")["End_hg38"].max(),
            dmrs.groupby("cluster")["CHR_hg38"].first(),
        ],
        axis=1,
    )
    dmrs["name"] = [
        f"DMR_{c.capitalize()}:{s:.0f}-{e:.0f}"
        for c, s, e in dmrs[["CHR_hg38", "Start_hg38", "End_hg38"]].values
    ]

    ss_all.to_excel(f"tables/LOO_samplesheet.xlsx")
    # ss_all = pd.read_excel(f"tables/LOO_samplesheet.xlsx", index_col=0)

    """
    Validation set
    """
    ss_all_validation, methy_b_validation = MLiquidBiopsy.read_test_datasets("all")
    ss_all_validation["Type"] = ss_all_validation["y_true"].replace(
        {0: "Other", 1: "HCC"}
    )
    BinaryTrainingPlot.plot_number_samples_per_dataset(
        ss_all_validation, name="methy_validation"
    )
    # methy_b_validation.to_csv(f"tables/LOO_methylation_validation_matrix.csv.gz", compression="gzip")

    pal_val_dataset = ss_all_validation.groupby("dataset_id")["dataset_color"].first()

    methy_b_validation_cluster = methy_b_validation[
        methy_b_validation.index.isin(methy_b_.index)
    ]
    methy_b_validation_cluster, _ = MLiquidBiopsy.probes_cluster(
        methy_b_validation_cluster, nmin=3
    )
    # methy_b_validation_cluster.round(5).to_csv(
    #     f"tables/LOO_methylation_validation_matrix_cpg_clusters.csv.gz", compression="gzip"
    # )

    Y_validation = ss_all_validation["y_true"].copy()
    X_validation = methy_b_validation_cluster.dropna().T

    pcs_validation, dims_validation, vexp_validation = BinaryTrainingPlot.dim_red(
        methy_b_validation.dropna().T,
        "methy_validation",
        ss_all_validation,
        pal_val_dataset,
        hue_by="dataset_id",
        n_components=10,
    )
    BinaryTrainingPlot.pairplot_pca(pcs_validation, vexp_validation, ss_all_validation)
    plt.savefig(
        f"reports/LOO_methy_validation_PCA_pairplot.pdf",
        bbox_inches="tight",
        dpi=600,
    )
    plt.close("all")

    BinaryTrainingPlot.plot_pca(
        pcs_validation,
        [("PC1", "PC2")],
        ss_all_validation,
        vexp_validation,
        pal_type,
        name="methy_validation",
    )

    MLiquidBiopsyPlot.dimension_reduction_plot(
        "Dim1",
        "Dim2",
        "dataset_id",
        dims_validation,
        method="UMAP",
        palette=pal_val_dataset,
    )
    plt.savefig(
        f"reports/LOO_methy_validation_UMAP_dataset.pdf", bbox_inches="tight", dpi=600
    )
    plt.close("all")

    """
    Matrices
    """
    Y = ss_all["label"].apply(MLiquidBiopsy.sample_class)
    X = methy_b_cluster[Y.index].T

    """
    Samples
    """
    ss_tissue = ss_all[~ss_all["label"].isin(LABELS_CFDNA + ["cfDNA"])]
    ss_cfdna = ss_all[ss_all["label"].isin(LABELS_CFDNA)]

    """
    Models
    """
    # Leave-one-out
    samples = pcs.query("label != 'cfDNA'")

    pred_loo, prob_loo, coefs_loo, anovas_loo = run_ml(
        X.loc[samples.index],
        Y.loc[samples.index],
        LeaveOneOut(),
        samples,
        fdr_thres=0.01,
    )

    pred_loo.to_csv(f"tables/LOO_pred.csv")
    # pred_loo = pd.read_csv(f"tables/LOO_pred.csv", index_col=0)
    prob_loo.to_csv(f"tables/LOO_prob.csv")
    # prob_loo = pd.read_csv(f"tables/LOO_prob.csv", index_col=0)
    coefs_loo.to_csv(f"tables/LOO_coefs.csv")
    # coefs_loo = pd.read_csv(f"tables/LOO_coefs.csv")

    N_FEATURES_REDUCTION["ANOVAs"] = anovas_loo
    N_FEATURES_REDUCTION["SVM features"] = list(coefs_loo.count())

    """
    Biomarkers
    """
    coefs_top = pd.concat(
        [
            coefs_loo.count(1).rename("count"),
            coefs_loo.mean(1).rename("mean"),
        ],
        axis=1,
    ).sort_values("count", ascending=False)
    coefs_top["dmr"] = dmrs.loc[coefs_top.index, "name"].values
    coefs_top.to_excel(f"tables/LOO_coefs_top.xlsx")
    # coefs_top = pd.read_excel(f"tables/LOO_coefs_top.xlsx", index_col=0)

    # Features barplot
    N_FEATURES = BinaryTrainingPlot.plot_feature_reduction_barplot(N_FEATURES_REDUCTION)

    # Feature regression
    plot_df = pd.DataFrame(dict(count=coefs_top["count"], mean=coefs_top["mean"].abs()))
    ax = GIPlot.gi_regression("count", "mean", plot_df, lowess=True)
    ax.ax_joint.axvline(
        coefs_top.head(N_FEATURES)["count"].min(),
        ls="--",
        lw=0.3,
        c=CrispyPlot.PAL_DTRACE[2],
    )
    ax.set_axis_labels("# times selected", "Mean coef (abs)")
    plt.savefig(f"reports/LOO_features_regression.pdf", bbox_inches="tight", dpi=600)
    plt.close("all")

    # Top features
    top_coefs = coefs_top.head(N_FEATURES)
    top_coefs["cpg_sites"] = cluster_map_cpg.loc[top_coefs.index].apply(list).values
    top_coefs = (
        top_coefs.explode("cpg_sites")
        .reset_index()
        .rename(columns=dict(index="cpg_cluster"))
    )
    top_coefs = pd.concat(
        [
            top_coefs,
            pannot.loc[top_coefs["cpg_sites"], MLiquidBiopsy.MANIFEST_COLS].reset_index(
                drop=True
            ),
        ],
        axis=1,
    )

    top_coefs["corr_global_methylation"] = methy_b_mean_corr[
        top_coefs["cpg_sites"]
    ].values
    top_coefs["corr_tumour_purity"] = methy_b_mean_corr_purity[
        top_coefs["cpg_sites"]
    ].values
    top_coefs.to_excel(f"tables/LOO_coefs_top_annotated.xlsx", index=False)
    # top_coefs = pd.read_excel(f"tables/LOO_coefs_top_annotated.xlsx")

    # Feature correlation heatmap
    BinaryTrainingPlot.plot_features_corr_heatmap(
        methy_b_cluster.loc[coefs_top.head(N_FEATURES).index]
        .rename(index=dmrs["name"])
        .T.corr()
    )

    # CpG feature overlap
    features_dict = MLiquidBiopsy.TOP_CPGS.copy()
    features_dict["DMR signature"] = set(top_coefs["cpg_sites"])
    features_dict = from_contents(features_dict)

    upsetplot(features_dict, element_size=16, show_counts=True)
    plt.savefig(
        f"reports/LOO_upsetplot_independent_features.pdf", bbox_inches="tight", dpi=600
    )
    plt.close("all")

    """
    Recursive feature selection
    """
    features_to_test = list(
        coefs_top.query(f"count > {round(Y.shape[0] * 0.05)}").index
    )

    sfs_df = []
    for i in range(30):
        LOG.info(f"SFS {i + 1} iteration")
        x_sfs_train, y_sfs_train = build_matrices(X[features_to_test], Y, samples)

        x_sfs_test = X[features_to_test].drop(index=x_sfs_train.index)
        y_sfs_test = Y.drop(index=y_sfs_train.index)

        for n_features in np.arange(2, N_FEATURES + 1, 2):
            # Select best features
            sfs = SequentialFeatureSelector(
                estimator=LinearSVC(
                    penalty="l1",
                    C=1.5,
                    dual=False,
                    max_iter=1e5,
                ),
                n_features_to_select=n_features,
                direction="forward",
                n_jobs=2,
            ).fit(x_sfs_train, y_sfs_train)

            # Train
            clf = LinearSVC(
                penalty="l1",
                C=1.5,
                dual=False,
                max_iter=1e5,
            ).fit(x_sfs_train.loc[:, sfs.get_support()], y_sfs_train)

            # Selected features
            s_features = list(x_sfs_train.loc[:, sfs.get_support()])

            # Predict
            y_sfs_train_pred = pd.Series(
                clf.predict(x_sfs_train[s_features]), index=y_sfs_train.index
            )
            y_sfs_test_pred = pd.Series(
                clf.predict(x_sfs_test[s_features]), index=y_sfs_test.index
            )
            y_sfs_val_pred = pd.Series(
                clf.predict(X_validation[s_features]), index=Y_validation.index
            )

            # Eval
            sfs_eval_train = MLiquidBiopsy.eval_pred(
                y_sfs_train, y_sfs_train_pred, as_str=False
            )
            sfs_eval_test = MLiquidBiopsy.eval_pred(
                y_sfs_test, y_sfs_test_pred, as_str=False
            )
            sfs_eval_val = MLiquidBiopsy.eval_pred(
                Y_validation, y_sfs_val_pred, as_str=False
            )

            # Store
            sfs_df_train = dict(
                etype="train",
                n_features=n_features,
                features=s_features,
            )
            sfs_df.append({**sfs_df_train, **sfs_eval_train})

            sfs_df_test = dict(
                etype="test",
                n_features=n_features,
                features=s_features,
            )
            sfs_df.append({**sfs_df_test, **sfs_eval_test})

            sfs_df_val = dict(
                etype="validation",
                n_features=n_features,
                features=s_features,
            )
            sfs_df.append({**sfs_df_val, **sfs_eval_val})

    sfs_df = pd.DataFrame(sfs_df)
    sfs_df.to_csv(f"tables/LOO_sfs_eval.csv", index=False)

    plot_df = pd.melt(
        sfs_df,
        id_vars=["etype", "n_features"],
        value_vars=["recall", "precision"],
        var_name="metric",
    ).rename(columns=dict(etype="dataset"))

    pal = dict(
        train=CrispyPlot.PAL_DTRACE[3],
        test=CrispyPlot.PAL_DTRACE[2],
        validation=CrispyPlot.PAL_DTRACE[4],
    )

    fig, ax = plt.subplots(1, 1, figsize=(3, 2.5), sharey="all", sharex="all")

    sns.lineplot(
        x="n_features",
        y="value",
        hue="dataset",
        style="metric",
        data=plot_df,
        palette=pal,
        err_kws=dict(lw=0),
        ax=ax,
    )

    ax.legend(
        frameon=False,
        title="",
        prop={"size": 5},
        loc=4,
    )

    ax.grid(True, ls="-", lw=0.1, alpha=1.0, zorder=0)
    ax.set_xlabel("Number of DMRs")
    ax.set_ylabel("Metric value")

    plt.savefig(f"reports/LOO_sfs_lineplot.pdf", bbox_inches="tight", dpi=600)
    plt.close("all")

    #
    optimal_features = pd.Series(
        [
            f
            for l in sfs_df.query(f"n_features == {N_FEATURES} & etype == 'train'")[
                "features"
            ]
            for f in l
        ]
    ).value_counts()

    plot_df = optimal_features.rename("count").reset_index().head(38)

    order = list(plot_df["index"])

    _, ax = plt.subplots(1, 1, figsize=(2, 6))

    sns.barplot(
        x="count",
        y="index",
        data=plot_df,
        order=order,
        color=CrispyPlot.PAL_DTRACE[2],
        saturation=1,
        lw=0,
        ax=ax,
    )

    for y_var in order:
        n = plot_df.set_index("index").loc[y_var, "count"]
        ax.text(
            n + 0.5,
            order.index(y_var),
            f"{n}",
            fontsize=6,
            va="center",
        )

    ax.grid(axis="x", lw=0.1, color="#e1e1e1", zorder=0)

    ax.set_xlabel("Count")
    ax.set_ylabel("")
    ax.set_title("Number of times DMR\nselected as optimal feature")

    plt.savefig(
        f"reports/LOO_optimal_features_barplot.pdf", bbox_inches="tight", dpi=600
    )
    plt.close("all")

    """
    Evaluation
    """
    feature_set = {
        k: v.intersection(methy_b.index) for k, v in MLiquidBiopsy.TOP_CPGS.items()
    }
    feature_set["DMR signature"] = set(top_coefs["cpg_cluster"])

    plot_df = dict(
        Binary=dict(
            y_true=Y.loc[pred_loo.index],
            y_pred=pred_loo.loc[pred_loo.index],
            y_proba=prob_loo.loc[pred_loo.index],
        )
    )

    MLiquidBiopsyPlot.eval_barplot(plot_df, palette=features_pal)
    plt.savefig(f"reports/LOO_benchmark_barplots.pdf", bbox_inches="tight", dpi=600)
    plt.close("all")

    MLiquidBiopsyPlot.eval_recall_precision_curve(
        plot_df, palette=features_pal, feature_set=feature_set
    )
    plt.savefig(f"reports/LOO_benchmark_curves.pdf", bbox_inches="tight", dpi=600)
    plt.close("all")

    MLiquidBiopsyPlot.eval_confusion_matrices(plot_df)
    plt.savefig(
        f"reports/LOO_benchmark_confusion_matrix.pdf",
        bbox_inches="tight",
        dpi=600,
    )
    plt.close("all")

    """
    Benchmark
    """
    df_tissue = dict(
        Binary=dict(
            y_true=Y[ss_tissue.index].copy(), y_pred=pred_loo.loc[ss_tissue.index]
        )
    )
    df_cfdna = dict(
        Binary=dict(
            y_true=Y[ss_cfdna.index].copy(), y_pred=pred_loo.loc[ss_cfdna.index]
        )
    )

    df_eval_tissue = MLiquidBiopsyPlot.eval_barplot(
        df_tissue, palette=features_pal, plot_it=False
    )
    df_eval_cfdna = MLiquidBiopsyPlot.eval_barplot(
        df_cfdna, palette=features_pal, plot_it=False
    )

    metrics = ["precision", "recall"]
    dtypes = ["Tissue", "cfDNA"]

    plot_df = pd.concat(
        [
            df_eval_tissue.query(f"metric in @metrics").assign(type="Tissue"),
            df_eval_cfdna.query(f"metric in @metrics").assign(type="cfDNA"),
        ]
    )

    for t in dtypes:
        plot_df_t = pd.pivot_table(
            plot_df.query(f"type == '{t}'"),
            index="ftype",
            columns="metric",
            values="value",
        )

        ax = BinaryTrainingPlot.plot_recall_precision_scatter(
            plot_df_t,
            features,
            Y[ss_cfdna.index if t == "cfDNA" else ss_tissue.index],
            lim=(0.0, 1.0) if t == "cfDNA" else (0.5, 1.0),
        )
        ax.set_title(f"Train & Test\n{t}")

        plt.savefig(
            f"reports/LOO_benchmark_{t}.pdf",
            bbox_inches="tight",
            dpi=600,
        )
        plt.close("all")

    """
    Independent validation
    ["GSE75041", "GSE56588", "GSE54503", "GSE77269", "GSE89852", "GSE136583", "GSE113019"]
    """
    Y_validation_pred, Y_validation_prob = dict(), dict()
    for ftype in features:
        if ftype in MLiquidBiopsy.TOP_CPGS:
            # Benchmark sets
            ftype_cpgs = (
                MLiquidBiopsy.TOP_CPGS[ftype]
                .intersection(methy_b_validation.index)
                .intersection(methy_b.index)
            )
            ftype_x_test = (
                methy_b_validation.reindex(index=ftype_cpgs, columns=Y_validation.index)
                .dropna()
                .T
            )
            ftype_x_train = (
                methy_b.reindex(index=ftype_x_test.columns, columns=Y.index).dropna().T
            )
        else:
            ftype_cpgs = set(top_coefs["cpg_cluster"]).intersection(
                methy_b_validation_cluster.index
            )
            ftype_x_test = (
                methy_b_validation_cluster.reindex(
                    index=ftype_cpgs, columns=Y_validation.index
                )
                .dropna()
                .T
            )
            ftype_x_train = (
                methy_b_cluster.reindex(index=ftype_x_test.columns, columns=Y.index)
                .dropna()
                .T
            )

        clfs = [
            ("Ridge", RidgeClassifierCVwithProba(alphas=[0.1, 0.5, 1, 2, 4, 10])),
            ("Logistic", LogisticRegressionCV(max_iter=1e6, tol=1e-6)),
        ]
        clf = VotingClassifier(estimators=clfs, voting="soft").fit(
            ftype_x_train, Y.loc[ftype_x_train.index]
        )

        Y_validation_pred[ftype] = pd.Series(
            clf.predict(ftype_x_test), index=ftype_x_test.index
        )
        Y_validation_prob[ftype] = pd.Series(
            clf.predict_proba(ftype_x_test)[:, 1], index=ftype_x_test.index
        )

    Y_validation_pred = pd.DataFrame(Y_validation_pred)
    Y_validation_prob = pd.DataFrame(Y_validation_prob)

    # Eval
    plot_df = dict(
        Binary=dict(
            y_true=Y_validation.copy(),
            y_pred=Y_validation_pred.loc[Y_validation.index],
            y_proba=Y_validation_prob.loc[Y_validation.index],
        )
    )

    MLiquidBiopsyPlot.eval_barplot(plot_df, palette=features_pal)
    plt.savefig(
        f"reports/LOO_benchmark_barplots_validation.pdf", bbox_inches="tight", dpi=600
    )
    plt.close("all")

    MLiquidBiopsyPlot.eval_recall_precision_curve(
        plot_df, palette=features_pal, feature_set=feature_set
    )
    plt.savefig(
        f"reports/LOO_benchmark_curves_validation.pdf", bbox_inches="tight", dpi=600
    )
    plt.close("all")

    MLiquidBiopsyPlot.eval_confusion_matrices(plot_df)
    plt.savefig(
        f"reports/LOO_benchmark_confusion_matrix_validation.pdf",
        bbox_inches="tight",
        dpi=600,
    )
    plt.close("all")

    #
    df_validation = dict(
        Binary=dict(
            y_true=Y_validation.copy(), y_pred=Y_validation_pred.loc[Y_validation.index]
        )
    )
    df_eval_validation = MLiquidBiopsyPlot.eval_barplot(
        df_validation, palette=features_pal, plot_it=False
    )

    plot_df = df_eval_validation.query(f"metric in @metrics")
    plot_df_t = pd.pivot_table(plot_df, index="ftype", columns="metric", values="value")

    ax = BinaryTrainingPlot.plot_recall_precision_scatter(
        plot_df_t,
        features,
        Y_validation,
        lim=(0.6, 1),
    )
    ax.set_title("Validation\nTissue")

    plt.savefig(
        f"reports/LOO_benchmark_validation.pdf",
        bbox_inches="tight",
        dpi=600,
    )
    plt.close("all")

    """
    Top regions methylation distributions
    """
    # Methylation regions
    dbs = [
        ("Tissue", list(ss_tissue["label"].unique())),
        ("cfDNA", list(ss_cfdna["label"].unique())),
    ]

    f_clusters = set.union(
        set(
            top_coefs.groupby("cpg_cluster")["count"]
            .first()
            .sort_values(ascending=False)
            .head(10)
            .index
        ),
        {
            "chr17_Cluster178720",
            "chr19_Cluster211956",
            "chr19_Cluster205554",
            "chr10_Cluster63685",
            "chr5_Cluster343774",
            "chr7_Cluster371128",
        },
    )

    for f_cluster in f_clusters:
        # f_cluster = "chr7_Cluster371128"
        for t, ss_subset in dbs:
            feat = cluster_map_cpg[f_cluster]
            ax = MLiquidBiopsyPlot.methylation_region(
                list(feat)[0],
                methy_b[ss_all.query(f"label in @ss_subset").index],
                pannot,
                ss_all,
                hue="Type",
                palette=pal_type,
                hue_order=["HCC", "Other"],
                genomic_range=1e3,
                feature_highlights=feat,
            )

            ax.set_title(f"{t}\n{dmrs.loc[f_cluster, 'name']}")
            plt.legend(
                frameon=False,
                prop=dict(size=6),
                loc="center left",
                bbox_to_anchor=(1, 0.1),
            )
            plt.savefig(
                f"reports/LOO_methylation_region_{f_cluster}_{t}.pdf",
                bbox_inches="tight",
                dpi=600,
            )
            plt.close("all")

    """
    Score iterations
    """
    SCORE_N_ITERATIONS = 1000
    feature_weights_iters = []
    for i in range(SCORE_N_ITERATIONS):
        LOG.info(f"Iterations: {i+1}")
        x_iter, y_iter = build_matrices(X, Y, samples)

        for ftype in features:
            if ftype in MLiquidBiopsy.TOP_CPGS:
                ftype_features = MLiquidBiopsy.TOP_CPGS[ftype].intersection(
                    methy_b.index
                )
                ftype_x_train = (
                    methy_b.reindex(index=ftype_features, columns=y_iter.index)
                    .dropna()
                    .T.copy()
                )

            else:
                ftype_features = feature_set["DMR signature"]
                ftype_x_train = x_iter.reindex(
                    index=y_iter.index, columns=ftype_features
                )

            clf = RidgeClassifier(alpha=1).fit(ftype_x_train, y_iter)
            feature_weights_iters.append(
                pd.Series(clf.coef_[0], index=ftype_features, name="coef")
                .reset_index()
                .assign(feature=ftype, iter=i)
            )
    feature_weights_iters = pd.concat(feature_weights_iters, ignore_index=True)
    feature_weights_iters.to_csv("tables/LOO_feature_weights_iters.csv", index=False)
    # feature_weights_iters = pd.read_csv("tables/LOO_feature_weights_iters.csv")

    fw = feature_weights_iters.query("feature == 'DMR signature'")
    fw = fw.drop("feature", axis=1).rename(columns=dict(index="cpg_cluster"))
    fw["dmr"] = (
        fw["cpg_cluster"].replace(dmrs.loc[fw["cpg_cluster"].unique(), "name"]).values
    )

    """
    
    """
    weight_scores = []

    for ftype in features:
        # ftype = "DMR signature"
        ftype_feature_weights = feature_weights_iters.query(f"feature == '{ftype}'")
        order = ftype_feature_weights.groupby("index")["coef"].mean().sort_values()

        # Boxplots
        _, ax = plt.subplots(1, 1, figsize=(2, 0.2 * len(feature_set[ftype])))
        sns.boxplot(
            "coef",
            "index",
            data=ftype_feature_weights,
            notch=True,
            showcaps=False,
            saturation=1.0,
            orient="h",
            order=list(order.index),
            color=GIPlot.PAL_DTRACE[0],
            boxprops=GIPlot.BOXPROPS,
            whiskerprops=GIPlot.WHISKERPROPS,
            flierprops=GIPlot.FLIERPROPS,
            medianprops=dict(linestyle="-", linewidth=1.0, c="red"),
            ax=ax,
        )
        if ftype == "DMR signature":
            ax.set_yticklabels(
                [dmrs.loc[t.get_text(), "name"] for t in ax.get_yticklabels()]
            )

        ax.grid(axis="x", lw=0.1, color="#e1e1e1", zorder=0)
        ax.set_xlabel("Coefficients\nLinear classifier")
        ax.set_ylabel("")
        plt.savefig(
            f"reports/LOO_weights_boxplots_{ftype}.pdf", bbox_inches="tight", dpi=600
        )
        plt.close("all")

        #
        if ftype == "DMR signature":
            ftype_val_m = methy_b_validation_cluster.loc[order.index].dropna().T
            sample_scores = ftype_val_m.dot(order.loc[ftype_val_m.columns])

        else:
            ftype_val_m = methy_b_validation.loc[order.index].dropna().T
            sample_scores = ftype_val_m.dot(order.loc[ftype_val_m.columns])

        weight_scores.append(sample_scores.rename(ftype))

    weight_scores = pd.concat(weight_scores, axis=1)
    weight_scores["y_true"] = ss_all_validation.loc[
        weight_scores.index, "y_true"
    ].values
    weight_scores.to_excel(f"tables/LOO_weights_scores.xlsx")

    _, ax = plt.subplots(1, 1, figsize=(2, 2))

    for ftype in features:
        _, t, r, p = MLiquidBiopsyPlot.recall_precision_curve(
            weight_scores["y_true"],
            weight_scores[ftype],
            color=features_pal[ftype],
            plot_legend=False,
            plot_random=False,
            plot_f1curves=ftype == "DMR signature",
            ax=ax,
        )

    ax.legend(
        frameon=False,
        loc=3,
        prop=dict(size=4),
    )

    ax.axhline(
        weight_scores["y_true"].sum() / weight_scores["y_true"].shape[0],
        c="#d9d9d9",
        lw=1.0,
        label="random",
        zorder=0,
        ls="--",
    )

    plt.savefig(
        f"reports/LOO_weights_precision_recall_curves_validation.pdf",
        bbox_inches="tight",
        dpi=600,
    )
    plt.close("all")

    """
    
    """
    weight_scores_ci = []

    _, ax = plt.subplots(1, 1, figsize=(2, 2))

    for i in range(SCORE_N_ITERATIONS):
        iter_weights = feature_weights_iters.query(
            f"(feature == 'DMR signature') & (iter == {i})"
        )
        iter_weights = iter_weights.groupby("index")["coef"].mean().sort_values()

        iter_scores = pd.concat(
            [
                methy_b_cluster.loc[iter_weights.index]
                .T.dot(iter_weights)
                .rename("score"),
                samples["label"].apply(MLiquidBiopsy.sample_class).rename("y_true"),
            ],
            axis=1,
        ).loc[samples.index]

        _, t, r, p = MLiquidBiopsyPlot.recall_precision_curve(
            iter_scores["y_true"],
            iter_scores["score"],
            color=CrispyPlot.PAL_DTRACE[0],
            plot_legend=False,
            plot_random=False,
            plot_f1curves=i == 0,
            ax=ax,
        )

        res = iter_weights.to_dict()
        res["thres"] = t
        res["recall"] = r
        res["precision"] = p

        weight_scores_ci.append(res)

    ax.axhline(
        weight_scores["y_true"].sum() / weight_scores["y_true"].shape[0],
        c="#d9d9d9",
        lw=1.0,
        label="random",
        zorder=0,
        ls="--",
    )

    plt.savefig(
        f"reports/LOO_weights_precision_recall_curves_ci.pdf",
        bbox_inches="tight",
        dpi=600,
    )
    plt.close("all")

    weight_scores_ci = pd.DataFrame(weight_scores_ci)
    weight_scores_ci.to_excel("tables/LOO_threshold_scores.xlsx", index=False)
    # weight_scores_ci = pd.read_excel("tables/LOO_threshold_scores.xlsx")

    """
    Top DMRs downsample evaluation
    """
    dmrs_order = weight_scores_ci.mean().drop(["thres", "recall", "precision"])
    dmrs_order = dmrs_order[dmrs_order.abs().sort_values(ascending=False).index]

    dmrs_order_df = []
    for i in range(1, len(dmrs_order) + 1):
        LOG.info(f"Number of features = {i}")

        # Subset
        order_x_train = X[dmrs_order.head(i).index]
        order_x_test = X_validation[dmrs_order.head(i).index]

        # Train
        clfs = [
            ("Ridge", RidgeClassifierCVwithProba(alphas=[0.1, 0.5, 1, 2, 4, 10])),
            ("Logistic", LogisticRegressionCV(max_iter=1e6, tol=1e-6)),
        ]
        clf = VotingClassifier(estimators=clfs, voting="soft").fit(
            order_x_train, Y.loc[order_x_train.index]
        )

        # Predict
        Y_validation_pred = pd.Series(clf.predict(order_x_test), index=order_x_test.index)
        Y_validation_prob = pd.Series(clf.predict_proba(order_x_test)[:, 1], index=order_x_test.index)

        # Eval
        Y_validation_eval = MLiquidBiopsy.eval_pred(
            Y_validation.loc[order_x_test.index],
            Y_validation_pred.loc[order_x_test.index],
            as_str=False,
        )

        # Store
        dmrs_order_dict = dict(
            etype="validation",
            n_features=i,
        )
        dmrs_order_df.append({**dmrs_order_dict, **Y_validation_eval})

    dmrs_order_df = pd.DataFrame(dmrs_order_df)
    dmrs_order_df.to_csv(f"tables/LOO_sfs_top_dmr_eval.csv", index=False)

    # Lineplot
    plot_df = pd.melt(
        dmrs_order_df,
        id_vars=["etype", "n_features"],
        value_vars=["recall", "precision"],
        var_name="metric",
    )

    fig, ax = plt.subplots(1, 1, figsize=(3, 2.5), sharey="all", sharex="all")

    sns.lineplot(
        x="n_features",
        y="value",
        style="metric",
        data=plot_df,
        color=CrispyPlot.PAL_DTRACE[4],
        err_kws=dict(lw=0),
        ax=ax,
    )

    ax.legend(
        frameon=False,
        title="",
        prop={"size": 5},
        loc=4,
    )

    ax.grid(True, ls="-", lw=0.1, alpha=1.0, zorder=0)
    ax.set_xlabel("Top N DMRs")
    ax.set_ylabel("Metric value")

    plt.savefig(f"reports/LOO_sfs_top_DMR_eval_lineplot.pdf", bbox_inches="tight", dpi=600)
    plt.close("all")

    """
    Samples score
    """
    sample_scores_all = []

    for ftype in features:
        ftype_feature_weights = feature_weights_iters.query(f"feature == '{ftype}'")
        order = ftype_feature_weights.groupby("index")["coef"].mean().sort_values()

        if ftype == "DMR signature":
            ftype_val_m = methy_b_validation_cluster.loc[order.index].dropna().T
            sample_scores_val = ftype_val_m.dot(order.loc[ftype_val_m.columns])

            ftype_tt_m = methy_b_cluster.loc[order.index].dropna().T
            sample_scores_tt = ftype_tt_m.dot(order.loc[ftype_tt_m.columns])

        else:
            ftype_val_m = methy_b_validation.loc[order.index].dropna().T
            sample_scores_val = ftype_val_m.dot(order.loc[ftype_val_m.columns])

            ftype_tt_m = methy_b.loc[order.index].dropna().T
            sample_scores_tt = ftype_tt_m.dot(order.loc[ftype_tt_m.columns])

        sample_scores_all.append(
            pd.concat([sample_scores_val, sample_scores_tt]).rename(ftype)
        )

    sample_scores_all = pd.DataFrame(sample_scores_all).T
    sample_scores_all = pd.concat(
        [
            pd.concat(
                [
                    ss_all[["Type", "dataset_id"]],
                    ss_all_validation[["Type", "dataset_id"]],
                ]
            ),
            sample_scores_all,
        ],
        axis=1,
    ).sort_values("DMR signature", ascending=False)
    sample_scores_all.to_excel("tables/LOO_sample_scores_all.xlsx")
    # sample_scores_all = pd.read_excel("tables/LOO_sample_scores_all.xlsx", index_col=0)

    """
    DMR signature Risk Score
    """
    feature_weights = (
        feature_weights_iters.query(f"feature == 'DMR signature'")
        .groupby("index")["coef"]
        .mean()
    )
    plot_df = pd.concat(
        [
            methy_b_cluster.loc[feature_weights.index]
            .T.dot(feature_weights)
            .rename("score"),
            methy_b_cluster.loc[f_clusters].T,
            ss_all,
        ],
        axis=1,
    )
    plot_df["y_true"] = plot_df["label"].apply(MLiquidBiopsy.sample_class).values
    plot_df = plot_df.rename(columns=dmrs["name"])

    pal = {
        "Other": CrispyPlot.PAL_DTRACE[0],
        "HCC": CrispyPlot.PAL_DTRACE[1],
        "Cancer": CrispyPlot.PAL_DTRACE[3],
        "Healthy": "#63BF55",
    }

    for subss in ["Tissue", "cfDNA"]:
        if subss == "cfDNA":
            subss_list = list(set(ss_cfdna["label"])) + ["cfDNA"]
            plot_df_subss = plot_df.query("label in @subss_list")

            cfdna_cancer = {
                "GSM3455808_200687110099_R05C01",
                "GSM3455809_200687110099_R06C01",
                "GSM3455810_200687110099_R07C01",
                "GSM3455811_200687110099_R08C01",
                "GSM3455841_202410280057_R05C01",
                "GSM3455842_202410280057_R06C01",
                "GSM3455843_202410280057_R07C01",
                "GSM3455844_202410280057_R08C01",
                "GSM3455849_202410280077_R05C01",
                "GSM3455850_202410280077_R06C01",
                "GSM3455851_202410280077_R07C01",
            }

            cfdna_healthy = set(
                plot_df[
                    plot_df["label"].isin(["cfDNA healthy", "cfDNA healthy pooled"])
                ].index
            )

            plot_df_subss.loc[cfdna_cancer, "y_true"] = 1
            plot_df_subss.loc[cfdna_cancer, "Type"] = "Cancer"

            plot_df_subss.loc[cfdna_healthy, "y_true"] = 0
            plot_df_subss.loc[cfdna_healthy, "Type"] = "Healthy"

            hue_order = ["Other", "Healthy", "Cancer", "HCC"]

        else:
            subss_list = [
                "Liver obese",
                "HCC + EtOH",
                "Solid Tissue Normal - Liver",
                # 'Primary Tumor - Liver',
                # 'Cirrhosis + HCV',
                "Liver normal",
                # 'Cirrhosis + AATD',
                # 'Cirrhosis + Other',
                # 'Cirrhosis + HBV',
                "HCC + HCV",
                # 'Cirrhosis + EtOH',
                "AATD",
            ]
            plot_df_subss = plot_df.query("label in @subss_list")

            hue_order = ["Other", "HCC"]

        ax, t, r, p = MLiquidBiopsyPlot.recall_precision_curve(
            plot_df_subss["y_true"],
            plot_df_subss["score"],
            color=CrispyPlot.PAL_DTRACE[2],
        )
        ax.set_title(
            f"Train & Test\n{subss} - Holdout samples"
            if subss == "Tissue"
            else f"Train & Test\n{subss} - All samples"
        )
        plt.savefig(
            f"reports/LOO_score_recall_curve_{subss}.pdf", bbox_inches="tight", dpi=600
        )
        plt.close("all")

        for y_var in dmrs.loc[f_clusters, "name"]:
            ax = GIPlot.gi_tissue_plot(
                "score",
                y_var,
                plot_df=plot_df_subss,
                hue="Type",
                hue_order=hue_order,
                plot_reg=False,
                annot=False,
                pal=pal,
            )

            ax.axvline(
                t,
                lw=1,
                ls="--",
                c=CrispyPlot.PAL_DTRACE[2],
            )
            ax.text(
                t + 0.01,
                ax.get_ylim()[0],
                f"Recall={r:.2f}\nPrecision={p:.2f}",
                va="bottom",
                color=CrispyPlot.PAL_DTRACE[2],
                fontsize=8,
                zorder=3,
            )

            ax.set_title(
                f"Train & Test\n{subss} - Holdout samples"
                if subss == "Tissue"
                else f"Train & Test\n{subss} - All samples"
            )

            ax.set_xlabel(f"DMR signature score")
            ax.set_ylabel(f"{y_var}\nDNA methylation (beta)")

            plt.savefig(
                f"reports/LOO_score_scatter_{subss}_{y_var}.pdf",
                bbox_inches="tight",
                transparent=True,
                dpi=600,
            )
            plt.close("all")

    """
    Independent validation - DMR signature score
    """
    train_thres = weight_scores_ci["thres"].mean()
    train_thres_sd = weight_scores_ci["thres"].std()

    # Sample scores
    sample_scores_test = methy_b_validation_cluster.loc[feature_weights.index].T.dot(
        feature_weights
    )
    sample_scores_test = pd.concat(
        [
            sample_scores_test.rename("score"),
            methy_b_validation_cluster.loc[f_clusters].T,
            ss_all_validation,
        ],
        axis=1,
    )
    sample_scores_test["Type"] = (
        sample_scores_test["y_true"].replace({1: "HCC", 0: "Other"}).values
    )
    sample_scores_test = sample_scores_test.rename(columns=dmrs["name"])
    sample_scores_test[["score", "dataset_id", "Type"]].to_excel(
        f"tables/LOO_validation_scores.xlsx"
    )

    # Test scores - Precision/Recall curve
    (
        ax,
        test_thres,
        test_recall,
        test_precision,
    ) = MLiquidBiopsyPlot.recall_precision_curve(
        sample_scores_test["y_true"],
        sample_scores_test["score"],
        color=CrispyPlot.PAL_DTRACE[2],
    )
    ax.set_title(f"Validation\nTissue")
    plt.savefig(
        f"reports/LOO_score_recall_curve_validation.pdf", bbox_inches="tight", dpi=600
    )
    plt.close("all")

    # Test scores - Score scatters
    hue_order = ["Other", "HCC"]

    for y_var in dmrs.loc[f_clusters, "name"]:
        ax = GIPlot.gi_tissue_plot(
            "score",
            y_var,
            plot_df=sample_scores_test,
            hue="Type",
            hue_order=hue_order,
            plot_reg=False,
            annot=False,
            pal=pal,
        )

        ax.axvline(
            train_thres,
            lw=0.5,
            ls="--",
            c=CrispyPlot.PAL_DTRACE[2],
            zorder=0,
        )
        ax.axvspan(
            train_thres - train_thres_sd,
            train_thres + train_thres_sd,
            alpha=0.3,
            color=CrispyPlot.PAL_DTRACE[0],
            zorder=0,
            lw=0,
        )

        r = recall_score(
            sample_scores_test["y_true"],
            (sample_scores_test["score"] > train_thres).astype(int),
        )
        p = precision_score(
            sample_scores_test["y_true"],
            (sample_scores_test["score"] > train_thres).astype(int),
        )
        ax.text(
            train_thres + 0.01,
            ax.get_ylim()[0],
            f"Mean Train & Test threshold\nRecall={r:.2f}, Precision={p:.2f}",
            va="bottom",
            color=CrispyPlot.PAL_DTRACE[2],
            fontsize=5,
            zorder=3,
        )

        ax.set_xlabel(f"DMR signature score")
        ax.set_ylabel(f"{y_var}\nDNA methylation (beta)")
        ax.set_title(f"Validation\nTissue")

        ax.grid(True, ls="-", lw=0.1, alpha=1.0, zorder=0)

        plt.savefig(
            f"reports/LOO_independent_score_scatter_validation_{y_var}.pdf",
            bbox_inches="tight",
            transparent=True,
            dpi=600,
        )
        plt.close("all")
```
