## Supplementary File 1 for "DNA methylation fingerprint for the diagnosis and monitoring of hepatocellular carcinoma from tissue and liquid biopsies": BinaryTrainingPlot.py

```
import numpy as np
import pandas as pd
import seaborn as sns
import matplotlib.pyplot as plt
from crispy import CrispyPlot
from crispy.GIPlot import GIPlot
from matplotlib.lines import Line2D
from adjustText import adjust_text
from MLiquidBiopsy import MLiquidBiopsyPlot, MLiquidBiopsy


class BinaryTrainingPlot:
    @classmethod
    def purity_regression(cls, pc, pcs, vexp, methy_b_mean_corr_pc5, pal):
        g = GIPlot.gi_regression(pc, "purity_CPE", pcs.dropna(subset=[pc, "purity_CPE"]), hue="Type", palette=pal)
        g.set_axis_labels(f"{pc} Train & Test Methylation\n(Var. exp. {vexp[pc] * 100:.1f}%)", "Tumour Purity")
        plt.savefig(f"reports/LOO_tumour_purity_{pc}_corrplot.pdf", bbox_inches="tight", dpi=600)
        plt.close("all")

        _, ax = plt.subplots(1, 1, figsize=(2, 1.5), sharex="all", sharey="all")

        sns.distplot(
            methy_b_mean_corr_pc5,
            hist=True,
            kde=False,
            bins=100,
            hist_kws=dict(linewidth=0, alpha=1),
            color=CrispyPlot.PAL_DTRACE[2],
            ax=ax,
        )

        ax.grid(True, ls="-", lw=0.1, alpha=1.0, zorder=0)
        ax.set_xlabel("CpG correlation with PC5\n(Pearson's R)")
        ax.set_ylabel("Number of CpGs")
        plt.savefig(
            f"reports/LOO_global_methylation_corr_{pc}_distplot.pdf",
            bbox_inches="tight",
            dpi=600,
        )
        plt.close("all")

    @classmethod
    def purity_corr_pcs_barplot(cls, pcs, vexp):
        plot_df = pd.concat([
            pcs.corr()["purity_CPE"].rename("corr"),
            vexp.rename("variance_explained"),
        ], axis=1).dropna().sort_values("corr").reset_index()

        order = list(plot_df["index"])

        fig, ax = plt.subplots(1, 1, figsize=(3, 0.2 * len(order)))

        sns.barplot(
            x="corr",
            y="index",
            order=order,
            data=plot_df,
            color=CrispyPlot.PAL_DTRACE[2],
            saturation=1,
            lw=0,
            ax=ax,
        )

        for y_var in order:
            t = f"{plot_df.loc[order.index(y_var), 'variance_explained'] * 100:,.1f}%"
            x = plot_df.loc[order.index(y_var), 'corr']
            ax.text(
                x,
                order.index(y_var),
                t,
                fontsize=6,
                ha="right" if x < 0 else "left",
                va="center",
            )

        ax.grid(axis="x", lw=0.1, color="#e1e1e1", zorder=0)

        ax.set_xlabel("Correlation with Tumour Purity\nPearson's R")
        ax.set_ylabel("Train & Test Methylation PCs")
        ax.set_xlim(-0.5, 0.7)

        plt.savefig(f"reports/LOO_purity_pcs_corr_barplot.pdf", bbox_inches="tight", dpi=600)
        plt.close("all")

    @classmethod
    def pairplot_pca(cls, pcs, vexp, ss, hue_by="Type", pal=None, y_vars=None):
        if pal is None:
            pal = {'Other': '#E1E1E1', 'HCC': '#fc8d62'}

        if y_vars is None:
            y_vars = ["PC1", "PC2", "PC3"]

        plot_df = pd.concat([pcs[y_vars], ss[hue_by]], axis=1)

        g = sns.PairGrid(
            data=plot_df,
            vars=y_vars,
            despine=False,
            size=1.5,
            hue=hue_by,
            palette=pal,
            height=2,
        )

        g = g.map_diag(sns.histplot, linewidth=0, alpha=0.5)
        g = g.map_offdiag(plt.scatter, s=8, edgecolor="white", lw=0.1, alpha=0.8)
        g = g.add_legend()

        for i, ax in enumerate(g.axes):
            ax[0].set_ylabel(f"PC{i+1} ({vexp[f'PC{i+1}']*100:.1f}%)")

        for i, ax in enumerate(g.axes[len(y_vars) - 1]):
            ax.set_xlabel(f"PC{i+1} ({vexp[f'PC{i+1}']*100:.1f}%)")

    @staticmethod
    def dim_red(df, name, ss, pal, n_components=2, hue_by="label", is_plot=True, is_export=True):
        methy_pcs, methy_umap, methy_vexp = MLiquidBiopsy.dimension_reduction(
            df, ss, n_components=n_components
        )

        if is_export:
            methy_pcs.to_csv(f"tables/LOO_{name}_PCs.csv")
            methy_umap.to_csv(f"tables/LOO_{name}_UMAP.csv")

        if is_plot:
            MLiquidBiopsyPlot.dimension_reduction_plot(
                "PC1", "PC2", hue_by, methy_pcs, vexp=methy_vexp, palette=pal
            )
            plt.savefig(f"reports/LOO_{name}_PCA.pdf", bbox_inches="tight", dpi=600)
            plt.close("all")

            MLiquidBiopsyPlot.dimension_reduction_plot(
                "Dim1", "Dim2", hue_by, methy_umap, method="UMAP", palette=pal
            )
            plt.savefig(f"reports/LOO_{name}_UMAP.pdf", bbox_inches="tight", dpi=600)
            plt.close("all")

        return methy_pcs, methy_umap, methy_vexp

    @staticmethod
    def plot_pca(pcs, pcs_to_plot, ss, vexp, pal, name="methy"):
        for x_var, y_var in pcs_to_plot:
            plot_df = pd.concat([pcs, ss["Type"]], axis=1)

            MLiquidBiopsyPlot.dimension_reduction_plot(
                x_var,
                y_var,
                "Type",
                plot_df,
                vexp=vexp,
                palette=pal,
                hue_order=["Other", "HCC"],
            )
            plt.savefig(
                f"reports/LOO_{name}_PCA_{x_var}_{y_var}.pdf",
                bbox_inches="tight",
                dpi=600,
            )
            plt.close("all")

    @staticmethod
    def plot_number_samples_per_dataset(ss, name, color=CrispyPlot.PAL_DTRACE[2], outpath=None):
        if outpath is None:
            outpath = f"reports/LOO_{name}_dataset_countplot.pdf"

        plot_df = ss["dataset_id"].value_counts().sort_values(ascending=True).rename("count").reset_index()

        fig, ax = plt.subplots(1, 1, figsize=(2, plot_df.shape[0] * 0.2))

        ax.barh(
            plot_df.index,
            plot_df["count"],
            color=color,
            linewidth=0,
        )

        for y_var in plot_df.index:
            ax.text(
                plot_df.loc[y_var, "count"],
                y_var,
                f"{plot_df.loc[y_var, 'count']:.0f}",
                fontsize=6,
                va="center",
            )

        ax.grid(axis="x", lw=0.1, color="#e1e1e1", zorder=0)

        ax.set_xlabel("Number of samples")
        ax.set_ylabel("Datasets")

        ax.set_yticks(plot_df.index)
        ax.set_yticklabels(plot_df["index"])

        plt.savefig(
            outpath, bbox_inches="tight", dpi=600
        )
        plt.close("all")

    @staticmethod
    def plot_number_samples(ss, pal, outpath=f"reports/LOO_sample_types_countplot.pdf"):
        # Number of samples
        plot_df = ss["label"].value_counts().sort_values().reset_index()
        plot_df = pd.concat(
            [
                plot_df,
                plot_df["index"]
                .apply(MLiquidBiopsy.sample_class)
                .replace({0: "Other", 1: "HCC"})
                .rename("Type"),
            ],
            axis=1,
        )

        order = list(plot_df.index)

        fig, ax = plt.subplots(1, 1, figsize=(2, 2.5))

        ax.barh(
            plot_df.index,
            plot_df["label"],
            color=[pal[c] for c in plot_df.loc[order, "Type"]],
            linewidth=0,
        )

        for y_var in order:
            ax.text(
                plot_df.loc[y_var, "label"],
                order.index(y_var),
                f"{plot_df.loc[y_var, 'label']:.0f}",
                fontsize=6,
                va="center",
            )

        ax.grid(axis="x", lw=0.1, color="#e1e1e1", zorder=0)

        ax.set_xlabel("Number of samples")
        ax.set_ylabel("Sample types")

        ax.set_yticks(order)
        ax.set_yticklabels(plot_df.loc[order, "index"])

        ax.legend(
            [Line2D([0], [0], color=pal[k], lw=4) for k in pal],
            list(pal.keys()),
            loc=4,
            frameon=False,
            prop=dict(size=5),
        )

        plt.savefig(
            outpath, bbox_inches="tight", dpi=600
        )
        plt.close("all")

    @staticmethod
    def plot_global_corr_hist(corr_df):
        _, ax = plt.subplots(1, 1, figsize=(2, 1.5), sharex="all", sharey="all")

        sns.distplot(
            corr_df,
            hist=True,
            kde=False,
            bins=100,
            hist_kws=dict(linewidth=0, alpha=1),
            color=CrispyPlot.PAL_DTRACE[2],
            ax=ax,
        )

        ax.grid(True, ls="-", lw=0.1, alpha=1.0, zorder=0)
        ax.set_xlabel("CpG correlation with global methylation\n(Pearson's R)")
        ax.set_ylabel("Number of CpGs")
        plt.savefig(
            f"reports/LOO_global_methylation_corr_distplot.pdf",
            bbox_inches="tight",
            dpi=600,
        )
        plt.close("all")

    @staticmethod
    def plot_clusters_length(pannot, methy_b_, methy_b_cluster):
        cluster_size = (
            pannot.loc[methy_b_.index]
            .groupby("cluster")["Start_hg38"]
            .agg(["max", "min"])
            .eval("max - min")
        )
        cluster_size = cluster_size.loc[methy_b_cluster.index]

        _, ax = plt.subplots(1, 1, figsize=(2, 1.5), sharex="all", sharey="all")

        sns.distplot(
            cluster_size,
            hist=True,
            kde=False,
            bins=100,
            hist_kws=dict(linewidth=0, alpha=1),
            color=CrispyPlot.PAL_DTRACE[2],
            ax=ax,
        )

        ax.axvline(
            np.median(cluster_size),
            c="red",
            ls="--",
            lw=0.5,
        )
        ax.text(
            np.median(cluster_size),
            10,
            f"Median size: {np.median(cluster_size)}",
            fontsize=6,
            ha="left",
            c="white",
        )

        ax.grid(True, ls="-", lw=0.1, alpha=1.0, zorder=0)
        ax.set_xlabel("CpG cluster base-pair")
        ax.set_ylabel("Number of CpGs clusters")
        plt.savefig(
            f"reports/LOO_cpg_cluster_size_distplot.pdf", bbox_inches="tight", dpi=600
        )
        plt.close("all")

    @staticmethod
    def plot_features_corr_heatmap(df):
        fig = sns.clustermap(
            df,
            cmap="Spectral",
            center=0,
            annot=True,
            fmt=".2f",
            annot_kws={"size": 5},
            figsize=(8.5, 8.5),
        )

        fig.ax_heatmap.set_xlabel("CpG clusters")
        fig.ax_heatmap.set_ylabel("CpG clusters")

        fig.ax_heatmap.set_yticklabels(fig.ax_heatmap.get_yticklabels(), rotation=0)
        plt.savefig(f"reports/LOO_features_corr_heatmap.pdf", bbox_inches="tight", dpi=600)
        plt.close("all")

    @staticmethod
    def plot_feature_reduction_barplot(df_dict):
        plot_df = pd.concat([
            pd.DataFrame(dict(filter=k, n_features=v)) for k, v in df_dict.items()
        ])

        order = list(df_dict)

        fig, ax = plt.subplots(1, 1, figsize=(3, 0.2 * len(plot_df["filter"].unique())))

        sns.barplot(
            x="n_features",
            y="filter",
            order=order,
            data=plot_df,
            color=CrispyPlot.PAL_DTRACE[2],
            saturation=1,
            lw=0,
            ax=ax,
        )

        for y_var in order:
            if y_var in ["ANOVAs", "SVM features"]:
                t = f"Median = {np.median(df_dict[y_var]):,.0f}"
                ax.text(
                    np.max(df_dict[y_var]) * 1.05,
                    order.index(y_var),
                    t,
                    fontsize=6, va="center",
                )
            else:
                t = f"N = {df_dict[y_var][0]:,.0f}"
                ax.text(
                    np.max(df_dict[y_var]) * 0.9,
                    order.index(y_var),
                    t, ha="right", color="white",
                    fontsize=6, va="center",
                )

        ax.grid(axis="x", lw=0.1, color="#e1e1e1", zorder=0)

        ax.set_xlabel("Number of features (log10)")
        ax.set_ylabel("Filtering steps")

        plt.xscale('log', base=10)

        plt.savefig(f"reports/LOO_neaftures_reduction_barplot.pdf", bbox_inches="tight", dpi=600)
        plt.close("all")

        return int(np.median(df_dict["SVM features"]))

    @staticmethod
    def palette_feature():
        features = ["DMR signature"] + list(MLiquidBiopsy.TOP_CPGS)
        features_pal = sns.color_palette("tab20c").as_hex()
        features_pal = pd.Series(
            [features_pal[0]] + features_pal[4 : 4 + len(features) - 1], index=features
        )
        return features_pal

    @staticmethod
    def plot_recall_precision_scatter(df, order, y_true, lim=(0, 1), adjust_annotation=True, patent_palette=False):
        plot_df = df.loc[order].reset_index()
        plot_df["markers"] = plot_df["ftype"].apply(MLiquidBiopsyPlot.sample_markers).values
        plot_df["index"] = list(plot_df.index + 1)

        _, ax = plt.subplots(1, 1, figsize=(1.5, 1.5))

        for f_score in np.linspace(0.2, 0.8, num=4):
            if f_score > min(lim):
                f_x = np.linspace(0.01, 1., 1000)
                f_y = f_score * f_x / (2 * f_x - f_score)

                ax.plot(
                    f_x[(f_y >= 0) & (f_y <= 1.5)],
                    f_y[(f_y >= 0) & (f_y <= 1.5)],
                    color="#CCCCCC" if patent_palette else "gray",
                    alpha=1 if patent_palette else 0.2,
                    lw=0.5,
                )

                ax.annotate(
                    f"f1={f_score:0.1f}", xy=(1, f_score / (2 - f_score)),
                    fontsize=5,
                    ha="right",
                )

        labels = []

        for f in order:
            f_df = plot_df.query(f"ftype == '{f}'").iloc[0]

            ax.scatter(
                x=f_df["recall"],
                y=f_df["precision"],
                marker=f_df["markers"],
                c=MLiquidBiopsyPlot.sample_color(f, patent_color=patent_palette),
                label=f"{f_df['index']}. {f}",
                s=MLiquidBiopsyPlot.sample_size(f),
            )

            labels.append(
                ax.text(
                    f_df["recall"],
                    f_df["precision"],
                    f_df["index"],
                    fontsize=5,
                )
            )

        if adjust_annotation:
            adjust_text(
                labels,
                arrowprops=dict(arrowstyle="-", color="#CCCCCC", alpha=0.75, lw=0.3),
                avoid_points=False,
                avoid_text=False,
                avoid_self=False,
            )

        ax.axhline(
            y_true.sum() / y_true.shape[0],
            c="#CCCCCC" if patent_palette else "#d9d9d9",
            lw=1.0,
            label="random",
            zorder=0,
            ls="--",
        )

        ax.set_xlabel("Recall")
        ax.set_ylabel("Precision")
        ax.grid(axis="both", lw=0.1, color="#CCCCCC" if patent_palette else "#e1e1e1", zorder=0)

        ax.set_xticks(np.linspace(min(lim), 1, num=6))
        ax.set_yticks(np.linspace(min(lim), 1, num=6))

        ax.set_xlim(lim)
        ax.set_ylim(lim)

        plt.legend(
            frameon=False,
            loc="center left",
            bbox_to_anchor=(1, 0.5),
            prop={"size": 5},
        )

        return ax
```
