## Supplementary File 1 for "DNA methylation fingerprint for the diagnosis and monitoring of hepatocellular carcinoma from tissue and liquid biopsies": BinaryTrainingSignature.py

```
import numpy as np
import pandas as pd
import seaborn as sns
from LiquidBiopsy import LOG
from crispy import CrispyPlot
import matplotlib.pyplot as plt
from crispy.GIPlot import GIPlot
from sklearn.preprocessing import MinMaxScaler
from BinaryTrainingPlot import BinaryTrainingPlot
from MLiquidBiopsy import MLiquidBiopsy, MLiquidBiopsyPlot
from sklearn.linear_model import (
    PassiveAggressiveClassifier,
    LogisticRegressionCV,
    RidgeClassifierCV,
    Perceptron,
    RidgeClassifier,
)

LABELS = [
    "Primary Tumor - Liver",
    "HCC + EtOH",
    "HCC + HCV",
    "cfDNA HCC and cirrhosis",
    "cfDNA cirrhosis only",
    "Cirrhosis + EtOH",
    "Cirrhosis + HCV",
    "Cirrhosis + HBV",
    "Cirrhosis + Other",
    "Cirrhosis + AATD",
    "AATD",
    "Liver obese",
    "Solid Tissue Normal - Liver",
    "Liver normal",
    "cfDNA",
    "cfDNA healthy pooled",
    "cfDNA healthy",
]

LABELS_CFDNA = [
    "cfDNA HCC and cirrhosis",
    "cfDNA cirrhosis only",
    "cfDNA healthy",
    "cfDNA healthy pooled",
]


def build_matrices(x_train, y_train, ss, labels_hepatocc=None, labels_cirrhosis=None):
    if labels_hepatocc is None:
        labels_hepatocc = [
            "Primary Tumor - Liver",
        ]

    if labels_cirrhosis is None:
        labels_cirrhosis = [
            "Cirrhosis + HBV",
            "Cirrhosis + AATD",
            "Cirrhosis + HCV",
            "Cirrhosis + EtOH",
        ]

    ss = ss.query(f"dataset_id != 'GSE60753' | PC2 < 200")

    nmin_samples = min(
        ss.query("label == 'cfDNA HCC and cirrhosis'").sample(frac=1.0).shape[0],
        ss.query("label == 'cfDNA cirrhosis only'").sample(frac=1.0).shape[0],
    )

    x_train_ = pd.concat(
        [
            x_train.loc[
                ss.query("label in @labels_hepatocc").sample(nmin_samples).index
            ],
            x_train.loc[
                ss.query("label == 'cfDNA HCC and cirrhosis'")
                .sample(nmin_samples)
                .index
            ],
            x_train.loc[
                ss.query("label in @labels_cirrhosis").sample(nmin_samples).index
            ],
            x_train.loc[
                ss.query("label == 'cfDNA cirrhosis only'").sample(nmin_samples).index
            ],
        ]
    ).copy()

    y_train_ = y_train[x_train_.index].copy()

    return x_train_, y_train_


if __name__ == '__main__':
    """
    Imports
    """
    # Samplesheet
    methy_b, ss_all, pal_label, pal_type = MLiquidBiopsy.import_dataset(
        f"~/datasets/DNAMethylationPipeline_MergedSubset_methy_b.csv",
        subset_labels=LABELS,
    )

    # Probe annotation information
    pannot = MLiquidBiopsy.read_probe_annot()

    # Dataset palette
    pal_dataset = ss_all.groupby("dataset_id")["dataset_color"].first()

    # Feature palette
    features = ["DMR signature"] + list(MLiquidBiopsy.TOP_CPGS)
    markers = ["o"] + ["x"] * len(list(MLiquidBiopsy.TOP_CPGS))
    linestyles = ["-"] + ["--"] * len(list(MLiquidBiopsy.TOP_CPGS))
    features_pal = BinaryTrainingPlot.palette_feature()

    """
    Feature reduction
    """
    # Drop NaNs
    methy_b = methy_b.dropna()

    # Dimension reduction
    pcs, dims, vexp = BinaryTrainingPlot.dim_red(
        methy_b.T, "methy", ss_all, pal_label, n_components=10
    )

    # Drop probes not in hg38 lift annotation
    methy_b_ = methy_b.loc[methy_b.index.isin(pannot.index)].copy()

    # Drop sex chromosome
    psex = MLiquidBiopsy.probes_sex_chr()
    methy_b_ = methy_b_.drop(index=psex, errors="ignore")

    # Global methylation
    methy_b_mean = methy_b_.T.mean(1)
    methy_b_mean_corr = methy_b_.T.corrwith(methy_b_mean)
    BinaryTrainingPlot.plot_global_corr_hist(methy_b_mean_corr)

    ss_all["GlobalMethylation"] = methy_b_mean[ss_all.index].values

    # Tumour purity
    tumour_purity = ss_all["purity_CPE"].dropna()
    methy_b_mean_corr_purity = methy_b_[tumour_purity.index].T.corrwith(tumour_purity)
    methy_b_mean_corr_pc5 = methy_b_[pcs.index].T.corrwith(pcs["PC5"])

    ss_all["SamplePurityPC5"] = pcs.loc[ss_all.index, "PC5"].values

    # Cluster methylation
    methy_b_cluster, cluster_map = MLiquidBiopsy.probes_cluster(methy_b_, nmin=3)
    cluster_map_cpg = cluster_map.reset_index().groupby("cluster")["IlmnID"].agg(set)

    Y = ss_all["label"].apply(MLiquidBiopsy.sample_class)
    X = methy_b_cluster[Y.index].T

    """
    Validation set
    """
    ss_all_validation, methy_b_validation = MLiquidBiopsy.read_test_datasets("all")
    ss_all_validation["Type"] = ss_all_validation["y_true"].replace({0: "Other", 1: "HCC"})

    methy_b_validation_cluster = methy_b_validation[methy_b_validation.index.isin(methy_b_.index)]
    methy_b_validation_cluster, _ = MLiquidBiopsy.probes_cluster(methy_b_validation_cluster, nmin=3)

    """
    Matrices
    """
    Y_validation = ss_all_validation["y_true"].copy()
    X_validation = methy_b_validation_cluster.dropna().T

    """
    DMRs naming
    """
    dmrs = pannot.loc[cluster_map.index]
    dmrs = pd.concat(
        [
            dmrs.groupby("cluster")["Start_hg38"].min(),
            dmrs.groupby("cluster")["End_hg38"].max(),
            dmrs.groupby("cluster")["CHR_hg38"].first(),
        ],
        axis=1,
    )
    dmrs["name"] = [
        f"DMR_{c.capitalize()}:{s:.0f}-{e:.0f}"
        for c, s, e in dmrs[["CHR_hg38", "Start_hg38", "End_hg38"]].values
    ]

    """
    Exported tables
    """
    fw = pd.read_excel("tables/LOO_threshold_scores.xlsx")
    coefs_top = pd.read_excel(f"tables/LOO_coefs_top.xlsx", index_col=0)
    top_coefs = pd.read_excel(f"tables/LOO_coefs_top_annotated.xlsx")

    """
    Feature weights scaling
    """
    fw_mean = fw.mean().drop(["thres", "recall", "precision"])
    # fw_mean = fw_mean.sample(frac=0.7)

    nrom_factor = fw_mean.abs().sum()

    fw_mean = fw_mean / nrom_factor
    train_thres = fw["thres"].mean() / nrom_factor

    samples = pcs.query("label != 'cfDNA'")

    """
    
    """
    def probes_cluster_undersample(df, top_dmr_n=5, dmr_cpg_n=2):
        # Read probe annotation
        pcluster_annot = MLiquidBiopsy.read_probe_annot().dropna(subset=["cluster"])
        pcluster_annot = pcluster_annot[pcluster_annot.index.isin(df.index)]

        # Subset to top DMR
        cannot = pcluster_annot[
            pcluster_annot["cluster"].isin(list(coefs_top.head(top_dmr_n).index))
        ]

        # Random downsample
        cannot = cannot.reset_index().groupby("cluster").sample(dmr_cpg_n).set_index("IlmnID")

        # Calculate DMR methylation
        df_cluster = df.groupby(cannot["cluster"]).mean()

        return df_cluster.T, cannot

    #
    SCORE_N_ITERATIONS = 10

    fw_subset = []

    for top_dmr in [8, 10, 20, 38]:
        for dmr_cpgs in [1, 2, 3]:
            for j in range(SCORE_N_ITERATIONS):
                LOG.info(f"N CpGs={dmr_cpgs}; Top DMRs={top_dmr}")
                X_subset, DMR_subset = probes_cluster_undersample(methy_b_, top_dmr, dmr_cpgs)

                fw_subset_iter = []
                for i in range(SCORE_N_ITERATIONS):
                    x_iter, y_iter = build_matrices(X_subset.loc[Y.index], Y, samples)
                    ftype_x_train = x_iter.reindex(index=y_iter.index)

                    clf = RidgeClassifier(alpha=1).fit(ftype_x_train, y_iter)
                    fw_subset_iter.append(
                        pd.Series(clf.coef_[0], index=x_iter.columns, name="coef")
                        .reset_index()
                        .assign(feature="DMR signature", iter=i)
                    )
                fw_subset_iter = pd.concat(fw_subset_iter, ignore_index=True)

                # Estimate thresholds
                fw_subset_iter_ci = []

                _, ax = plt.subplots(1, 1, figsize=(2, 2))

                for i in range(SCORE_N_ITERATIONS):
                    iter_weights = fw_subset_iter.query(
                        f"(feature == 'DMR signature') & (iter == {i})"
                    )
                    iter_weights = iter_weights.groupby("cluster")["coef"].mean().sort_values()

                    iter_scores = pd.concat(
                        [
                            methy_b_cluster.loc[iter_weights.index]
                                .T.dot(iter_weights)
                                .rename("score"),
                            samples["label"].apply(MLiquidBiopsy.sample_class).rename("y_true"),
                        ],
                        axis=1,
                    ).loc[samples.index]

                    _, t, r, p = MLiquidBiopsyPlot.recall_precision_curve(
                        iter_scores["y_true"],
                        iter_scores["score"],
                        color=CrispyPlot.PAL_DTRACE[0],
                        plot_legend=False,
                        plot_random=False,
                        plot_f1curves=i == 0,
                        ax=ax,
                    )

                    res = iter_weights.to_dict()
                    res["thres"] = t
                    res["recall"] = r
                    res["precision"] = p

                    fw_subset_iter_ci.append(res)

                plt.close("all")

                fw_subset_iter_ci = pd.DataFrame(fw_subset_iter_ci)

                fw_subset_iter_agg = fw_subset_iter_ci.agg([np.mean, np.std]).T
                fw_subset_iter_agg["dmr"] = dmrs.reindex(index=fw_subset_iter_agg.index, columns=["name"]).values

                metrics = ["thres", "recall", "precision"]
                order = list(fw_subset_iter_agg["mean"].abs().drop(metrics).sort_values(ascending=False).index) + metrics
                fw_subset_iter_agg = fw_subset_iter_agg.loc[order]

                fw_subset.append(
                    fw_subset_iter_agg.assign(n_cpgs=dmr_cpgs, top_dmrs=top_dmr).reset_index()
                )

    fw_subset = pd.concat(fw_subset)

    #
    fw_subset_metric = "recall"
    for fw_subset_metric in ["recall", "precision"]:
        for fw_subset_submetric in ["mean", "std"]:
            plot_df = fw_subset.query(f"index == '{fw_subset_metric}'")
            plot_df = pd.pivot_table(
                plot_df, index="n_cpgs", columns="top_dmrs", values=fw_subset_submetric, aggfunc=np.mean
            )

            _, ax = plt.subplots(1, 1, figsize=(2.5, 2.5))

            fig = sns.clustermap(
                plot_df.rename(columns=dmrs["name"]),
                cmap="binary",
                annot_kws={"size": 5},
                yticklabels=True,
                xticklabels=True,
                annot=True,
                col_cluster=False,
                row_cluster=False,
                cbar=False,
                dendrogram_ratio=0.1,
                colors_ratio=0.01,
                figsize=(2.5, 2.5),
                cbar_pos=None,
            )

            fig.ax_heatmap.set_ylabel("Randomly sample N CpG sites per DMR")
            fig.ax_heatmap.set_xlabel("Top N DMRs")
            fig.ax_heatmap.set_yticklabels(
                fig.ax_heatmap.get_yticklabels(), rotation=90, fontsize=6
            )
            fig.ax_heatmap.set_title(f"{fw_subset_submetric.capitalize()} {fw_subset_metric.capitalize()}")

            plt.savefig(
                f"reports/LOO_Signature_downsample_heatmap_{fw_subset_metric}_{fw_subset_submetric}.pdf",
                bbox_inches="tight",
                dpi=600,
            )
            plt.close("all")

    """
    Score iterations
    """
    samples = pcs.query("label != 'cfDNA'")

    TDP_DMR_N = 2

    SCORE_N_ITERATIONS = 1000
    fw_subset_iter = []
    for i in range(SCORE_N_ITERATIONS):
        LOG.info(f"Iterations: {i+1}")
        x_iter, y_iter = build_matrices(X, Y, samples)

        ftype_features = list(coefs_top.head(TDP_DMR_N).index)
        # ftype_features = list(fw_mean.index)
        ftype_x_train = x_iter.reindex(
            index=y_iter.index, columns=ftype_features
        )

        clf = RidgeClassifier(alpha=1).fit(ftype_x_train, y_iter)
        fw_subset_iter.append(
            pd.Series(clf.coef_[0], index=ftype_features, name="coef")
            .reset_index()
            .assign(feature="DMR signature", iter=i)
        )

    fw_subset_iter = pd.concat(fw_subset_iter, ignore_index=True)

    # Estimate thresholds
    fw_subset_iter_ci = []

    _, ax = plt.subplots(1, 1, figsize=(2, 2))

    for i in range(SCORE_N_ITERATIONS):
        iter_weights = fw_subset_iter.query(
            f"(feature == 'DMR signature') & (iter == {i})"
        )
        iter_weights = iter_weights.groupby("index")["coef"].mean().sort_values()

        iter_scores = pd.concat(
            [
                methy_b_cluster.loc[iter_weights.index]
                    .T.dot(iter_weights)
                    .rename("score"),
                samples["label"].apply(MLiquidBiopsy.sample_class).rename("y_true"),
            ],
            axis=1,
        ).loc[samples.index]

        _, t, r, p = MLiquidBiopsyPlot.recall_precision_curve(
            iter_scores["y_true"],
            iter_scores["score"],
            color=CrispyPlot.PAL_DTRACE[0],
            plot_legend=False,
            plot_random=False,
            plot_f1curves=i == 0,
            ax=ax,
        )

        res = iter_weights.to_dict()
        res["thres"] = t
        res["recall"] = r
        res["precision"] = p

        fw_subset_iter_ci.append(res)

    plt.savefig(
        f"reports/LOO_Signature_precision_recall_curves_ci_Top{TDP_DMR_N}.pdf",
        bbox_inches="tight",
        dpi=600,
    )
    plt.close("all")

    fw_subset_iter_ci = pd.DataFrame(fw_subset_iter_ci)

    fw_subset_iter_agg = fw_subset_iter_ci.agg([np.mean, np.std]).T
    fw_subset_iter_agg["dmr"] = dmrs.reindex(index=fw_subset_iter_agg.index, columns=["name"]).values

    metrics = ["thres", "recall", "precision"]
    order = list(fw_subset_iter_agg["mean"].abs().drop(metrics).sort_values(ascending=False).index) + metrics
    fw_subset_iter_agg = fw_subset_iter_agg.loc[order]

    fw_subset_iter_agg.to_excel(f"tables/LOO_DMRs_top{TDP_DMR_N}_coefficientss_and_classification_thres.xlsx")

    #
    plot_df = pd.concat([
        fw.mean().drop(metrics).rename(f"Top 38"),
        fw_subset_iter_ci.mean().drop(metrics).rename(f"Top {TDP_DMR_N}"),
    ], axis=1)

    _, ax = plt.subplots(1, 1, figsize=(2, 2))

    grid = GIPlot.gi_regression(
        "Top 38",
        f"Top {TDP_DMR_N}",
        plot_df,
        xlim=(-1.5, 1.5),
        ylim=(-1.5, 1.5),
        reg_line_color="black",
    )

    grid.ax_joint.plot([0, 1], [0, 1], transform=grid.ax_joint.transAxes, ls="--", c="k", lw=0.5, zorder=0)

    plt.savefig(
        f"reports/LOO_Signature_top{TDP_DMR_N}_coefficients_corr.pdf",
        bbox_inches="tight",
        dpi=600,
    )
    plt.close("all")

    """
    Score samples
    """
    X_scores = X[fw_mean.index].dot(fw_mean)
    X_validation_scores = X_validation[fw_mean.index].dot(fw_mean)

    """
    Plot
    """
    y_var = "chr8_Cluster409529"

    plot_df = pd.concat([
        pd.concat([X_scores, X_validation_scores]).rename("Score"),
        pd.concat([X[y_var], X_validation[y_var]]),
        pd.concat([ss_all["Type"], ss_all_validation["Type"]]),
        pd.concat([
            pd.Series(["Train&Test"] * X_scores.shape[0], index=X_scores.index),
            pd.Series(["Validation"] * X_validation_scores.shape[0], index=X_validation_scores.index),
        ]).rename("Dataset"),
    ], axis=1)

    pal = {
        "Other": CrispyPlot.PAL_DTRACE[0],
        "HCC": CrispyPlot.PAL_DTRACE[1],
        "Cancer": CrispyPlot.PAL_DTRACE[3],
        "Healthy": "#63BF55",
    }

    hue_order = ["Other", "HCC"]

    _, ax = plt.subplots(figsize=(2, 2), dpi=600)

    GIPlot.gi_tissue_plot(
        "Score",
        y_var,
        plot_df=plot_df.query("Dataset == 'Train&Test'"),
        hue="Type",
        hue_order=hue_order,
        plot_reg=False,
        annot=False,
        pal=pal,
        markers="o",
        ax=ax,
    )

    GIPlot.gi_tissue_plot(
        "Score",
        y_var,
        plot_df=plot_df.query("Dataset == 'Validation'"),
        hue="Type",
        hue_order=hue_order,
        plot_reg=False,
        annot=False,
        pal=pal,
        markers="x",
        ax=ax,
    )

    ax.axvline(
        train_thres,
        lw=0.5,
        ls="--",
        c=CrispyPlot.PAL_DTRACE[2],
        zorder=0,
    )
    # ax.axvspan(
    #     train_thres - train_thres_sd,
    #     train_thres + train_thres_sd,
    #     alpha=0.3,
    #     color=CrispyPlot.PAL_DTRACE[0],
    #     zorder=0,
    #     lw=0,
    # )

    ax.set_xlabel(f"DMR signature score")
    ax.set_ylabel(f"{y_var}\nDNA methylation (beta)")
    ax.set_title(f"Validation\nTissue")

    ax.grid(True, ls="-", lw=0.1, alpha=1.0, zorder=0)

    plt.savefig(
        f"reports/LOO_Signature_Score_{y_var}.pdf",
        bbox_inches="tight",
        transparent=True,
        dpi=600,
    )
    plt.close("all")
```
