## Supplementary File 1 for "DNA methylation fingerprint for the diagnosis and monitoring of hepatocellular carcinoma from tissue and liquid biopsies": BinaryTrainingValidation.py

```
import numpy as np
import pandas as pd
import seaborn as sns
import matplotlib.pyplot as plt
from crispy.LMModels import LModel, LMModels
from scipy.stats import pearsonr, spearmanr
from statsmodels.stats.multitest import multipletests
from statsmodels.stats.weightstats import ttest_ind
from LiquidBiopsy import LOG, two_vars_correlation
from crispy import CrispyPlot
from crispy.GIPlot import GIPlot
from crispy.Enrichment import Enrichment
from sklearn.preprocessing import StandardScaler
from upsetplot import from_contents
from upsetplot import plot as upsetplot
from matplotlib.lines import Line2D
from limix.qtl import scan
from limix.stats import linear_kinship
from BinaryTrainingPlot import BinaryTrainingPlot
from sklearn.metrics import precision_score, recall_score
from MLiquidBiopsy import MLiquidBiopsy, MLiquidBiopsyPlot
from sklearn.feature_selection import SequentialFeatureSelector
from sklearn.model_selection import LeaveOneOut


PAL_LABEL = pd.Series(
    sns.color_palette("Set1", n_colors=2).as_hex(), index=["Cirrhosis", "HCC"]
)

LABELS = [
    "Primary Tumor - Liver",
    "HCC + EtOH",
    "HCC + HCV",
    "cfDNA HCC and cirrhosis",
    "cfDNA cirrhosis only",
    "Cirrhosis + EtOH",
    "Cirrhosis + HCV",
    "Cirrhosis + HBV",
    "Cirrhosis + Other",
    "Cirrhosis + AATD",
    "AATD",
    "Liver obese",
    "Solid Tissue Normal - Liver",
    "Liver normal",
    "cfDNA",
    "cfDNA healthy pooled",
    "cfDNA healthy",
]

if __name__ == "__main__":
    """
    Imports
    """
    # Samplesheet
    methy_b, ss_all, pal_label, pal_type = MLiquidBiopsy.import_dataset(
        f"~/datasets/DNAMethylationPipeline_MergedSubset_methy_b.csv",
        subset_labels=LABELS,
    )
    ss_all = pd.read_excel(f"tables/LOO_samplesheet.xlsx", index_col=0)

    # Probe annotation information
    pannot = MLiquidBiopsy.read_probe_annot()

    # Dataset palette
    pal_dataset = ss_all.groupby("dataset_id")["dataset_color"].first()

    # Feature palette
    features = ["DMR signature"] + list(MLiquidBiopsy.TOP_CPGS)
    markers = ["o"] + ["x"] * len(list(MLiquidBiopsy.TOP_CPGS))
    linestyles = ["-"] + ["--"] * len(list(MLiquidBiopsy.TOP_CPGS))
    features_pal = BinaryTrainingPlot.palette_feature()

    """
    Feature reduction
    """
    # Drop NaNs
    methy_b = methy_b.dropna()
    pcs, dims, vexp = BinaryTrainingPlot.dim_red(
        methy_b.T, "methy", ss_all, pal_label, n_components=10
    )

    # Drop probes not in hg38 lift annotation
    methy_b_ = methy_b.loc[methy_b.index.isin(pannot.index)].copy()

    # Drop sex chromosome
    psex = MLiquidBiopsy.probes_sex_chr()
    methy_b_ = methy_b_.drop(index=psex, errors="ignore")

    # Cluster methylation
    methy_b_cluster, cluster_map = MLiquidBiopsy.probes_cluster(methy_b_, nmin=3)
    cluster_map_cpg = cluster_map.reset_index().groupby("cluster")["IlmnID"].agg(set)

    BinaryTrainingPlot.plot_clusters_length(pannot, methy_b_, methy_b_cluster)

    """
    DMRs
    """
    dmrs = pannot.loc[cluster_map.index]
    dmrs = pd.concat(
        [
            dmrs.groupby("cluster")["Start_hg38"].min(),
            dmrs.groupby("cluster")["End_hg38"].max(),
            dmrs.groupby("cluster")["CHR_hg38"].first(),
        ],
        axis=1,
    )
    dmrs["name"] = [
        f"DMR_{c.capitalize()}:{s:.0f}-{e:.0f}"
        for c, s, e in dmrs[["CHR_hg38", "Start_hg38", "End_hg38"]].values
    ]

    """
    Matrices
    """
    Y = ss_all["label"].apply(MLiquidBiopsy.sample_class)
    X = methy_b_cluster[Y.index].T

    """
    Validation set
    """
    ss_all_validation, methy_b_validation = MLiquidBiopsy.read_test_datasets("all")
    ss_all_validation["Type"] = ss_all_validation["y_true"].replace(
        {0: "Other", 1: "HCC"}
    )

    pal_val_dataset = ss_all_validation.groupby("dataset_id")["dataset_color"].first()

    methy_b_validation_cluster = methy_b_validation[
        methy_b_validation.index.isin(methy_b_.index)
    ]
    methy_b_validation_cluster, _ = MLiquidBiopsy.probes_cluster(
        methy_b_validation_cluster, nmin=3
    )

    Y_validation = ss_all_validation["y_true"].copy()
    X_validation = methy_b_validation_cluster.dropna().T

    """
    TCGA gene expression
    """
    ss_tcga = ss_all.dropna(subset=["barcode", "age_at_index", "gender"])
    ss_tcga["barcode_simple"] = ss_tcga["barcode"].apply(lambda v: v[:15]).values

    gexp = pd.read_csv(
        "~/datasets/TCGA_RNAseq/GSE62944_merged_expression_voom.tsv",
        sep="\t",
        index_col=0,
    )
    gexp = gexp.groupby([c[:15] for c in gexp], axis=1).mean()
    gexp = gexp.loc[:, gexp.columns.isin(set(ss_tcga["barcode_simple"]))]

    methy_b_tcga = methy_b.reindex(columns=ss_tcga.index).rename(
        columns=ss_tcga["barcode_simple"]
    )[gexp.columns]
    methy_b_cluster_tcga = methy_b_cluster.reindex(columns=ss_tcga.index).rename(
        columns=ss_tcga["barcode_simple"]
    )[gexp.columns]

    """
    Biomarkers
    """
    coefs_top = pd.read_excel(f"tables/LOO_coefs_top.xlsx", index_col=0)

    coefs_top_ftr = coefs_top.head(38)

    coefs_top_ftr["cpg_sites"] = (
        cluster_map_cpg.loc[coefs_top_ftr.index].apply(list).values
    )
    coefs_top_ftr = (
        coefs_top_ftr.explode("cpg_sites").reset_index().set_index("cpg_sites")
    )

    extra_covs = ss_all[["GlobalMethylation", "SamplePurityPC5"]]
    coefs_top_ftr = pd.concat(
        [
            coefs_top_ftr,
            MLiquidBiopsy.anova_cov(
                methy_b.loc[coefs_top_ftr.index].T,
                Y,
                ss_all,
                dataset_as_cov=False,
                extra_covs=extra_covs,
            ).add_prefix("TrainTest_"),
        ],
        axis=1,
    )

    coefs_top_ftr = pd.concat(
        [
            coefs_top_ftr,
            pannot.loc[coefs_top_ftr.index, MLiquidBiopsy.MANIFEST_COLS],
        ],
        axis=1,
    ).rename(columns=dict(index="cpg_cluster"))

    promoter_regions = ["TSS1500", "TSS200", "5'UTR", "1stExon"]
    coefs_top_ftr["is_promoter"] = [
        len(set(s.split(";")).intersection(promoter_regions)) > 0
        if str(s) != "nan"
        else False
        for s in coefs_top_ftr["UCSC_RefGene_Group"]
    ]

    coefs_top_ftr.to_excel(
        f"tables/LOO_Validation_CpGclusters_annotated.xlsx", index=False
    )

    """
    Methylation clustermaps
    """
    plot_df = pd.concat(
        [
            X.reindex(columns=coefs_top_ftr["cpg_cluster"].unique()),
            X_validation.reindex(columns=coefs_top_ftr["cpg_cluster"].unique()),
        ],
        axis=0,
    )

    row_colors = pd.concat([ss_all, ss_all_validation])[
        ["Type", "dataset_color"]
    ].replace(dict(Type=pal_type))
    row_colors = row_colors.rename(columns=dict(Type="HCC", dataset_color="Dataset"))
    row_colors["Train & Test"] = [
        CrispyPlot.PAL_DTRACE[3 if i in ss_all.index else 0] for i in row_colors.index
    ]

    col_colors = pd.DataFrame(
        {
            "Top Features": pd.Series(
                plot_df.columns.isin(coefs_top.head(38).index), index=plot_df.columns
            ).replace({True: CrispyPlot.PAL_DTRACE[2], False: "#FFFFFF"})
        }
    ).rename(index=dmrs["name"])

    fig = sns.clustermap(
        plot_df.rename(columns=dmrs["name"]),
        cmap="Spectral",
        center=0.5,
        annot_kws={"size": 5},
        yticklabels=False,
        xticklabels=True,
        col_colors=col_colors,
        row_colors=row_colors,
        dendrogram_ratio=0.1,
        colors_ratio=0.01,
        figsize=(13, 6),
        cbar_pos=(0.02, 0.90, 0.02, 0.15),
    )

    fig.ax_heatmap.set_xlabel("")
    fig.ax_heatmap.set_ylabel("Samples")
    fig.ax_heatmap.set_xticklabels(
        fig.ax_heatmap.get_xticklabels(), rotation=90, fontsize=5
    )
    fig.ax_cbar.set_title("Methylation\n(beta)")

    plt.savefig(
        f"reports/LOO_Validation_clustermap.pdf",
        bbox_inches="tight",
        dpi=600,
    )
    plt.close("all")

    #
    fig = sns.clustermap(
        plot_df[coefs_top.head(38).index].rename(columns=dmrs["name"]),
        cmap="Spectral",
        center=0.5,
        annot_kws={"size": 5},
        yticklabels=False,
        xticklabels=True,
        row_colors=row_colors,
        dendrogram_ratio=0.1,
        colors_ratio=0.035,
        figsize=(5, 6),
        cbar_pos=(0.05, 0.125, 0.02, 0.1),
    )

    fig.ax_heatmap.set_xlabel("")
    fig.ax_heatmap.set_ylabel("Samples")
    fig.ax_heatmap.set_xticklabels(
        fig.ax_heatmap.get_xticklabels(), rotation=90, fontsize=5
    )
    fig.ax_cbar.set_title("Methylation\n(beta)")

    plt.savefig(
        f"reports/LOO_Validation_clustermap_top_features_only.pdf",
        bbox_inches="tight",
        dpi=600,
    )
    plt.close("all")

    """
    Chromosome ideogram
    """
    plot_df = pd.concat(
        [
            pd.DataFrame(
                dict(
                    cpg=list(
                        coefs_top_ftr.query(
                            f"TrainTest_fdr < 0.01 & TrainTest_ANOVA_fdr < 0.01"
                        ).index
                    ),
                    db="DMR signature",
                )
            ),
            MLiquidBiopsy.independent_features(methy_b),
        ],
        ignore_index=True,
    )

    plot_df = plot_df.groupby("cpg").first().reset_index()
    plot_df = plot_df[plot_df["cpg"].isin(pannot.index)]

    plot_df = pd.concat(
        [
            plot_df,
            pannot.loc[plot_df["cpg"]][MLiquidBiopsy.MANIFEST_COLS].reset_index(
                drop=True
            ),
        ],
        axis=1,
    )

    plot_df = plot_df.rename(
        columns=dict(CHR_hg38="chrom", Start_hg38="start", End_hg38="end", cpg="name")
    )[["chrom", "start", "end", "name", "db"]]

    plot_df["colors"] = plot_df["db"].apply(MLiquidBiopsyPlot.sample_color)
    plot_df["db"] = pd.Categorical(
        plot_df["db"],
        ordered=True,
        categories=features,
    )
    plot_df = plot_df.sort_values("db", ascending=True).query("db == 'DMR signature'")

    plot_df["beta"] = coefs_top_ftr.loc[plot_df["name"], "TrainTest_beta"].values
    plot_df["db"] = plot_df["beta"].apply(lambda v: "Hyper" if v > 0 else "Hypo").values
    plot_df["colors"] = (
        plot_df["beta"].apply(lambda v: "#2980D9" if v > 0 else "#C0392B").values
    )

    for n, chr_list in [
        ("part1", [f"chr{i}" for i in range(1, 12)]),
        ("part2", [f"chr{i}" for i in range(12, 23)]),
    ]:
        ax = MLiquidBiopsyPlot.chromosome_ideogram(
            plot_df,
            chrom_height=0.3,
            gene_height=0.6,
            gene_padding=0,
            chromosome_list=chr_list,
            figsize=(3, 2),
        )

        ax.set_xlabel("Genomic position")
        ax.set_ylabel("Chromosome")

        hue_order = ["Hyper", "Hypo"]
        ax.legend(
            [
                Line2D([0], [0], color="#2980D9" if s == "Hyper" else "#C0392B", lw=4)
                for s in hue_order
            ],
            hue_order,
            loc=4,
            frameon=False,
            prop=dict(size=6),
            title="Methylation",
        ).get_title().set_fontsize("6")

        plt.savefig(
            f"reports/LOO_Validation_chromosome_ideogram_{n}.pdf",
            bbox_inches="tight",
            dpi=600,
        )
        plt.close("all")

    #
    chr_signatures = (
        pannot.loc[cluster_map.index]
        .reset_index()
        .groupby("CHR_hg38")["IlmnID"]
        .agg(set)
    )

    chr_cpgsites_enr = pd.DataFrame(
        {
            c: Enrichment.hypergeom_test(
                chr_signatures[c],
                set(cluster_map.index),
                set(
                    coefs_top_ftr.query(
                        f"TrainTest_fdr < 0.01 & TrainTest_ANOVA_fdr < 0.01"
                    ).index
                ),
            )
            for c in set(coefs_top_ftr["CHR_hg38"])
        },
        index=["pval", "intersection"],
    ).T.sort_values("pval")
    chr_cpgsites_enr["percent"] = (
        chr_cpgsites_enr["intersection"] / coefs_top_ftr.shape[0] * 100
    )
    chr_cpgsites_enr["fdr"] = multipletests(chr_cpgsites_enr["pval"], method="fdr_bh")[
        1
    ]

    chr_cpgsites_enr.to_excel(f"tables/LOO_Validation_CpGclusters_enrichment.xlsx")

    """
    Differential gexp LMM
    """
    lmm_m = (
        pd.concat(
            [
                ss_tcga["gender_predicted"]
                .str.get_dummies()["F"]
                .rename("Female")
                .rename(index=ss_tcga["barcode_simple"]),
                ss_tcga[
                    [
                        "age_at_index",
                        "age_predicted_horvath",
                        "age_predicted_hannum",
                        "GlobalMethylation",
                        "SamplePurityPC5",
                    ]
                ].rename(index=ss_tcga["barcode_simple"]),
            ],
            axis=1,
        )
        .rename(index=ss_tcga["barcode_simple"])
        .dropna()
    )
    lmm_m["intercept"] = 1

    samples = set.intersection(
        set(ss_tcga["barcode_simple"]), set(methy_b_tcga), set(gexp), set(lmm_m.index)
    )

    lmm_y = methy_b_cluster_tcga.loc[
        coefs_top_ftr["cpg_cluster"].unique(), samples
    ].T.copy()

    lmm_x = gexp[samples].T.copy()
    lmm_x = LMModels.transform_matrix(lmm_x, t_type="scale")

    lmm_k = pd.DataFrame(
        linear_kinship(gexp[samples].T.values), index=lmm_y.index, columns=lmm_y.index
    )

    lmm_m = lmm_m.loc[samples]

    lmm_cluster_assoc = []
    for c in lmm_y:
        lmm_cg = scan(
            lmm_x, lmm_y[c], K=lmm_k.values, M=lmm_m, lik="normal", verbose=False
        )

        cols_rename = dict(
            effect_name="gene", pv20="pval", effsize="beta", effsize_se="beta_se"
        )

        effect_sizes = lmm_cg.effsizes["h2"].query("effect_type == 'candidate'")
        effect_sizes = effect_sizes.set_index("test").drop(
            columns=["trait", "effect_type"]
        )

        pvalues = lmm_cg.stats["pv20"]

        lmm_res = pd.concat([effect_sizes, pvalues], axis=1, sort=False)
        lmm_res = (
            lmm_res.reset_index(drop=True).rename(columns=cols_rename).set_index("gene")
        )
        lmm_res["fdr"] = multipletests(lmm_res["pval"], method="fdr_bh")[1]
        lmm_res["cpg_cluster"] = c

        lmm_cluster_assoc.append(lmm_res)

    lmm_cluster_assoc = pd.concat(lmm_cluster_assoc).sort_values("pval")

    lmm_cluster_assoc["dmr"] = (
        lmm_cluster_assoc["cpg_cluster"].replace(dmrs.loc[lmm_y.columns, "name"]).values
    )

    lmm_cluster_assoc.query("fdr < 0.1").to_excel(
        "tables/LOO_Validation_Gexp_associations.xlsx"
    )

    pd.Series(list(set(lmm_cluster_assoc.query("fdr < 0.1").index))).to_csv(
        f"reports/LOO_Validation_Signif_DMR_Gexp_GeneList.txt",
        index=False,
        header=False,
    )

    #
    c, g = "chr7_Cluster375405", "HOXA7"

    for c, g in [
        ("chr7_Cluster375405", "HOXA6"),
        ("chr7_Cluster375405", "HOXA3"),
        ("chr7_Cluster375405", "HOXA5"),
        ("chr7_Cluster375405", "HOXA7"),
        ("chr7_Cluster375405", "HOXA4"),
        ("chr10_Cluster63685", "ZNF518B"),
        ("chr5_Cluster343774", "ZNF502"),
        ("chr7_Cluster371128", "ZNF132"),
    ]:
        plot_df = (
            pd.concat(
                [
                    gexp.loc[g],
                    methy_b_cluster_tcga.loc[c],
                    ss_tcga.set_index("barcode_simple"),
                ],
                axis=1,
            )
            .loc[samples]
            .dropna(subset=[c, g])
        )

        grid = GIPlot.gi_regression(c, g, plot_df, hue="Type", palette=pal_type)
        grid.set_axis_labels(
            f"{dmrs.loc[c, 'name']}\nDNA methylation (beta)",
            f"{g}\nGene expression (RNA-Seq voom)",
        )
        plt.savefig(
            f"reports/LOO_Validation_regression_{c}_{g}.pdf",
            bbox_inches="tight",
            dpi=600,
        )
        plt.close("all")
```
