## Supplementary File 1 for "DNA methylation fingerprint for the diagnosis and monitoring of hepatocellular carcinoma from tissue and liquid biopsies": Deconvolution.py

```
import numpy as np
import palettable
import pandas as pd
import seaborn as sns
import matplotlib.pyplot as plt
from umap import UMAP
from crispy import CrispyPlot
from natsort import natsorted
from crispy.GIPlot import GIPlot
from sklearn.svm import LinearSVC
from matplotlib.lines import Line2D
from sklearn.preprocessing import StandardScaler
from sklearn.linear_model import LinearRegression, Ridge, Lasso
from LiquidBiopsy import LOG, manifest_label
from sklearn.model_selection import StratifiedKFold, StratifiedShuffleSplit
from statsmodels.stats.multitest import multipletests
from sklearn.feature_selection import VarianceThreshold, f_classif
from sklearn.metrics import (
    recall_score,
    precision_score,
    r2_score,
    matthews_corrcoef,
    average_precision_score,
)


"""
Imports
"""

# Samplesheet
ss = pd.read_csv(f"~/datasets/DNAMethylationPipeline_samplesheet.csv.gz", index_col=0)
ss = pd.concat(
    [
        ss,
        pd.DataFrame(
            {
                i: manifest_label(r["sample_type"], r["tissue_or_organ_of_origin"])
                for i, r in ss.iterrows()
            }
        ).T,
    ],
    axis=1,
)
ss["stage"] = ss["ajcc_pathologic_stage"].apply(
    lambda v: v[:-1] if str(v) != "nan" and v.endswith(("A", "B", "C")) else v
)

label_pal = ss.groupby("label")["color"].first()

# Methylation beta
methy_b_450k = pd.read_csv(f"~/datasets/DNAMethylationPipeline_methy_b.csv")
methy_b_epic = pd.read_csv(f"~/datasets/DNAMethylationPipeline_methy_b_EPIC.csv")

# Probe annotation
probe_annot = pd.read_csv(f"~/datasets/DNAMethylationPipeline_methy_annotation.csv.gz")

# Methylation atlas - GSE122126
methy_atlas = pd.read_csv(
    f"~/datasets/Methylation_450k_GSE122126/full_atlas.csv.gz", index_col=0
)
methy_atlas_small = pd.read_csv(
    f"~/datasets/Methylation_450k_GSE122126/reference_atlas.txt", index_col=0
)

# cfDNA samples classification
cfdna_types = pd.read_csv(
    f"~/datasets/Methylation_450k_GSE122126/cfdna_samples_classification.tsv",
    index_col=1,
    sep="\t",
)

cfdna_pal = cfdna_types["type"].value_counts()
cfdna_pal = pd.Series(
    sns.color_palette("tab10").as_hex()[:len(cfdna_pal)],
    index=cfdna_pal.index,
)


"""
Top features
"""

top_features = pd.read_csv(f"reports/TopFeatures_Coef.csv", index_col=0)
top_features = top_features[top_features.index.isin(methy_b_epic.index)]

umap = pd.read_csv("reports/TopFeatures_UMAP.csv", index_col=0)


"""
DNA methylation reference atlas
"""

hcc_stage = ss.query("label == 'Primary Tumor - Liver'").reset_index().groupby("stage")["filename"].agg(set).to_dict()
tcga_stage = {f"HCC {k}": methy_b_450k.reindex(columns=hcc_stage[k]).mean(1) for k in hcc_stage}
tcga_stage["HCC"] = methy_b_450k.reindex(columns=ss.query("label == 'Primary Tumor - Liver'").index).mean(1)

tcga_stage["Normal Liver"] = methy_b_450k.reindex(
    columns=ss.query("label == 'Solid Tissue Normal - Liver'").index
).mean(1)
tcga_stage["Normal Colon"] = methy_b_450k.reindex(
    columns=ss.query("label == 'Solid Tissue Normal - Colon'").index
).mean(1)
tcga_stage["Normal Breast"] = methy_b_450k.reindex(
    columns=ss.query("(sample_type == 'Solid Tissue Normal') & (tissue_or_organ_of_origin == 'Breast')").index
).mean(1)

tcga_stage = pd.DataFrame(tcga_stage)

ref_matrix = pd.concat([
    tcga_stage,
    methy_atlas,
], axis=1)

ref_matrix = ref_matrix.fillna(ref_matrix.mean())


"""
DNA methylation signal deconvolution
"""

y_ss = ss.query("label == 'cfDNA'")

top = top_features.loc[
    top_features["coef"].abs().sort_values(ascending=False).index
].head(30)

y = (
    pd.concat(
        [
            methy_b_450k[set(methy_b_450k).intersection(y_ss.index)],
            methy_b_epic[set(methy_b_epic).intersection(y_ss.index)],
        ],
        axis=1,
    )
    .reindex(top.index)
    .dropna()
)

# Reference matrix X
x = ref_matrix.loc[y.index, [
    "Normal Liver", "Normal Breast", "Normal Colon",
    "Hepatocytes", "Colon_epithelial_cells", "Erythrocyte_progenitors", "Monocytes_EPIC", "Breast",
    "HCC Stage I", "HCC Stage II", "HCC Stage III", "HCC Stage IV",
    # "HCC",
]].copy()

g = sns.clustermap(x, cmap="YlGnBu", figsize=(4, 4))
plt.savefig(f"reports/Deconvolution_reference_matrix.pdf", bbox_inches="tight", dpi=600)
plt.close("all")

LOG.info(f"Number of probes and reference maps: {x.shape}")

# Linear regressions
lm_coefs = {}
for s in y.columns:
    s_lm = LinearRegression(positive=True, normalize=True).fit(x, y[s])
    s_coef = pd.Series(s_lm.coef_, index=x.columns)
    lm_coefs[s] = s_coef
lm_coefs = pd.DataFrame(lm_coefs)

# Plot features heatmap
plot_df = lm_coefs.copy().rename(columns=ss["sample_type"])

col_colors = cfdna_pal.loc[cfdna_types.loc[plot_df.columns]["type"]]
col_colors.index = plot_df.columns

g = sns.clustermap(plot_df, cmap="YlGnBu", figsize=(15, 5), col_colors=col_colors, annot=True, fmt=".2f", annot_kws=dict(size=5))

g.ax_col_dendrogram.legend(
    [Line2D([0], [0], color=cfdna_pal[i], lw=4) for i in cfdna_pal.index],
    list(cfdna_pal.index),
    frameon=False,
    loc="center left",
    bbox_to_anchor=(1, 0.9),
)

plt.savefig(f"reports/Deconvolution_clustermap.pdf", bbox_inches="tight", dpi=600)
plt.close("all")
```
