## Supplementary File 1 for "DNA methylation fingerprint for the diagnosis and monitoring of hepatocellular carcinoma from tissue and liquid biopsies": DifferentialMethylationMergedSubset.R

```
### Imports
library(minfi)
library(minfiData)
library(sva)
library(maxprobes)
library(doParallel)
library(bumphunter)

### Setup
setwd("~/jupyter_notebooks/LiquidBiopsy/")
registerDoParallel(cores = 20)

### Read IDAT files
# TCGA HCC Normals - 450k
RGSet_HCC_normals <- read.metharray.exp("~/datasets/Methylation_450k_TCGA_LIHC_normals/idats/")

# GSE122126 - EPIC
RGSet_GSE122126 <- read.metharray.exp("~/datasets/Methylation_450k_GSE122126/idats_epic/", force = T)
RGSet <- combineArrays(RGSet_GSE122126, RGSet_HCC_normals, outType = "IlluminaHumanMethylation450k", verbose = T)
colData(RGSet) <- rbind(colData(RGSet_GSE122126), colData(RGSet_HCC_normals))

# cfDNA - 450k
RGSet_cfdna <- read.metharray.exp("~/datasets/Methylation_450k_GSE129374/idats/")
RGSet <- combineArrays(RGSet_cfdna, RGSet, outType = "IlluminaHumanMethylation450k", verbose = T)
colData(RGSet) <- rbind(colData(RGSet_cfdna), colData(RGSet_GSE122126), colData(RGSet_HCC_normals))

# GSE122126 - 450k
RGSet_GSE <- read.metharray.exp("~/datasets/Methylation_450k_GSE122126/idats_450k/")
RGSet <- combineArrays(RGSet_GSE, RGSet, outType = "IlluminaHumanMethylation450k", verbose = T)
colData(RGSet) <- rbind(colData(RGSet_GSE), colData(RGSet_cfdna), colData(RGSet_GSE122126), colData(RGSet_HCC_normals))

# TCGA HCC - 450k
RGSet_HCC <- read.metharray.exp("~/datasets/Methylation_450k_TCGA_LIHC/idats/")
RGSet <- combineArrays(RGSet_HCC, RGSet, outType = "IlluminaHumanMethylation450k", verbose = T)
colData(RGSet) <- rbind(colData(RGSet_HCC), colData(RGSet_GSE), colData(RGSet_cfdna), colData(RGSet_GSE122126), colData(RGSet_HCC_normals))

# Obese liver - 450k
RGSet_obese <- read.metharray.exp("~/datasets/Methylation_450k_GSE61446/idats/")
RGSet <- combineArrays(RGSet_obese, RGSet, outType = "IlluminaHumanMethylation450k", verbose = T)
colData(RGSet) <- rbind(colData(RGSet_obese), colData(RGSet_HCC), colData(RGSet_GSE), colData(RGSet_cfdna), colData(RGSet_GSE122126), colData(RGSet_HCC_normals))

# Cirrhotic liver - 450k
RGSet_GSE60753 <- read.metharray.exp("~/datasets/Methylation_450k_GSE60753/idats/")
RGSet <- combineArrays(RGSet_GSE60753, RGSet, outType = "IlluminaHumanMethylation450k", verbose = T)
colData(RGSet) <- rbind(colData(RGSet_GSE60753), colData(RGSet_obese), colData(RGSet_HCC), colData(RGSet_GSE), colData(RGSet_cfdna), colData(RGSet_GSE122126), colData(RGSet_HCC_normals))

# GSE119100 - EPIC
RGSet_GSE119100 <- read.metharray.exp("~/datasets/Methylation_450k_GSE119100/idats/", force = T)
RGSet <- combineArrays(RGSet_GSE119100, RGSet, outType = "IlluminaHumanMethylation450k", verbose = T)
colData(RGSet) <- rbind(colData(RGSet_GSE119100), colData(RGSet_GSE60753), colData(RGSet_obese), colData(RGSet_HCC), colData(RGSet_GSE), colData(RGSet_cfdna), colData(RGSet_GSE122126), colData(RGSet_HCC_normals))

RGSet

### Preprocess
MSet <- preprocessFunnorm(RGSet, ratioConvert=F)
# MSet <- preprocessQuantile(RGSet, removeBadSamples=T)

### QC-plot
qc <- getQC(MSet)
MSet <- addQC(MSet, qc)

pdf("~/datasets/DNAMethylationPipelineQCReport_MergedSubset_plotQC.pdf")
plotQC(qc)
dev.off()

### Sex prediction
predictedSex <- getSex(MSet, cutoff = -2)
MSet <- addSex(MSet, predictedSex)

pdf("~/datasets/DNAMethylationPipelineQCReport_MergedSubset_Gender.pdf")
plotSex(MSet)
dev.off()

### SNPs
MSet <- addSnpInfo(MSet)
MSet <- dropLociWithSnps(MSet, snps=c("SBE","CpG"), maf = 0.01)

### Remove cross-reactive probes: devtools::install_github("markgene/maxprobes")
MSet <- maxprobes::dropXreactiveLoci(MSet)

### Export
RSet <- ratioConvert(MSet, what = "beta", keepCN = F, offset = 100)
GRset <- mapToGenome(RSet)

methy_b <- getBeta(MSet)
write.table(round(methy_b, 5), "~/datasets/DNAMethylationPipeline_MergedSubset_methy_b.csv", sep = ",", quote = F)

methy_annot <- getAnnotation(MSet)
write.table(methy_annot, "~/datasets/DNAMethylationPipeline_methy_MergedSubset_annotation.csv", sep = ",", quote = F)
```
