## Supplementary File 1 for "DNA methylation fingerprint for the diagnosis and monitoring of hepatocellular carcinoma from tissue and liquid biopsies": DNAMethylationPipeline.R

```
### Imports
library(minfi)
library(minfiData)
library(sva)
library(maxprobes)
library(doParallel)
library(bumphunter)

### Setup
setwd("~/jupyter_notebooks/LiquidBiopsy/")
registerDoParallel(cores = 20)

### Read IDAT files
# cfDNA
RGSet_cfdna <- read.metharray.exp("~/datasets/Methylation_450k_GSE129374/idats/")

# GSE122126
RGSet_GSE <- read.metharray.exp("~/datasets/Methylation_450k_GSE122126/idats_450k/")

# TCGA
RGSet_tcga <- read.metharray.exp("~/datasets/Methylation_450k_TCGA_normals/idats/")

# Obese liver
RGSet_obese <- read.metharray.exp("~/datasets/Methylation_450k_GSE61446/idats/")

# Cirrhotic liver
RGSet_GSE60753 <- read.metharray.exp("~/datasets/Methylation_450k_GSE60753/idats/")

# Whole-blood
redf <- "~/datasets/Methylation_450k_GSE40279/HannumpaperSamples_noRS/Hannum_Red_Channel_noRS_paperSamples.csv"
greenf <-  "~/datasets/Methylation_450k_GSE40279/HannumpaperSamples_noRS/Hannum_Green_Channel_noRS_paperSamples.csv"

redCh <- read.csv(redf, header = T, row.names=1, check.names = F)
redChMat <- as.matrix(redCh)

greenCh <- read.csv(greenf, header = T, row.names = 1, check.names = F)
greenChMat <- as.matrix(greenCh)

RGSet_blood <- RGChannelSet(
  Green=greenChMat,
  Red=redChMat,
  annotation=c(array="IlluminaHumanMethylation450k", annotation="ilmn12.hg19")
)

### Merge datasets
RGSet <- combineArrays(RGSet_tcga, RGSet_cfdna, outType = "IlluminaHumanMethylation450k", verbose = T)
colData(RGSet) <- rbind(colData(RGSet_tcga), colData(RGSet_cfdna))

RGSet <- combineArrays(RGSet_GSE, RGSet, outType = "IlluminaHumanMethylation450k", verbose = T)
colData(RGSet) <- rbind(colData(RGSet_GSE), colData(RGSet_tcga), colData(RGSet_cfdna))

RGSet <- combineArrays(RGSet_obese, RGSet, outType = "IlluminaHumanMethylation450k", verbose = T)
colData(RGSet) <- rbind(colData(RGSet_obese), colData(RGSet_GSE), colData(RGSet_tcga), colData(RGSet_cfdna))

RGSet <- combineArrays(RGSet_blood, RGSet, outType = "IlluminaHumanMethylation450k", verbose = T)
colData(RGSet) <- rbind(colData(RGSet_blood), colData(RGSet_obese), colData(RGSet_GSE), colData(RGSet_tcga), colData(RGSet_cfdna))

RGSet <- combineArrays(RGSet_GSE60753, RGSet, outType = "IlluminaHumanMethylation450k", verbose = T)
colData(RGSet) <- rbind(colData(RGSet_GSE60753), colData(RGSet_blood), colData(RGSet_obese), colData(RGSet_GSE), colData(RGSet_tcga), colData(RGSet_cfdna))

RGSet

### Preprocess
MSet <- preprocessFunnorm(RGSet, ratioConvert=F)

### QC-plot
qc <- getQC(MSet)
MSet <- addQC(MSet, qc)

pdf("~/datasets/DNAMethylationPipelineQCReport_plotQC.pdf")
plotQC(qc)
dev.off()

### Sex prediction
predictedSex <- getSex(MSet, cutoff = -2)
MSet <- addSex(MSet, predictedSex)

pdf("~/datasets/DNAMethylationPipelineQCReport_Gender.pdf")
plotSex(MSet)
dev.off()

### SNPs
MSet <- addSnpInfo(MSet)
MSet <- dropLociWithSnps(MSet, snps=c("SBE","CpG"), maf = 0.01)

### Remove cross-reactive probes: devtools::install_github("markgene/maxprobes")
MSet <- maxprobes::dropXreactiveLoci(MSet)

### Export
RSet <- ratioConvert(MSet, what = "beta", keepCN = F, offset = 100)
GRset <- mapToGenome(RSet)

methy_b <- getBeta(GRset)
write.table(round(methy_b, 5), "~/datasets/DNAMethylationPipeline_methy_b.csv", sep = ",", quote = F)

methy_annot <- getAnnotation(RSet)
write.table(methy_annot, "~/datasets/DNAMethylationPipeline_methy_annotation.csv", sep = ",", quote = F)
```
