## Supplementary File 1 for "DNA methylation fingerprint for the diagnosis and monitoring of hepatocellular carcinoma from tissue and liquid biopsies": DNAMethylationPipelineEpic.R

```
### Imports
library(minfi)
library(minfiData)
library(sva)
library(maxprobes)
library(doParallel)
library(bumphunter)

### Setup
setwd("~/jupyter_notebooks/LiquidBiopsy/")
registerDoParallel(cores = 20)

### Read IDAT files
# GSE122126
RGSet_GSE122126 <- read.metharray.exp("~/datasets/Methylation_450k_GSE122126/idats_epic/", force = T)

# GSE122126
RGSet_GSE119100 <- read.metharray.exp("~/datasets/Methylation_450k_GSE119100/idats/", force = T)

### Merge datasets
RGSet <- combineArrays(RGSet_GSE122126, RGSet_GSE119100, outType = "IlluminaHumanMethylationEPIC", verbose = T)
colData(RGSet) <- rbind(colData(RGSet_GSE122126), colData(RGSet_GSE119100))

### Preprocess
MSet <- preprocessFunnorm(RGSet, ratioConvert = F)

### QC-plot
qc <- getQC(MSet)
MSet <- addQC(MSet, qc)

pdf("~/datasets/DNAMethylationPipelineQCReport_plotQC_EPIC.pdf")
plotQC(qc)
dev.off()

### Sex prediction
predictedSex <- getSex(MSet, cutoff = -2)
MSet <- addSex(MSet, predictedSex)

pdf("~/datasets/DNAMethylationPipelineQCReport_Gender_EPIC.pdf")
plotSex(MSet)
dev.off()

### SNPs
MSet <- addSnpInfo(MSet)
MSet <- dropLociWithSnps(MSet, snps=c("SBE","CpG"), maf = 0.01)

### Remove cross-reactive probes: devtools::install_github("markgene/maxprobes")
MSet <- maxprobes::dropXreactiveLoci(MSet)

### Export
RSet <- ratioConvert(MSet, what = "beta", keepCN = F, offset = 100)
GRset <- mapToGenome(RSet)

methy_b <- getBeta(GRset)
write.table(round(methy_b, 5), "~/datasets/DNAMethylationPipeline_methy_b_EPIC.csv", sep = ",", quote = F)

methy_annot <- getAnnotation(RSet)
write.table(methy_annot, "~/datasets/DNAMethylationPipeline_methy_annotation_EPIC.csv", sep = ",", quote = F)
```
