## Supplementary File 1 for "DNA methylation fingerprint for the diagnosis and monitoring of hepatocellular carcinoma from tissue and liquid biopsies": MLiquidBiopsy.py

```
import os
import numpy as np
import pandas as pd
import seaborn as sns
import matplotlib.pyplot as plt
from crispy.LMModels import LModel, LMModels
from scipy.cluster import hierarchy
from scipy.stats import spearmanr
from umap import UMAP
from LiquidBiopsy import LOG
from limix.qtl import scan
from crispy import CrispyPlot
from crispy.GIPlot import GIPlot
from matplotlib.lines import Line2D
from sklearn.decomposition import PCA
from upsetplot import from_contents
from sklearn.svm import LinearSVC, SVC
from upsetplot import plot as upsetplot
from LiquidBiopsy import specificity_score
from more_itertools import unique_everseen
from sklearn.preprocessing import StandardScaler
from matplotlib.collections import BrokenBarHCollection
from sklearn.ensemble import RandomForestClassifier
from sklearn.utils.multiclass import type_of_target
from sklearn.feature_selection import f_classif, chi2
from statsmodels.stats.multitest import multipletests
from sklearn.metrics import (
    recall_score,
    precision_score,
    matthews_corrcoef,
    average_precision_score,
    precision_recall_curve,
    confusion_matrix,
    accuracy_score,
    balanced_accuracy_score,
)
from sklearn.linear_model import (
    LinearRegression,
    PassiveAggressiveClassifier,
    LogisticRegressionCV,
    RidgeClassifierCV,
    Perceptron,
    SGDClassifier,
    LogisticRegression,
)


# TSS1500, TSS200, 5′ UTR, and 1st exon

Top_CpGs_Hlady = {"cg04645914", "cg06215569", "cg07610777", "cg23663760"}

Top_CpGs_Xu_diagnosis = {
    "cg10428836",
    "cg26668608",
    "cg25754195",
    "cg05205842",
    "cg11606215",
    "cg24067911",
    "cg18196829",
    "cg23211949",
    "cg17213048",
    "cg25459300",
}

Top_CpGs_Xu_prognosis = {
    "cg23461741",
    "cg06482904",
    "cg25574765",
    "cg07459019",
    "cg20490031",
    "cg01643250",
    "cg11397370",
    "cg11825899",
}

Top_CpGs_villanueva = {
    "cg00043788",
    "cg00586537",
    "cg00696540",
    "cg01227537",
    "cg02318629",
    "cg02571816",
    "cg03578886",
    "cg03732762",
    "cg03920233",
    "cg04274978",
    "cg04321866",
    "cg04961553",
    "cg05412664",
    "cg06521280",
    "cg06671706",
    "cg07732116",
    "cg08379212",
    "cg09321747",
    "cg09472203",
    "cg10481660",
    "cg12881557",
    "cg13414212",
    "cg15019790",
    "cg15065139",
    "cg16113530",
    "cg17157630",
    "cg17179589",
    "cg18703913",
    "cg21085535",
    "cg22447539",
    "cg22639895",
    "cg23083315",
    "cg23624808",
    "cg23664775",
    "cg26527638",
    "cg26793648",
}

Top_CpGs_Cheng_et_al = {
    "cg23565942",
    "cg21908638",
    "cg11223367",
    "cg03509671",
    "cg05569109",
    "cg11481534",
}

Top_CpGs_patent_KR102103885B1 = {"cg12137206", "cg03792768"}

Top_CpGs_patent_US2019300965A1 = {
    "cg07360250",
    "cg08550839",
    "cg13499300",
    "ch.7.135065R",
    "cg14054357",
    "cg10673833",
}

Top_CpGs_patent_US2020263256A1 = {
    "cg10673833",
    "cg11225410",
    "cg00338116",
    "cg00552226",
    "cg24496475",
    "cg05414338",
    "cg12041340",
    "cg08343881",
    "cg03431741",
    "cg17126142",
    "cg18004756",
    "cg26205771",
    "cg08436738",
    "cg01604601",
    "cg27252696",
    "cg24917945",
    "cg11252953",
    "cg03550506",
    "cg06903569",
    "cg12865837",
    "cg18440897",
    "cg19255783",
    "cg20634573",
}

Top_CpGs_patent_CN110603329A = {
    "cg10428836",
    "cg26668608",
    "cg25754195",
    "cg05205842",
    "cg11606215",
    "cg24067911",
    "cg18196829",
    "cg23211949",
    "cg17213048",
    "cg25459300",
}

Top_CpGs_patent_US20180216195A1 = {"cg21790626"}

Top_CpGs_patent_JP2018508228A_1 = {
    "cg20468939",
    "cg24790419",
    "cg26836479",
    "cg16911583",
    "cg15139596",
    "cg16927606",
    "cg12967050",
    "cg21122474",
    "cg06064964",
    "cg11779113",
    "cg12042264",
    "cg27377213",
    "cg26680502",
    "cg12504877",
    "cg21913888",
    "cg26683005",
    "cg24166457",
    "cg27141915",
    "cg17122157",
    "cg09844573",
    "cg03087897",
    "cg2476505",
    "cg17126555",
    "cg13911392",
    "cg18901104",
    "cg25982880",
    "cg15797834",
    "cg27125093",
    "cg17518965",
    "cg20695297",
    "cg04858553",
    "cg09419005",
    "cg25490145",
    "cg11252953",
    "cg18456621",
    "cg07058988",
    "cg17864646",
    "cg06153925",
    "cg27410601",
    "cg03297901",
    "cg06853339",
    "cg12900649",
    "cg27219182",
    "cg15759721",
    "cg27023597",
    "cg02782634",
    "cg18942579",
    "cg01409343",
    "cg10530767",
    "cg26112797",
    "cg00253248",
    "cg0172227",
    "cg22589778",
    "cg07137244",
    "cg04147906",
    "cg23878564",
    "cg07860918",
    "cg00206490",
    "cg07644807",
    "cg00558804",
    "cg05304979",
    "cg27598656",
    "cg03549146",
    "cg22190721",
    "cg01660934",
    "cg02358862",
    "cg23093496",
    "cg07641284",
    "cg01681367",
    "cg26769927",
    "cg08480068",
    "cg02914427",
    "cg03653601",
    "cg01990910",
    "cg00933696",
    "cg09866569",
    "cg20357538",
    "cg22460896",
    "cg07116712",
    "cg10186131",
    "cg06380123",
    "cg18610205",
    "c g12353452",
    "cg10590292",
    "cg00037681",
    "cg05596756",
    "cg03569637",
    "cg02522196",
    "cg11655490",
    "cg19693177",
    "cg26363363",
    "cg21249754",
    "cg23147227",
    "cg01657186",
    "cg23764129",
    "cg04514998",
    "cg07332880",
    "cg16061668",
    "cg25574765",
    "cg14088196",
    "cg03758697",
    "cg05398700",
    "cg14058476",
    "cg18158859",
    "cg19300307",
    "cg18842353",
    "cg10732611",
    "cg24480810",
    "cg02053964",
    "cg25927251",
    "cg25954028",
    "cg14642045",
    "cg2465921",
    "cg18215449",
    "cg16402452",
    "cg21376733",
    "cg16509569",
    "cg08075204",
    "cg14556909",
    "cg07119472",
    "cg14999168",
    "cg09399878",
    "cg02874908",
    "cg10542975",
    "cg15698795",
    "cg11791526",
    "cg00862408",
    "cg16260696",
    "cg00220455",
    "cg20826709",
    "cg11436362",
    "cg13924996",
    "cg07420137",
    "cg24301930",
    "cg13395086",
    "cg20136100",
    "cg09153080",
    "cg09902130",
    "cg07380416",
    "cg27284288",
    "cg139321307",
    "cg10511890",
    "cg0024235",
    "cg04314978",
    "cg25225070",
    "cg20411756",
    "cg24247537",
    "cg04330884",
    "cg23130731",
    "cg04888360",
    "cg00907272",
    "cg05979232",
    "cg00025044",
    "cg04441857",
    "cg09684112",
    "cg27388962",
    "cg05931497",
    "cg13408086",
    "cg13555415",
    "cg22552736",
    "cg16191087",
    "cg13925432",
    "cg13464240",
    "cg14633252",
    "cg19252956",
    "cg00015530",
    "cg08632810",
    "cg1273392",
    "cg26769700",
    "cg03218479",
    "cg02609337",
    "cg10351284",
    "cg23555416",
    "cg19021985",
    "cg21031128",
    "cg19421584",
    "cg17984956",
    "cg05177060",
    "cg24107852",
    "cg25652701",
    "cg00282244",
    "cg18887230",
    "cg08486903",
    "cg09335715",
    "cg12629796",
    "cg16454130",
    "cg26433975",
    "cg10673833",
    "cg06787669",
    "cg12192582",
    "cg05098343",
    "cg07573366",
    "cg11105292",
    "cg05287480",
    "cg16748008",
    "cg16644023",
    "cg06488150",
    "cg09450197",
    "cg20336172",
    "cg08858130",
    "cg12098228",
    "cg26811313",
    "cg25435218",
    "cg1622899",
    "cg12359001",
    "cg01209642",
    "cg14564351",
    "cg23429794",
    "cg26401541",
    "cg20046343",
    "cg20847580",
    "cg03431741",
    "cg07417146",
    "cg09001226",
    "cg06482498",
    "cg03891050",
    "cg00899907",
    "cg13597051",
    "cg18113826",
    "cg04859102",
    "cg01620360",
    "cg14083015",
    "cg15046123",
    "cg03190513",
    "cg01456691",
    "cg17207512",
    "cg20510285",
    "cg01149192",
    "cg05614346",
    "cg064396655",
    "cg11334870",
    "cg0891922",
    "cg23021796",
    "cg2483 948",
    "cg10393744",
    "cg07428959",
    "cg17694130",
    "cg03956042",
    "cg19266387",
    "cg13512830",
    "cg19982684",
    "cg22513455",
    "cg07186032",
    "cg08052292",
    "cg27366280",
    "cg06825448",
    "cg25451702",
    "cg08098128",
    "cg13821008",
    "cg27405400",
    "cg09366118",
    "cg15341833",
    "cg02233149",
    "cg14247287",
    "cg23824762",
    "cg01604601",
    "cg05656900",
    "cg08132573",
    "cg24668618",
    "cg05352688",
    "cg18384097",
    "cg16266227",
    "cg19675731",
    "cg21461981",
    "Cg2577654",
    "cg26394055",
    "cg20685713",
    "cg23589035",
    "cg01903374",
    "cg23612220",
    "cg26315985",
    "cg188856478",
    "cg23229016",
    "cg21004490",
    "cg24742520",
    "cg23013029",
    "cg197053055",
    "cg0757590055",
    "cg0757590055",
}

Top_CpGs_patent_JP2018508228A_2 = {
    "cg25927251",
    "cg25443218",
    "cg23612220",
    "cg23113731",
    "cg139113392",
    "cg113334870",
    "cg11252953",
    "cg10542975",
    "cg08098128",
    "cg02874908",
    "cg267669927",
    "cg26769700",
    "cg25557465",
    "cg25490145",
    "cg18384097",
    "cg171256555",
    "cg142472873",
    "cg0274733",
    "cg0509873433",
    "cg0509873433",
    "cg66457",
    "cg19300307",
    "cg17122157",
    "cg13555415",
    "cg11436362",
    "cg10673833",
    "cg09866569",
    "cg08075204",
    "cg05614346",
    "cg02053964",
    "cg27377213",
    "cg24480810",
    "cg24301930",
    "cg22513455",
    "cg19693177",
    "cg19675731",
    "cg19252956",
    "cg18856478",
    "cg16509569",
    "cg15797834",
    "cg15698795",
    "cg15341833",
    "cg14556909",
    "cg14083015",
    "cg14058476",
    "cg12192582",
    "cg10590292",
    "cg0678769",
    "cg06439655",
    "cg0522196",
    "cg0223349",
    "cg00558804",
    "cg26680502",
    "cg23013029",
    "cg22552736",
    "cg21376733",
    "cg20847580",
    "cg19704755",
    "cg18842353",
    "cg16622899",
    "cg14999168",
    "cg13925432",
    "cg12967050",
    "cg11105292",
    "cg09419005",
    "cg09153080",
    "cg07380416",
    "cg06825448",
    "cg05596756",
    "cg03891050",
    "cg01681367",
    "cg01456691",
    "cg00015530",
    "cg27410601",
    "cg27366280",
    "cg26683005",
    "cg25666403",
    "cg24706505",
    "cg24107852",
    "cg23824762",
    "cg23021796",
    "cg21122474",
    "cg20336172",
    "cg18610205",
    "cg18456621",
    "cg17518965",
    "cg16748008",
    "cg16191087",
    "cg16061668",
    "cg14642045",
    "cg13924996",
    "cg12353452",
    "cg09335715",
    "cg08858130",
    "cg08480068",
    "cg08052292",
    "cg07428959",
    "cg06153925",
    "cg04147906",
    "cg03431741",
    "cg00282244",
    "cg00025044",
}

Top_CpGs_patent_US20200299776A1 = {"cg10673833"}


class RegressOut:
    @staticmethod
    def fit_transform(x, y):
        if y.ndim == 1:
            y = y.to_frame()
        confound_model = LinearRegression().fit(y, x)
        return x - confound_model.predict(y)


class MLiquidBiopsy:
    MANIFEST_COLS = [
        "CHR_hg38",
        "Start_hg38",
        "End_hg38",
        "Strand_hg38",
        "SourceSeq",
        "UCSC_CpG_Islands_Name",
        "Relation_to_UCSC_CpG_Island",
        "UCSC_RefGene_Name",
        "UCSC_RefGene_Group",
        "DNase_Hypersensitivity_NAME",
        "DNase_Hypersensitivity_Evidence_Count",
        "HMM_Island",
        "X450k_Enhancer",
    ]

    TOP_CPGS = {
        "Hlady et al.": Top_CpGs_Hlady,
        "Xu et al. Diagnosis": Top_CpGs_Xu_diagnosis,
        "Xu et al. prognosis": Top_CpGs_Xu_prognosis,
        "Villanueva et al.": Top_CpGs_villanueva,
        "Cheng et al.": Top_CpGs_Cheng_et_al,
        "KR102103885B1": Top_CpGs_patent_KR102103885B1,
        "US-2019300965-A1": Top_CpGs_patent_US2019300965A1,
        "US-2020263256-A1": Top_CpGs_patent_US2020263256A1,
        # "CN110603329A": Top_CpGs_patent_CN110603329A,
        "US20180216195A1": Top_CpGs_patent_US20180216195A1,
        "JP2018508228A_1": Top_CpGs_patent_JP2018508228A_1,
        "JP2018508228A_2": Top_CpGs_patent_JP2018508228A_2,
        "US20200299776A1": Top_CpGs_patent_US20200299776A1,
    }

    @staticmethod
    def make_it_float(v):
        try:
            v = float(v)
        except ValueError:
            return np.nan
        return v

    @classmethod
    def read_test_dataset_GSE89852(cls):
        df = pd.read_csv(
            f"~/datasets/Methylation_450k_GSE89852/GSE89852_Matrix_Processed.csv.gz",
            index_col=0,
        )
        df = df.loc[:, [not c.startswith("Detection Pval") for c in df]]
        df = df.applymap(cls.make_it_float).dropna().round(5)

        mnf = pd.read_csv(
            "~/datasets/Methylation_450k_GSE89852/GSE89852_series_matrix.txt",
            sep="\t",
            index_col="Sample_title",
        ).T.reset_index()
        mnf.index = mnf["Sample_supplementary_file_grn"].apply(
            lambda v: v.split("/")[-1].split("_")[1]
        )
        mnf["y_true"] = (
            mnf["Sample_characteristics_ch1_sample"]
            == "tissue: Hepatocellular carcinoma"
        ).astype(int)

        mnf["dataset_id"] = "GSE89852"
        mnf["dataset_color"] = "#DA5565"

        return mnf, df

    @classmethod
    def read_test_dataset_GSE77269(cls):
        df_files = os.listdir(
            f"/Users/emanuel/datasets/Methylation_450k_GSE77269/idats/"
        )

        df = pd.concat(
            [
                pd.read_csv(
                    f"~/datasets/Methylation_450k_GSE77269/idats/{f}",
                    index_col=0,
                    sep="\t",
                    header=None,
                )[1].rename(f.split("_")[0])
                for f in df_files
                if f.endswith(".txt.gz")
            ],
            axis=1,
        )
        df = df.astype(float).round(5).dropna()

        mnf = (
            pd.read_csv(
                "~/datasets/Methylation_450k_GSE77269/GSE77269_series_matrix.txt",
                sep="\t",
                index_col="Sample_title",
            )
            .T.reset_index()
            .set_index("Sample_geo_accession")
        )
        mnf["y_true"] = (
            mnf["Sample_characteristics_ch1_tissue"]
            .isin(
                ["tissue: Primary tumor", "tissue: Portal vein tumor thrombosis (PVTT)"]
            )
            .astype(int)
        )

        mnf["dataset_id"] = "GSE77269"
        mnf["dataset_color"] = "#4A89DC"

        return mnf, df

    @classmethod
    def read_test_dataset_GSE75041(cls):
        df = pd.read_csv(
            f"~/datasets/Methylation_450k_GSE75041/GSE75041_series_matrix_beta.txt",
            index_col=0,
            sep="\t",
        )
        df = df.astype(float).round(5).dropna()

        mnf = (
            pd.read_csv(
                "~/datasets/Methylation_450k_GSE75041/GSE75041_series_matrix.txt",
                sep="\t",
                index_col="Sample_title",
            )
            .T.reset_index()
            .set_index("Sample_geo_accession")
        )
        mnf["y_true"] = (
            mnf["Sample_characteristics_ch1_tissue"]
            .isin(["tissue: early-stage hepatocellular carcinoma"])
            .astype(int)
        )

        mnf["dataset_id"] = "GSE75041"
        mnf["dataset_color"] = "#F6BB42"

        return mnf, df

    @classmethod
    def read_test_dataset_GSE54503(cls):
        df = pd.read_csv(
            f"~/datasets/Methylation_450k_GSE54503/GSE54503_series_matrix_beta.txt",
            index_col=0,
            sep="\t",
        )
        df = df.astype(float).round(5).dropna()

        mnf = (
            pd.read_csv(
                "~/datasets/Methylation_450k_GSE54503/GSE54503_series_matrix.txt",
                sep="\t",
                index_col="Sample_title",
            )
            .T.reset_index()
            .set_index("Sample_geo_accession")
        )
        mnf["y_true"] = mnf["Sample_source_name_ch1"].isin(["HCC tumor"]).astype(int)

        mnf["dataset_id"] = "GSE54503"
        mnf["dataset_color"] = "#D770AD"

        return mnf, df

    @classmethod
    def read_test_dataset_GSE56588(cls):
        mnf = pd.read_csv(
            "~/datasets/Methylation_450k_GSE56588/GSE56588_series_matrix.txt",
            sep="\t",
            index_col="Sample_title",
        ).T
        mnf["y_true"] = mnf["Sample_source_name_ch1"].isin(["HCC_liver"]).astype(int)

        df = pd.read_csv(
            f"~/datasets/Methylation_450k_GSE56588/GSE56588_Heptromics_avrg_beta.txt.gz",
            index_col=0,
            sep="\t",
        )
        df = df.loc[:, [not c.endswith("_pval") for c in df]]
        df = df.loc[:, df.columns.isin(mnf.index)]

        mnf["dataset_id"] = "GSE56588"
        mnf["dataset_color"] = "#37BC9B"

        return mnf, df

    @classmethod
    def read_test_dataset_GSE136583(cls):
        mnf = (
            pd.read_csv(
                "~/datasets/Methylation_450k_GSE136583/GSE136583_series_matrix.txt",
                sep="\t",
                index_col="Sample_title",
            )
            .T.reset_index()
            .set_index("Sample_geo_accession")
        )
        mnf["y_true"] = (
            mnf["Sample_characteristics_ch1_tissue"]
            .isin(["tissue: hepatocellular carcinoma"])
            .astype(int)
        )

        df = pd.read_csv(
            f"~/datasets/Methylation_450k_GSE136583/GSE136583_series_matrix_beta.txt",
            index_col=0,
            sep="\t",
        )
        df = df.astype(float).round(5).dropna()

        mnf["dataset_id"] = "GSE136583"
        mnf["dataset_color"] = "#967ADC"

        return mnf, df

    @classmethod
    def read_test_dataset_GSE113019(cls):
        mnf = (
            pd.read_csv(
                "~/datasets/Methylation_450k_GSE113019/GSE113019_series_matrix.txt",
                sep="\t",
                index_col="Sample_title",
            )
            .T.reset_index()
            .set_index("Sample_geo_accession")
        )
        mnf["y_true"] = (
            mnf["Sample_characteristics_ch1"]
            .isin(
                [
                    "tissue: Liver cancer, primary tumorous tissue",
                    "tissue: Liver cancer, recurrent tumorous tissue",
                    "tissue: Liver cancer, re-recurrent tumorous tissue",
                ]
            )
            .astype(int)
        )

        df = pd.read_csv(
            f"~/datasets/Methylation_450k_GSE113019/GSE113019_series_matrix_beta.txt",
            index_col=0,
            sep="\t",
        )
        df = df.astype(float).round(5).dropna()

        mnf["dataset_id"] = "GSE113019"
        mnf["dataset_color"] = "#8CC152"

        return mnf, df

    @classmethod
    def read_test_datasets(cls, dataset_id):
        if dataset_id == "GSE89852":
            return cls.read_test_dataset_GSE89852()

        elif dataset_id == "GSE77269":
            return cls.read_test_dataset_GSE77269()

        elif dataset_id == "GSE75041":
            return cls.read_test_dataset_GSE75041()

        elif dataset_id == "GSE54503":
            return cls.read_test_dataset_GSE54503()

        elif dataset_id == "GSE56588":
            return cls.read_test_dataset_GSE56588()

        elif dataset_id == "GSE136583":
            return cls.read_test_dataset_GSE136583()

        elif dataset_id == "GSE113019":
            return cls.read_test_dataset_GSE113019()

        else:
            return cls.read_all_test_datasets()

    @classmethod
    def read_all_test_datasets(cls, dataset_ids=None):
        if dataset_ids is None:
            dataset_ids = [
                "GSE89852",
                "GSE77269",
                "GSE75041",
                "GSE54503",
                "GSE56588",
                "GSE136583",
                "GSE113019",
            ]

        dfs, mnfs = [], []

        for dataset_id in dataset_ids:
            mnf, df = cls.read_test_datasets(dataset_id)
            mnfs.append(mnf)
            dfs.append(df)

        df = pd.concat(dfs, axis=1)
        mnfs = pd.concat(mnfs)

        return mnfs, df

    @staticmethod
    def sample_class(stype, hcc_labels=None):
        if hcc_labels is None:
            hcc_labels = [
                "Primary Tumor - Liver",
                "cfDNA HCC and cirrhosis",
                "HCC + EtOH",
                "HCC + HCV",
            ]

        if stype in hcc_labels:
            return 1

        else:
            return 0

    @classmethod
    def import_samplesheet(cls, subset_labels=None, subset_samples=None):
        ss_all = pd.read_csv(
            f"~/datasets/DNAMethylationPipeline_samplesheet.csv.gz", index_col=0
        )
        ss_all["Type"] = (
            ss_all["label"]
            .apply(cls.sample_class)
            .replace({0: "Other", 1: "HCC"})
            .rename("Type")
            .values
        )

        if subset_labels is not None:
            ss_all = ss_all[ss_all["label"].isin(subset_labels)]

        if subset_samples is not None:
            ss_all = ss_all[ss_all.index.isin(subset_samples)]

        pal_label = ss_all.groupby("label")["palette"].first()
        pal_type = dict(Other=CrispyPlot.PAL_DTRACE[0], HCC=CrispyPlot.PAL_DTRACE[1])

        # Add predicted gender
        pred_gender = pd.read_csv(f"~/datasets/DNAMethylationPipelineQCReport_MergedSubset_GenderPredicted.csv", index_col=0)
        ss_all["gender_predicted"] = pred_gender.loc[ss_all.index, "predictedSex"].values

        # Add predicted age
        pred_age = pd.read_csv(f"~/datasets/DNAMethylationPipelineQCReport_MergedSubset_AgePredicted.csv", index_col=0)

        ss_all["age_predicted_horvath"] = pred_age.loc[ss_all.index, "horvath"].values
        ss_all["age_predicted_hannum"] = pred_age.loc[ss_all.index, "hannum"].values

        return ss_all, pal_label, pal_type

    @classmethod
    def import_dataset(cls, dpath, subset_labels=None):
        # Import dataset
        methy_b = pd.read_csv(dpath)

        # Exclude problematic samples
        methy_b = methy_b.drop(columns=["8784233035_R03C02"])

        # Import samplesheet
        ss_all, pal_label, pal_type = cls.import_samplesheet(
            subset_labels=subset_labels,
            subset_samples=set(methy_b),
        )

        # Overlap samples
        methy_b = methy_b[[c for c in methy_b if c in ss_all.index]]

        return methy_b, ss_all, pal_label, pal_type

    @staticmethod
    def independent_features(df=None):
        cpg_dbs = MLiquidBiopsy.TOP_CPGS.copy()
        cpg_dbs_df = pd.DataFrame(
            [dict(cpg=cpg, db=db) for db in cpg_dbs for cpg in cpg_dbs[db]]
        )

        if df is not None:
            cpg_dbs_df = cpg_dbs_df[cpg_dbs_df["cpg"].isin(df.index)]

        return cpg_dbs_df.reset_index(drop=True)

    @staticmethod
    def contrasts(x, y):
        contrasts = {1: 1 / sum(y == 1), 0: -1 / sum(y == 0)}
        contrasts = y.replace(contrasts)
        contrasts = x.T.dot(contrasts)
        return contrasts

    @classmethod
    def anova_cov(cls, x, y, ss, rank_by="pval", dataset_as_cov=False, extra_covs=None, add_contrasts=True):
        m = pd.concat([
            ss["gender_predicted"].str.get_dummies()["F"].rename("Female"),
            LMModels.transform_matrix(ss[[
                "age_predicted_horvath", "age_predicted_hannum"
            ]], t_type="scale"),
        ], axis=1).loc[x.index]

        if dataset_as_cov:
            m = pd.concat([
                m,
                ss.loc[x.index, "dataset_id"].str.get_dummies(),
            ], axis=1)

        if extra_covs is not None:
            m = pd.concat([
                m,
                extra_covs,
            ], axis=1)

        LOG.info(f"Covariates: {list(m)}")

        lm_model = LModel(Y=x, X=y[x.index].rename("HCC").to_frame(), M=m.loc[x.index]).fit_matrix()
        lm_model["fdr"] = multipletests(lm_model["pval"], method="fdr_bh")[1]
        lm_model = lm_model.set_index("y_id").sort_values(rank_by)

        # F-scores
        anova_vals = pd.DataFrame(
            f_classif(x.loc[x.index], y[x.index]),
            index=["ANOVA_fval", "ANOVA_pval"],
            columns=x.columns,
        ).T.dropna()
        anova_vals["ANOVA_fdr"] = multipletests(anova_vals["ANOVA_pval"], method="fdr_bh")[1]
        lm_model = pd.concat([lm_model, anova_vals], axis=1)

        # Contrasts
        if add_contrasts:
            lm_model["diff"] = cls.contrasts(x, y[x.index])[lm_model.index].values

        return lm_model

    @classmethod
    def anova(
        cls,
        x,
        y,
        add_contrasts=True,
        rank_by="pval",
        probe_varaince=None,
        force_multiclass=False,
    ):
        samples = set(x.dropna().index).intersection(set(y.dropna().index))

        # F-scores
        anova_vals = pd.DataFrame(
            f_classif(x.loc[samples], y[samples]),
            index=["fval", "pval"],
            columns=x.columns,
        ).T.dropna()
        anova_vals["fdr"] = multipletests(anova_vals["pval"], method="fdr_bh")[1]

        # Chi2
        chi2_vals = pd.DataFrame(
            chi2(x.loc[samples], y[samples]),
            index=["chi2", "chi2_pval"],
            columns=x.columns,
        ).T.dropna()
        chi2_vals["chi2_fdr"] = multipletests(chi2_vals["chi2_pval"], method="fdr_bh")[
            1
        ]

        vals = pd.concat([anova_vals, chi2_vals], axis=1)

        # Contrasts
        if add_contrasts:
            if type_of_target(y) == "multiclass" or force_multiclass:
                for c in y.unique():
                    vals[f"diff_{c}"] = cls.contrasts(x.loc[samples], y[samples] == c)[vals.index].values
            else:
                vals["diff"] = cls.contrasts(x.loc[samples], y[samples])[vals.index].values

        # Rank
        if (rank_by == "abs_diff") and add_contrasts:
            vals = vals[vals["diff"].abs().sort_values(ascending=False).index]
        else:
            vals = vals.sort_values("pval")

        # Probe variance
        if probe_varaince is not None:
            vals["variance"] = probe_varaince[vals.index].values

        vals["n_samples"] = len(samples)

        return vals

    @staticmethod
    def scale_dataframe(df):
        return pd.DataFrame(
            StandardScaler().fit_transform(df),
            index=df.index,
            columns=df.columns,
        ).dropna()

    @classmethod
    def dimension_reduction(cls, df, samplesheet=None, n_components=2):
        df_scaled = cls.scale_dataframe(df)

        # PCA
        pca = PCA(n_components=n_components).fit(df_scaled)
        pca_dim = pd.DataFrame(
            pca.transform(df_scaled),
            index=df_scaled.index,
            columns=[f"PC{d}" for d in range(1, n_components + 1)],
        )
        pca_vexp = pd.Series(
            pca.explained_variance_ratio_,
            index=[f"PC{d}" for d in range(1, n_components + 1)],
        )

        # UMAP
        umap = UMAP()
        umap_dim = pd.DataFrame(
            umap.fit_transform(df_scaled),
            index=df_scaled.index,
            columns=["Dim1", "Dim2"],
        )

        # Append samplesheet information
        if samplesheet is not None:
            pca_dim = pd.concat([pca_dim, samplesheet.loc[pca_dim.index]], axis=1)
            umap_dim = pd.concat([umap_dim, samplesheet.loc[umap_dim.index]], axis=1)

        return pca_dim, umap_dim, pca_vexp

    @staticmethod
    def read_probe_annot_450k():
        return pd.read_csv(
            f"~/datasets/DNAMethylationMergePreprocessed_methy_annotation.csv"
        )

    @staticmethod
    def read_probe_annot_epic():
        return pd.read_csv(
            f"~/datasets/DNAMethylationPipeline_methy_annotation_EPIC.csv"
        )

    @classmethod
    def read_probe_annot(cls):
        pannot = pd.read_csv(
            f"~/datasets/infinium-methylationepic-v-1-0-b5-manifest-file.csv",
            index_col=0,
        )
        return pannot

    @classmethod
    def probes_sex_chr(cls):
        sex_chr = ["chrX", "chrY"]
        return set(cls.read_probe_annot().query("CHR_hg38 in @sex_chr").index)

    @classmethod
    def probes_protein_coding(cls):
        pannot = cls.read_probe_annot()
        pannot = pannot.dropna(subset=["UCSC_RefGene_Name", "UCSC_RefGene_Group"])
        return set(pannot.index)

    @classmethod
    def calculate_islands(cls, df, nmin=5):
        islands = cls.read_probe_annot().dropna(subset=["Islands_Name"])

        p_sex = cls.probes_sex_chr()
        islands = islands[~islands.index.isin(p_sex)]

        islands = islands[islands.index.isin(df.index)]

        islands_counts = islands["Islands_Name"].value_counts()
        islands_counts = islands_counts[islands_counts >= nmin]
        islands = islands[islands["Islands_Name"].isin(islands_counts.index)]

        islands = df.groupby(islands["Islands_Name"]).mean()

        return islands

    @classmethod
    def calculate_promoter(cls, df, nmin=3, promoter_regions=None):
        if promoter_regions is None:
            promoter_regions = ["TSS1500", "TSS200", "5'UTR", "1stExon"]

        promoter = cls.read_probe_annot_epic().dropna(
            subset=["UCSC_RefGene_Name", "UCSC_RefGene_Group"]
        )

        p_sex = cls.probes_sex_chr()
        promoter = promoter[~promoter.index.isin(p_sex)]
        promoter = promoter[promoter.index.isin(df.index)]

        promoter = pd.Series(
            {
                i: g
                for i, r in promoter.iterrows()
                for g, r in zip(
                    r["UCSC_RefGene_Name"].split(";"),
                    r["UCSC_RefGene_Group"].split(";"),
                )
                if r in promoter_regions
            }
        )

        promoter_counts = promoter.value_counts()
        promoter_counts = promoter_counts[promoter_counts >= nmin]
        promoter = promoter[promoter.isin(promoter_counts.index)]
        promoter = promoter[promoter.index.isin(df.index)]

        df_promoter = df.groupby(promoter).mean()

        return df_promoter, promoter

    @classmethod
    def probes_cluster(cls, df, nmin=3):
        pannot = cls.read_probe_annot().dropna(subset=["cluster"])
        pannot = pannot[pannot.index.isin(df.index)]

        cannot = pannot["cluster"].copy()

        cluster_counts = cannot.value_counts()
        cluster_counts = cluster_counts[cluster_counts >= nmin]
        cannot = cannot[cannot.isin(cluster_counts.index)]

        df_cluster = df.groupby(cannot).mean()

        return df_cluster, cannot

    @staticmethod
    def eval_pred(y_true, y_pred, as_str=True):
        metrics = [
            ("recall", recall_score),
            ("precision", precision_score),
            ("mcc", matthews_corrcoef),
            ("accuracy", accuracy_score),
            ("balanced accuracy", balanced_accuracy_score),
        ]

        m_scores = dict()
        for m, mfunc in metrics:
            if m in ["recall", "precision"]:
                m_scores[m] = mfunc(
                    y_true,
                    y_pred,
                    average="macro"
                    if type_of_target(y_true) == "multiclass"
                    else "binary",
                )

            else:
                m_scores[m] = mfunc(y_true, y_pred)

        if as_str:
            m_scores = "; ".join([f"{m}={s:.2f}" for m, s in m_scores.items()])

        return m_scores

    @staticmethod
    def merge_corr_features(df, corr_linkage_thres=1, debug=False):
        corr = spearmanr(df).correlation
        corr_linkage = hierarchy.ward(corr)
        corr_cluster_ids = hierarchy.fcluster(
            corr_linkage, corr_linkage_thres, criterion="distance"
        )
        corr_cluster_ids = pd.Series(corr_cluster_ids, index=df.columns)

        df_reduced = df.groupby(corr_cluster_ids, axis=1).mean()
        df_reduced.columns = [f"Cluster{c}" for c in df_reduced]

        df_cluster_id = (
            corr_cluster_ids.reset_index()
            .groupby(0)["index"]
            .agg(lambda v: "; ".join(set(v)))
        )
        df_cluster_id.index = [f"Cluster{c}" for c in df_cluster_id.index]

        if debug:
            fig, (ax1, ax2) = plt.subplots(1, 2, figsize=(8, 6))

            dendro = hierarchy.dendrogram(corr_linkage, no_labels=True, ax=ax1)
            ax1.axhline(corr_linkage_thres, ls="--", lw=0.3)

            ax2.imshow(corr[dendro["leaves"], :][:, dendro["leaves"]])

            fig.tight_layout()
            plt.show()

        return df_reduced, df_cluster_id


class MLiquidBiopsyPlot:
    @staticmethod
    def anova_top_features_plot(x, y, anova_top_res):
        if type(anova_top_res) == set:
            plot_df = x.reindex(columns=anova_top_res).unstack().reset_index()
            n_feat = len(anova_top_res)

        else:
            plot_df = x[anova_top_res.index].unstack().reset_index()
            n_feat = anova_top_res.shape[0]

        plot_df.columns = ["feature", "sample", "beta"]
        plot_df["y"] = y.loc[plot_df["sample"]].values

        fig, ax = plt.subplots(1, 1, figsize=(2, 0.25 * n_feat))

        sns.boxenplot(
            x="beta",
            y="feature",
            hue="y",
            data=plot_df,
            linewidth=0.3,
            palette=CrispyPlot.PAL_DTRACE,
            saturation=1,
            orient="h",
            ax=ax,
        )

        ax.legend(
            [Line2D([0], [0], color=CrispyPlot.PAL_DTRACE[i], lw=4) for i in [0, 1]],
            ["No", "Yes"],
            frameon=False,
            loc="center left",
            bbox_to_anchor=(1, 0.5),
            title=y.name,
        )

        ax.set_xlabel("Methylation (beta)")
        ax.set_ylabel("")
        ax.grid(axis="x", lw=0.1, color="#e1e1e1", zorder=0)

        return ax

    @staticmethod
    def dimension_reduction_plot(
        x_var,
        y_var,
        hue,
        plot_df,
        method="PCA",
        vexp=None,
        alpha=1,
        palette=None,
        hue_order=None,
    ):
        ax = GIPlot.gi_tissue_plot(
            x_var,
            y_var,
            plot_df=plot_df,
            hue=hue,
            hue_order=hue_order,
            plot_reg=False,
            annot=False,
            pal=palette,
            alpha=alpha,
        )

        ax.set_xlabel(
            x_var if method == "UMAP" else f"{x_var} ({vexp[x_var]*100:.1f}%)"
        )
        ax.set_ylabel(
            y_var if method == "UMAP" else f"{y_var} ({vexp[y_var]*100:.1f}%)"
        )

        ax.set_title(method)

        return ax

    @staticmethod
    def eval_barplot(cls_results, palette=None, plot_it=True):
        if palette is None:
            n_colors = list(cls_results[list(cls_results)[0]]["features"])
            palette = pd.Series(
                sns.color_palette("tab20c", n_colors=len(n_colors)).as_hex(),
                index=n_colors,
            )

        # Barplot
        def specificity_score_(t, p, average=None):
            return specificity_score(t, p)

        def matthews_corrcoef_(t, p, average=None):
            return matthews_corrcoef(t, p)

        def balanced_accuracy_score_(t, p, average=None):
            return balanced_accuracy_score(t, p)

        metrics = [
            ("recall", recall_score),
            ("precision", precision_score),
            ("specificity", specificity_score_),
            ("mcc", matthews_corrcoef_),
            ("balanced accuracy", balanced_accuracy_score_),
        ]

        plot_df = pd.DataFrame(
            [
                dict(
                    ftype=ftype,
                    ptype=ptype,
                    metric=mtype,
                    value=mfunc(
                        cls_results[ptype]["y_true"],
                        cls_results[ptype]["y_pred"][ftype],
                        average="macro"
                        if type_of_target(cls_results[ptype]["y_true"]) == "multiclass"
                        else "binary",
                    ),
                )
                for ptype in cls_results
                for ftype in cls_results[ptype]["y_pred"]
                for mtype, mfunc in metrics
                if (type_of_target(cls_results[ptype]["y_true"]) != "multiclass")
                or (
                    type_of_target(cls_results[ptype]["y_true"]) == "multiclass"
                    and mtype not in ["specificity"]
                )
            ]
        )

        if plot_it:
            fig, axs = plt.subplots(
                len(cls_results),
                1,
                figsize=(2, 1.5 * len(cls_results)),
                sharey="all",
                sharex="all",
                squeeze=False,
            )
            axs = axs[:, 0]

            for i, y_var in enumerate(cls_results):
                df = plot_df.query(f"ptype == '{y_var}'")

                sns.barplot(
                    x="value",
                    y="metric",
                    hue="ftype",
                    data=df,
                    lw=0,
                    palette=palette,
                    saturation=1,
                    ax=axs[i],
                )

                axs[i].set_xlabel("")
                axs[i].set_ylabel(y_var)
                axs[i].grid(axis="x", lw=0.1, color="#e1e1e1", zorder=0)
                axs[i].legend(
                    frameon=False,
                    prop=dict(size=4),
                    loc="center left",
                    bbox_to_anchor=(1, 0.5),
                    title=f"Predicting {y_var}",
                ).get_title().set_fontsize("5")

            plt.subplots_adjust(hspace=0.05)

        return plot_df

    @staticmethod
    def eval_confusion_matrices(cls_results):
        nrows = len(cls_results)
        ncols = cls_results[list(cls_results)[0]]["y_pred"].shape[1]

        fig, axs = plt.subplots(
            nrows,
            ncols,
            figsize=(1 * ncols, 1 * nrows),
            sharey="all",
            sharex="all",
            squeeze=False,
        )

        for i, y_var in enumerate(cls_results):
            for j, ftype in enumerate(cls_results[y_var]["y_pred"]):
                labels = cls_results[y_var]["y_true"].unique()

                cf_matrix = confusion_matrix(
                    cls_results[y_var]["y_true"],
                    cls_results[y_var]["y_pred"][ftype],
                    labels=labels,
                )
                cf_matrix = pd.DataFrame(cf_matrix, index=labels, columns=labels)

                if len(labels) == 2:
                    cf_matrix = cf_matrix.reindex(index=[0, 1], columns=[0, 1])

                sns.heatmap(
                    cf_matrix,
                    annot=True,
                    cbar=False,
                    fmt=".0f",
                    annot_kws=dict(size=7),
                    cmap="Blues",
                    ax=axs[i][j],
                )

                axs[i][j].set_ylabel(
                    f"{y_var}\nTrue label" if j == 0 else "", fontsize=6
                )
                axs[i][j].set_xlabel(
                    f"Pred label" if i == nrows - 1 else "", fontsize=6
                )
                axs[i][j].set_title(f"{ftype}" if i == 0 else "", fontsize=6)

        plt.gca().invert_yaxis()

        plt.subplots_adjust(hspace=0.1, wspace=0.1)

    @staticmethod
    def recall_precision_curve(y_true, y_score, color=None, plot_f1curves=True, plot_random=True, plot_legend=True, ax=None, custom_marker=None):
        if ax is None:
            _, ax = plt.subplots(
                1,
                1,
                figsize=(2, 2),
            )

        if plot_f1curves:
            for f_score in np.linspace(0.2, 0.8, num=4):
                f_x = np.linspace(0.01, 1., 1000)
                f_y = f_score * f_x / (2 * f_x - f_score)

                ax.plot(
                    f_x[(f_y >= 0) & (f_y <= 1.5)],
                    f_y[(f_y >= 0) & (f_y <= 1.5)],
                    color="#CCCCCC",
                    alpha=0.2,
                    lw=0.5,
                )

                ax.annotate(
                    f"f1={f_score:0.1f}", xy=(1, f_score / (2 - f_score)),
                    fontsize=5,
                    ha="right",
                )

        precision, recall, thresholds = precision_recall_curve(y_true, y_score)
        f1_score_func = lambda p, r: 2 * (p * r) / (p + r)
        f1_scores = f1_score_func(precision, recall)

        f1_max = max(f1_scores)
        f1_max_idx = np.argmax(f1_scores)

        auc = average_precision_score(y_true, y_score)

        ax.plot(
            recall,
            precision,
            label=f"{'' if custom_marker is None else f'{custom_marker}. '}{y_score.name} (AUC={auc:.2f}; F1={f1_max:.2f})",
            c=CrispyPlot.PAL_DTRACE[2] if color is None else color,
        )

        ax.scatter(
            recall[f1_max_idx],
            precision[f1_max_idx],
            c=CrispyPlot.PAL_DTRACE[2] if color is None else color,
            marker="X",
            edgecolor="white",
            lw=0.3,
            s=20,
            alpha=1,
            zorder=3,
        )

        if custom_marker is not None:
            ax.text(
                recall[f1_max_idx],
                precision[f1_max_idx],
                custom_marker,
                fontsize=5,
            )

        if plot_random:
            ax.axhline(
                y_true.sum() / y_true.shape[0],
                c="#CCCCCC",
                lw=1.0,
                label="random",
                zorder=0,
                ls="--",
            )

        if plot_legend:
            ax.legend(
                frameon=False,
                loc=3,
                prop=dict(size=5),
            )

        ax.grid(axis="both", lw=0.1, color="#e1e1e1", zorder=0)

        ax.set_xlabel("Recall")
        ax.set_ylabel(f"Precision")

        ax.set_xlim([0.0, 1.0])
        ax.set_ylim([0.0, 1.0])

        return ax, thresholds[f1_max_idx], recall[f1_max_idx], precision[f1_max_idx]

    @staticmethod
    def eval_recall_precision_curve(cls_results, palette=None, feature_set=None):
        if palette is None:
            n_colors = list(cls_results[list(cls_results)[0]]["features"])
            palette = pd.Series(
                sns.color_palette("tab20c", n_colors=len(n_colors)).as_hex(),
                index=n_colors,
            )

        fig, axs = plt.subplots(
            len(cls_results),
            1,
            figsize=(3, 3 * len(cls_results)),
            sharey="all",
            sharex="all",
            squeeze=False,
        )
        axs = axs[:, 0]

        for i, y_var in enumerate(cls_results):
            for f_score in np.linspace(0.2, 0.8, num=4):
                f_x = np.linspace(0.01, 1)
                f_y = f_score * f_x / (2 * f_x - f_score)
                (f_l,) = axs[i].plot(
                    f_x[f_y >= 0], f_y[f_y >= 0], color="gray", alpha=0.2
                )
                axs[i].annotate(
                    f"f1={f_score:0.1f}", xy=(0.9, f_y[45] + 0.02), fontsize=4
                )

            for ftype in cls_results[y_var]["y_pred"]:
                if ftype == "random":
                    continue

                precision, recall, _ = precision_recall_curve(
                    cls_results[y_var]["y_true"], cls_results[y_var]["y_proba"][ftype]
                )
                auc = average_precision_score(
                    cls_results[y_var]["y_true"], cls_results[y_var]["y_proba"][ftype]
                )
                mcc = matthews_corrcoef(
                    cls_results[y_var]["y_true"], cls_results[y_var]["y_pred"][ftype]
                )

                if feature_set is None:
                    n_features = cls_results[y_var]["y_features"][ftype].dropna()
                    n_features = n_features[n_features != 0]
                else:
                    n_features = feature_set[ftype]

                axs[i].plot(
                    recall,
                    precision,
                    label=f"{ftype} (N={len(n_features)}, AUC={auc:.2f};MCC={mcc:.2f})",
                    c=palette[ftype],
                )

                p = precision_score(
                    cls_results[y_var]["y_true"], cls_results[y_var]["y_pred"][ftype]
                )
                r = recall_score(
                    cls_results[y_var]["y_true"], cls_results[y_var]["y_pred"][ftype]
                )

                axs[i].scatter(
                    r,
                    p,
                    c=palette[ftype],
                    marker="X",
                    edgecolor="white",
                    lw=0.3,
                    s=20,
                    alpha=1,
                    zorder=3,
                )

            axs[i].axhline(
                cls_results[y_var]["y_true"].sum()
                / cls_results[y_var]["y_true"].shape[0],
                c="#d9d9d9",
                lw=1.0,
                label="random",
                zorder=0,
                ls="--",
            )

            axs[i].grid(axis="both", lw=0.1, color="#e1e1e1", zorder=0)

            axs[i].set_xlabel("Recall" if i == len(cls_results) - 1 else "")
            axs[i].set_ylabel(f"{y_var}\nPrecision")

            axs[i].set_xlim([0.0, 1.0])
            axs[i].set_ylim([0.0, 1.0])

            axs[i].legend(
                frameon=False,
                prop=dict(size=6),
                loc="center left",
                bbox_to_anchor=(1, 0.5),
                title=f"Predicting {y_var}",
            ).get_title().set_fontsize("6")

        plt.subplots_adjust(hspace=0.05, wspace=0.05)

    @staticmethod
    def eval_features_venn(cls_results):
        features_dict = dict()

        for y_var in cls_results:
            for ftype in cls_results[y_var]["y_features"]:
                ftype_features = cls_results[y_var]["y_features"][ftype].dropna()
                ftype_features = ftype_features[ftype_features != 0]
                features_dict[f"{y_var} {ftype}"] = set(ftype_features.index)

        features_dict = from_contents(features_dict)

        ax = upsetplot(features_dict)

        return ax

    @staticmethod
    def methylation_region(
        feature,
        methy,
        probes_annotation,
        samplesheet,
        hue="label",
        palette=None,
        genomic_range=1e4,
        hue_order=None,
        feature_highlights=None,
        patent_palette=False,
    ):
        if feature_highlights is None:
            feature_highlights = {}

        # CpG annotation
        fannot = probes_annotation.loc[feature]

        # Select neighbour CpGs
        fneigh = probes_annotation.query(f"CHR_hg38 == '{fannot.CHR_hg38}'")

        fneigh = fneigh.loc[
            (fneigh.pos > (fannot.pos - genomic_range))
            | fneigh.index.isin(feature_highlights)
        ]
        fneigh = fneigh.loc[
            (fneigh.pos < (fannot.pos + genomic_range))
            | fneigh.index.isin(feature_highlights)
        ]

        # Build annotation
        plot_df = pd.concat(
            [
                fneigh[["CHR_hg38", "pos"]],
                methy.reindex(index=fneigh.index, columns=samplesheet.index),
            ],
            axis=1,
        ).reset_index()

        plot_df = pd.melt(plot_df, id_vars=["IlmnID", "CHR_hg38", "pos"]).dropna()
        plot_df[hue] = pd.Categorical(
            samplesheet.reindex(plot_df["variable"])[hue].values,
            ordered=True,
            categories=hue_order,
        )

        plot_df = plot_df.sort_values(["pos", "Type"])

        # Misc
        order = list(unique_everseen(plot_df["IlmnID"]))
        hue_order = set(plot_df[hue]) if hue_order is None else hue_order
        palette = (
            samplesheet.groupby(hue)["palette"].first().to_dict()
            if palette is None
            else palette
        )

        # Plot
        fig, ax = plt.subplots(1, 1, figsize=(len(order) * 0.35, 2))

        sns.boxplot(
            x="IlmnID",
            y="value",
            hue=hue,
            data=plot_df,
            boxprops=dict(linewidth=0.3),
            whiskerprops=dict(linewidth=0.3),
            medianprops=CrispyPlot.MEDIANPROPS,
            flierprops=CrispyPlot.FLIERPROPS,
            showcaps=False,
            saturation=1,
            palette=palette,
            order=order,
            hue_order=hue_order,
            ax=ax,
        )

        if patent_palette:
            ax.set_xlabel("")

        else:
            ax.set_xlabel(f"{feature} {fannot.CHR_hg38} {fannot.UCSC_CpG_Islands_Name} {fannot.Relation_to_UCSC_CpG_Island} {fannot.UCSC_RefGene_Name} {fannot.UCSC_RefGene_Group}\nGenomic range +/-{genomic_range:.0f}bp")

        ax.set_ylabel("Methylation (beta)")
        ax.grid(axis="y", lw=0.1, color="#CCCCCC" if patent_palette else "#e1e1e1", zorder=0)

        loc, labels = plt.xticks()
        ax.set_xticklabels(labels, rotation=45, ha="right")

        for i, label in enumerate(ax.get_xticklabels()):
            if label.get_text() == feature or label.get_text() in feature_highlights:
                ax.get_xticklabels()[i].set_color("#000000" if patent_palette else "red")
                ax.get_xticklabels()[i].set_weight("bold")

        ax.legend(
            [Line2D([0], [0], color=palette[i], lw=2) for i in hue_order],
            hue_order,
            frameon=False,
            prop=dict(size=4),
            loc="center left",
            bbox_to_anchor=(1, 0.5),
        )

        return ax

    @staticmethod
    def probe_annotation_name(
        probe_annotation, add_island=True, add_gene=True, add_regulatory=True
    ):
        p_label = f"{probe_annotation.name} {probe_annotation.chr}"

        if add_island and str(probe_annotation.Relation_to_Island) != "nan":
            p_label += f"; Island={probe_annotation.Relation_to_Island}"

        if add_gene and str(probe_annotation.UCSC_RefGene_Name).lower() != "nan":
            p_label += f"; Gene={probe_annotation.UCSC_RefGene_Name} {probe_annotation.UCSC_RefGene_Group}"

        if add_regulatory and str(probe_annotation.DHS).lower() != "nan":
            p_label += f"; DHS={probe_annotation.DHS}"

        if add_regulatory and str(probe_annotation.Enhancer).lower() != "nan":
            p_label += f"; Enhancer={probe_annotation.Enhancer}"

        return p_label

    @staticmethod
    def chromosome_collections(df, y_positions, height, **kwargs):
        del_width = False

        if "width" not in df.columns:
            del_width = True
            df["width"] = df["end"] - df["start"]

        for chrom, group in df.groupby("chrom"):
            yrange = (y_positions[chrom], height)
            xranges = group[["start", "width"]].values
            yield BrokenBarHCollection(
                xranges, yrange, facecolors=group["colors"], **kwargs
            )

        if del_width:
            del df["width"]

    @classmethod
    def chromosome_ideogram(
        cls,
        plot_df,
        chrom_height=1,
        chrom_spacing=1,
        gene_height=0.4,
        gene_padding=0.1,
        chromosome_list=None,
        autosomal_only=True,
        figsize=(6, 8)
    ):
        if chromosome_list is None:
            chromosome_list = [f"chr{i}" for i in range(1, 23)]

            if not autosomal_only:
                chromosome_list += ["chrM", "chrX", "chrY"]

        ybase = 0
        chrom_ybase = {}
        gene_ybase = {}
        chrom_centers = {}

        for chrom in chromosome_list[::-1]:
            chrom_ybase[chrom] = ybase
            chrom_centers[chrom] = ybase + chrom_height / 2.0
            gene_ybase[chrom] = ybase - gene_height - gene_padding
            ybase += chrom_height + chrom_spacing

        ideo = pd.read_table(
            "~/hg38_cytoBandIdeo.txt",
            skiprows=1,
            names=["chrom", "start", "end", "name", "gieStain"],
        )

        ideo = ideo[ideo.chrom.apply(lambda x: x in chromosome_list)]
        ideo["width"] = ideo.end - ideo.start

        color_lookup = {
            "gneg": (1.0, 1.0, 1.0),
            "gpos25": (0.6, 0.6, 0.6),
            "gpos50": (0.4, 0.4, 0.4),
            "gpos75": (0.2, 0.2, 0.2),
            "gpos100": (0.0, 0.0, 0.0),
            "acen": (0.8, 0.4, 0.4),
            "gvar": (0.8, 0.8, 0.8),
            "stalk": (0.9, 0.9, 0.9),
        }

        # Add a new column for colors
        ideo["colors"] = ideo["gieStain"].apply(lambda x: color_lookup[x])

        # plot
        fig, ax = plt.subplots(1, 1, figsize=figsize)

        for collection in cls.chromosome_collections(ideo, chrom_ybase, chrom_height, linewidth=0.1, edgecolor="k"):
            ax.add_collection(collection)

        for (n, c), df in plot_df.query(f"chrom in @chromosome_list").groupby(["db", "colors"]):
            for collection in cls.chromosome_collections(
                df, gene_ybase, gene_height, alpha=0.5, linewidth=1, edgecolor=c, label=n
            ):
                ax.add_collection(collection)

        # Axes tweaking
        ax.set_yticks([chrom_centers[i] for i in chromosome_list])
        ax.set_yticklabels(chromosome_list)
        ax.axis("tight")

        return ax

    @staticmethod
    def sample_markers(s):
        if s == "DMR signature":
            return "o"
        # elif s == "Hlady et al.":
        #     return "s"
        else:
            return "X"

    @staticmethod
    def sample_color(s, patent_color=False):
        if s == "o" or s == "DMR signature":
            if patent_color:
                return "#000000"
            else:
                return CrispyPlot.PAL_DTRACE[3]
        else:
            if patent_color:
                return "#CCCCCC"
            else:
                return CrispyPlot.PAL_DTRACE[0]

    @staticmethod
    def sample_label(s):
        if s == "o":
            return "DMR signature"
        elif s == "s":
            return "Hlady et al."
        else:
            return "Other"

    @staticmethod
    def sample_size(s):
        if s == "o" or s == "DMR signature":
            return 20
        # elif s == "s":
        #     return 10
        else:
            return 10
```
