## Supplementary File 1 for "DNA methylation fingerprint for the diagnosis and monitoring of hepatocellular carcinoma from tissue and liquid biopsies": MulticlassMergedTraining.py

```
import numpy as np
import pandas as pd
import seaborn as sns
import matplotlib.pyplot as plt
from LiquidBiopsy import LOG
from crispy import CrispyPlot
from crispy.GIPlot import GIPlot
from sklearn.svm import LinearSVC
from matplotlib_venn import venn3, venn3_circles
from sklearn.model_selection import StratifiedKFold
from sklearn.metrics import precision_score, recall_score
from MLiquidBiopsy import MLiquidBiopsy, MLiquidBiopsyPlot
from sklearn.feature_selection import SequentialFeatureSelector


if __name__ == "__main__":
    """
    Imports
    """
    # Samplesheet
    ss_all = pd.read_csv(
        f"~/datasets/DNAMethylationPipeline_samplesheet.csv.gz", index_col=0
    )
    pal_label = ss_all.groupby("label")["palette"].first()

    # Cols manifest
    manifest_cols = [
        "CHR_hg38", "Start_hg38", "End_hg38",
        "Strand_hg38", "SourceSeq", "UCSC_CpG_Islands_Name",
        "Relation_to_UCSC_CpG_Island", "UCSC_RefGene_Name", "UCSC_RefGene_Group"
    ]

    # Probe annotation information
    pannot = MLiquidBiopsy.read_probe_annot()

    # Methylation
    methy_b = pd.read_csv(f"~/datasets/DNAMethylationPipeline_MergedSubset_methy_b.csv")
    methy_b = methy_b[[c for c in methy_b if c in ss_all.index]]
    methy_b = methy_b.drop(columns=["8784233035_R03C02"])

    # Drop NaNs
    methy_b = methy_b.dropna()

    # Drop probes not in hg38 lift annotation
    methy_b = methy_b.loc[methy_b.index.isin(pannot.index)]

    # Dimension reduction
    methy_pcs, methy_umap, methy_vexp = MLiquidBiopsy.dimension_reduction(methy_b.T, ss_all)

    MLiquidBiopsyPlot.dimension_reduction_plot(
        "PC1", "PC2", "label", methy_pcs, vexp=methy_vexp, palette=pal_label
    )
    plt.savefig(f"reports/PredMultiClass_Merged_PCA_methy.pdf", bbox_inches="tight", dpi=600)
    plt.close("all")

    MLiquidBiopsyPlot.dimension_reduction_plot(
        "Dim1", "Dim2", "label", methy_umap, method="UMAP", palette=pal_label
    )
    plt.savefig(f"reports/PredMultiClass_Merged_UMAP_methy.pdf", bbox_inches="tight", dpi=600)
    plt.close("all")

    # Drop sex chromosome
    psex = MLiquidBiopsy.probes_sex_chr()
    methy_b_ = methy_b.drop(index=psex, errors="ignore")

    # Global methylation
    methy_b_mean = methy_b_.T.mean(1)
    methy_b_mean_corr = methy_b_.T.corrwith(methy_b_mean)
    methy_b_ = methy_b_.loc[methy_b_mean_corr[methy_b_mean_corr.abs() < 0.5].index]

    # Remove CpGs not mapping to genes
    pnocds = set(pannot[pannot["UCSC_RefGene_Name"].isna()].index)
    methy_b_ = methy_b_.drop(index=pnocds, errors="ignore")

    # # Promoter methylation
    # methy_b_promoter, promoter_map = MLiquidBiopsy.calculate_promoter(methy_b_, nmin=3)
    # promoter_map_gene = promoter_map.reset_index().groupby(0)["index"].agg(lambda v: set(v))

    # Cluster methylation
    methy_b_cluster, cluster_map = MLiquidBiopsy.probes_cluster(methy_b_, nmin=3)
    cluster_map_cpg = cluster_map.reset_index().groupby("cluster")["IlmnID"].agg(set)

    """
    ML assemble matrices
    """
    # Tissue
    ss_ftypes = [
        "Primary Tumor - Liver",
        "cfDNA HCC and cirrhosis",

        "Cirrhosis + EtOH",
        "Cirrhosis + HCV",
        "cfDNA cirrhosis only",

        "Solid Tissue Normal - Liver",
    ]
    ss = ss_all[ss_all["label"].isin(ss_ftypes)]

    X = methy_b_cluster[[c for c in methy_b_cluster if c in ss.index]].copy().T

    def tissue_multi_class(stype):
        if stype in ["Primary Tumor - Liver", "cfDNA HCC and cirrhosis", "HCC + EtOH", "HCC + HCV"]:
            return "HCC"

        elif stype in ["Solid Tissue Normal - Liver", "cfDNA healthy pooled", "cfDNA healthy"]:
            return "Healthy"

        elif stype in [
            "Cirrhosis + EtOH",
            "Cirrhosis + HBV",
            "Cirrhosis + HCV",
            "Cirrhosis + AATD",
            "Cirrhosis + Other",
            "cfDNA cirrhosis only",
        ]:
            return "Cirrhosis"

        else:
            return "Other"


    pal_labels = pd.Series(sns.color_palette("Set1", n_colors=3).as_hex(), index=["Cirrhosis", "HCC", "Healthy"])

    Y = ss.loc[X.index, "label"].apply(tissue_multi_class)

    ss = ss.loc[Y.index]

    """
    Dimension reduction
    """
    X_pcs, X_umap, X_vexp = MLiquidBiopsy.dimension_reduction(X, ss)

    MLiquidBiopsyPlot.dimension_reduction_plot(
        "PC1", "PC2", "label", X_pcs, vexp=X_vexp, palette=pal_label
    )
    plt.savefig(f"reports/PredMultiClass_Merged_PCA_X.pdf", bbox_inches="tight", dpi=600)
    plt.close("all")

    MLiquidBiopsyPlot.dimension_reduction_plot(
        "Dim1", "Dim2", "label", X_umap, method="UMAP", palette=pal_label
    )
    plt.savefig(f"reports/PredMultiClass_Merged_UMAP_X.pdf", bbox_inches="tight", dpi=600)
    plt.close("all")

    """
    Classifier
    """
    y_features, y_eval, y_pred, y_nfeatures = [], [], [], []

    for r_idx in range(100):
        # Undersample
        X_ = pd.concat(
            [
                X.loc[ss.query("label == 'Primary Tumor - Liver'").sample(22).index],
                X.loc[ss.query("label == 'cfDNA HCC and cirrhosis'").sample(22).index],
                X.loc[ss.query("label == 'Cirrhosis + HCV'").sample(11).index],
                X.loc[ss.query("label == 'Cirrhosis + EtOH'").sample(11).index],
                X.loc[ss.query("label == 'cfDNA cirrhosis only'").sample(22).index],
                X.loc[ss.query("label == 'Solid Tissue Normal - Liver'").sample(44).index],
            ]
        ).copy()
        Y_ = Y.loc[X_.index].copy()

        # Stratified K-fold
        y_cv = StratifiedKFold(shuffle=True, n_splits=5).split(X_, Y_)
        # y_cv = StratifiedShuffleSplit(test_size=0.3, n_splits=10).split(X_, Y_)

        # Objects to store results
        for cv_idx, (idx_train, idx_test) in enumerate(y_cv):
            # Misc
            fdr_thres, diff_thres = 0.01, 0.2

            # ML CV matrices
            cv_x_train, cv_y_train = X_.iloc[idx_train], Y_.iloc[idx_train]
            cv_x_test, cv_y_test = X_.iloc[idx_test], Y_.iloc[idx_test]

            # ANOVA
            ftype_cpgs = []
            for dtypes in [["HCC", "Cirrhosis"], ["HCC", "Healthy"]]:
                dtsamples = list(cv_y_train[cv_y_train.isin(dtypes)].index)
                dtanova = MLiquidBiopsy.anova(
                    cv_x_train.loc[dtsamples],
                    cv_y_train.loc[dtsamples],
                    force_multiclass=True
                )
                ftype_cpgs.append(set(dtanova.query(f"fdr < {fdr_thres}").index))
            ftype_cpgs = set.intersection(*ftype_cpgs)
            y_nfeatures.append(ftype_cpgs)
            LOG.info(f"{r_idx} {cv_idx}: #(CpGs)={len(ftype_cpgs)}")

            if len(ftype_cpgs) < 10:
                continue

            cv_x_train = cv_x_train.reindex(index=cv_y_train.index, columns=ftype_cpgs)
            cv_x_test = cv_x_test.reindex(index=cv_y_test.index, columns=ftype_cpgs)

            # Classifier
            clf = MLiquidBiopsy.get_classifier(ctype="LinearSVC", C=1).fit(cv_x_train, cv_y_train)

            # Eval
            clf_coefs = pd.Series((clf.coef_ != 0).sum(1), index=clf.classes_)
            clf_eval_train = MLiquidBiopsy.eval_pred(cv_y_train, clf.predict(cv_x_train), as_str=False)
            clf_eval_test = MLiquidBiopsy.eval_pred(cv_y_test, clf.predict(cv_x_test), as_str=False)

            LOG.info(f"{r_idx} {cv_idx}: Non-zero features={clf_coefs}")
            LOG.info(f"{r_idx} {cv_idx}: Train={'; '.join([f'{m}={s:.2f}' for m, s in clf_eval_train.items()])}")
            LOG.info(f"{r_idx} {cv_idx}: Test={'; '.join([f'{m}={s:.2f}' for m, s in clf_eval_test.items()])}")

            # Store results
            clf_features = pd.DataFrame(clf.coef_, index=clf.classes_, columns=ftype_cpgs).T
            clf_features = (clf_features.unstack().replace(0, np.nan).dropna().reset_index())
            clf_features.columns = ["ftype", "feature", "coef"]
            y_features.append(clf_features)

            clf_eval_test["etype"] = "test"
            y_eval.append(clf_eval_test)

            clf_eval_train["etype"] = "train"
            y_eval.append(clf_eval_train)

            cv_y_test_pred = pd.Series(clf.predict(cv_x_train), index=cv_x_train.index)
            y_pred.append(cv_y_test_pred)

    y_features = pd.concat(y_features, ignore_index=True)
    y_features.to_csv(f"reports/PredMultiClass_Merged_y_features.csv", index=False)

    y_eval = pd.DataFrame(y_eval)
    y_eval.to_csv(f"reports/PredMultiClass_Merged_y_eval.csv", index=False)

    """
    Recursive feature selection
    """
    best_features = y_features.groupby(["feature", "ftype"])["coef"].agg("count")
    best_features = set(best_features[best_features >= 5].reset_index()["feature"])

    sfs_eval = []
    for n_feat in [5, 10, 15, 20, 25, 30, 35, 40]:

        for r_idx in range(10):
            LOG.info(f"N features={n_feat}")

            # Train
            X_train_ = pd.concat(
                [
                    X.loc[ss.query("label == 'Primary Tumor - Liver'").sample(22).index],
                    X.loc[ss.query("label == 'cfDNA HCC and cirrhosis'").sample(22).index],
                    X.loc[ss.query("label == 'Cirrhosis + HCV'").sample(11).index],
                    X.loc[ss.query("label == 'Cirrhosis + EtOH'").sample(11).index],
                    X.loc[ss.query("label == 'cfDNA cirrhosis only'").sample(22).index],
                    X.loc[ss.query("label == 'Solid Tissue Normal - Liver'").sample(44).index],
                ]
            ).copy()[best_features]
            Y_train_ = Y.loc[X_train_.index].copy()

            # Test
            test_samples = set(ss_all[ss_all["label"].isin([
                "Cirrhosis + HBV",
                "Cirrhosis + Other",
                "Cirrhosis + AATD",
                "HCC + EtOH",
                "HCC + HCV",
                "Solid Tissue Normal - Liver"
            ])].index)
            X_test_ = methy_b_cluster.reindex(columns=test_samples, index=best_features).T.dropna()
            Y_test_ = ss_all.loc[X_test_.index, "label"].apply(tissue_multi_class)

            # Feature selection
            sfs = SequentialFeatureSelector(
                estimator=MLiquidBiopsy.get_classifier("LinearSVC_L2"),
                n_features_to_select=n_feat,
                direction="forward",
                n_jobs=-1
            ).fit(X_train_, Y_train_)

            sfs_features = set(X_train_.loc[:, sfs.support_])

            # Evaluate
            clf = MLiquidBiopsy.get_classifier("LinearSVC_L2").fit(X_train_[sfs_features], Y_train_)

            sfs_eval_train = MLiquidBiopsy.eval_pred(
                Y_train_, clf.predict(X_train_[sfs_features]), as_str=False
            )

            sfs_eval_test = MLiquidBiopsy.eval_pred(
                Y_test_, clf.predict(X_test_[sfs_features]), as_str=False
            )

            # Store
            sfs_eval_train["etype"] = "train"
            sfs_eval_train["n_feat"] = n_feat
            sfs_eval_train["features"] = sfs_features
            sfs_eval.append(sfs_eval_train)

            sfs_eval_test["etype"] = "test"
            sfs_eval_test["n_feat"] = n_feat
            sfs_eval_test["features"] = sfs_features
            sfs_eval.append(sfs_eval_test)

    sfs_eval = pd.DataFrame(sfs_eval)
    sfs_eval.to_csv(f"reports/PredMultiClass_Merged_sfs_eval.csv", index=False)
    # sfs_eval = pd.read_csv(f"reports/PredMultiClass_Merged_sfs_eval.csv")

    # Evaluation boxplots
    pal = dict(train=CrispyPlot.PAL_DTRACE[1], test=CrispyPlot.PAL_DTRACE[2])

    fig, axs = plt.subplots(2, 1, figsize=(2, 3), sharey="all", sharex="all")

    for i, y_var in enumerate(["recall", "precision"]):
        sns.boxplot(
            x="n_feat",
            y=y_var,
            hue="etype",
            data=sfs_eval,
            palette=pal,
            hue_order=["train", "test"],
            orient="v",
            notch=False,
            boxprops=dict(linewidth=0.3),
            whiskerprops=dict(linewidth=0.3),
            medianprops=CrispyPlot.MEDIANPROPS,
            flierprops=CrispyPlot.FLIERPROPS,
            showcaps=False,
            saturation=1.0,
            ax=axs[i],
        )

        axs[i].grid(True, ls="-", lw=0.1, alpha=1.0, zorder=0)

        axs[i].set_xlabel("Top CpG clusters" if i == 1 else "")
        axs[i].set_ylabel(y_var)
        axs[i].set_title("Sequential Feature Selection" if i == 0 else "")

        axs[i].legend(
            frameon=False,
            title="",
            prop={"size": 5},
            loc=4,
        )

    plt.savefig(
        f"reports/PredMultiClass_Merged_sfs_boxplots.pdf", bbox_inches="tight", dpi=600
    )
    plt.close("all")

    # Histogram number of features
    plot_df = [len(f) for f in y_nfeatures]

    fig, ax = plt.subplots(1, 1, figsize=(2, 1.5), sharex="all", sharey="all")

    sns.distplot(
        plot_df,
        hist=True,
        kde=False,
        bins=15,
        hist_kws=dict(linewidth=0, alpha=1),
        color=CrispyPlot.PAL_DTRACE[2],
        ax=ax,
    )

    ax.grid(True, ls="-", lw=0.1, alpha=1.0, zorder=0)
    ax.set_xlabel("Differentially methylated clusters")
    ax.set_ylabel("Counts")
    plt.savefig(
        f"reports/PredMultiClass_Merged_anova_nfeatures_distplot.pdf", bbox_inches="tight", dpi=600
    )
    plt.close("all")

    # Evaluation barplot
    plot_df = pd.melt(y_eval, id_vars=["etype"])

    pal = dict(train=CrispyPlot.PAL_DTRACE[1], test=CrispyPlot.PAL_DTRACE[2])

    _, ax = plt.subplots(1, 1, figsize=(2, 1.5), sharex="all", sharey="all")

    sns.barplot(
        x="value",
        y="variable",
        hue="etype",
        data=plot_df,
        lw=0,
        palette=pal,
        saturation=1,
        ax=ax,
    )

    ax.grid(True, ls="-", lw=0.1, alpha=1.0, zorder=0)
    ax.set_xlabel("Metric")
    ax.set_ylabel("")
    ax.legend(frameon=False, title="", prop={"size": 6}, loc="center left", bbox_to_anchor=(1, 0.5))
    plt.savefig(
        f"reports/PredMultiClass_Merged_eval_barplot.pdf", bbox_inches="tight", dpi=600
    )
    plt.close("all")

    #
    plot_df = y_features.groupby(["feature", "ftype"])["coef"].agg(["mean", "count"]).reset_index()
    plot_df["count"] /= (len(y_eval) / 2)
    plot_df["mean"] = plot_df["mean"].abs()

    _, ax = plt.subplots(1, 1, figsize=(2, 2), sharex="all", sharey="all")

    sns.scatterplot(
        x="count",
        y="mean",
        hue="ftype",
        s=3,
        lw=0,
        alpha=0.8,
        data=plot_df,
        palette=pal_labels.to_dict(),
        ax=ax,
    )

    sns.scatterplot(
        x="count",
        y="mean",
        hue="ftype",
        s=8,
        edgecolor="Grey",
        lw=.3,
        alpha=0.8,
        data=plot_df.query("mean > 2 | count > 0.2"),
        palette=pal_labels.to_dict(),
        legend=False,
        ax=ax,
    )

    ax.axhline(2, ls="--", lw=0.3, c=CrispyPlot.PAL_DTRACE[1])
    ax.axvline(.2, ls="--", lw=0.3, c=CrispyPlot.PAL_DTRACE[1])

    ax.grid(True, ls="-", lw=0.1, alpha=1.0, zorder=0)
    ax.set_xlabel("Feature frequency (%)")
    ax.set_ylabel("Feature weight (abs)")

    ax.legend(frameon=False, title="", prop={"size": 4})
    plt.savefig(
        f"reports/PredMultiClass_Merged_feature_frequency_scatter.pdf", bbox_inches="tight", dpi=600
    )
    plt.close("all")

    # Top features venn
    top_features = y_features.groupby(["feature", "ftype"])["coef"].agg(["mean", "count"]).reset_index()
    top_features["count"] /= (len(y_eval) / 2)
    top_features = top_features.query("abs(mean) > 2 | count > 0.2")
    top_features["cpgs"] = [list(cluster_map_cpg[f]) for f in top_features["feature"]]

    plot_df = top_features.groupby("ftype")["feature"].agg(set).to_dict()

    _, ax = plt.subplots(1, 1, figsize=(1.5, 1.5))

    venn3(plot_df.values(), set_labels=plot_df.keys(), set_colors=pal_labels[plot_df.keys()], alpha=0.8, ax=ax)
    venn3_circles(plot_df.values(), linestyle='--', linewidth=.5, color="k")

    plt.savefig(
        f"reports/PredMultiClass_Merged_top_features_venn.pdf", bbox_inches="tight", dpi=600
    )
    plt.close("all")

    """
    Annotate top features
    """
    top_features_cpgs = top_features.explode("cpgs").reset_index(drop=True)
    top_features_cpgs = pd.concat([
        top_features_cpgs,
        pannot.loc[top_features_cpgs["cpgs"], manifest_cols].reset_index(drop=True),
    ], axis=1).sort_values("count", ascending=False)
    top_features_cpgs.to_csv(f"reports/PredMultiClass_Merged_top_features_cpg_sites.csv", index=False)
    # top_features_cpgs = pd.read_csv(f"reports/PredMultiClass_Merged_top_features_cpg_sites.csv")

    LOG.info(
        f"#CpG clusters & sites: {len(set(top_features['feature']))}, {len(set(top_features_cpgs['cpgs']))}"
    )

    # Methylation regions
    for f_cluster in top_features["feature"].unique():
        features = cluster_map_cpg[f_cluster]
        MLiquidBiopsyPlot.methylation_region(
            list(features)[0],
            methy_b[Y.index],
            pannot,
            ss,
            hue_order=ss_ftypes,
            genomic_range=1e3,
            feature_highlights=features,
        )
        plt.savefig(
            f"reports/PredMultiClass_Merged_methylation_region_{f_cluster}.pdf",
            bbox_inches="tight",
            dpi=600,
        )
        plt.close("all")

    """
    Final model
    """
    # Matrices
    ss_cfdna = ss_all[
        ss_all["label"].isin(
            [
                "cfDNA HCC and cirrhosis",
                "cfDNA cirrhosis only",
                "cfDNA healthy pooled",
                "cfDNA healthy",
                # "cfDNA",
            ]
        )
    ]

    X = methy_b_cluster[[c for c in methy_b_cluster if c in ss_cfdna.index]].copy().T
    Y = ss_all.loc[X.index, "label"].apply(tissue_multi_class)

    # Feature sets
    features = ["TopDiff"] + list(MLiquidBiopsy.TOP_CPGS)
    features_pal = sns.color_palette("tab20c").as_hex()
    features_pal = pd.Series(
        [features_pal[0]] + features_pal[4:4 + len(features) - 1], index=features
    )

    # Model
    y_cv = StratifiedKFold(shuffle=True, n_splits=5).split(X, Y)

    final_res = dict()
    final_pred = {k: [] for k in features}

    for idx_train, idx_test in y_cv:
        # ML CV matrices
        cv_x_train, cv_y_train = X.iloc[idx_train], Y.iloc[idx_train]
        cv_x_test, cv_y_test = X.iloc[idx_test], Y.iloc[idx_test]

        for ftype in features:
            # Features
            if ftype in MLiquidBiopsy.TOP_CPGS:
                ftype_cpgs = MLiquidBiopsy.TOP_CPGS[ftype].intersection(methy_b.index)

            else:
                ftype_cpgs = set(top_features_cpgs["cpgs"])

            ftype_x_train = methy_b.reindex(index=ftype_cpgs, columns=cv_y_train.index).T
            ftype_x_test = methy_b.reindex(index=ftype_cpgs, columns=cv_y_test.index).T

            # Classifier
            clf = MLiquidBiopsy.get_classifier(ctype="LinearSVC", C=2).fit(ftype_x_train, cv_y_train)

            # Store results
            final_pred[ftype].append(pd.Series(clf.predict(ftype_x_test), index=cv_y_test.index))

    final_res["y_true"] = Y.copy()
    final_res["y_pred"] = pd.DataFrame({ftype: pd.concat(final_pred[ftype]) for ftype in features}).loc[Y.index]

    # Eval plots
    plot_df = MLiquidBiopsyPlot.eval_barplot(dict(MultiClass=final_res), palette=features_pal)
    plt.savefig(f"reports/PredMultiClass_Merged_barplot.pdf", bbox_inches="tight", dpi=600)
    plt.close("all")

    MLiquidBiopsyPlot.eval_confusion_matrices(dict(MultiClass=final_res))
    plt.savefig(
        f"reports/PredMultiClass_Merged_confusion_matrix.pdf",
        bbox_inches="tight",
        dpi=600,
    )
    plt.close("all")

    """
    Feature score
    """
    top_features = top_features_cpgs.groupby("feature").agg(dict(
        ftype="first", mean="first", count="first", cpgs=set
    ))

    def calculate_score(clf_type):
        weights = top_features[top_features["ftype"].isin(clf_type)]
        return pd.DataFrame(
            [methy_b.loc[f].mean() * w for f, w in weights[["cpgs", "mean"]].values]
        ).sum()

    plot_df = pd.concat(
        [
            calculate_score(["HCC"]).rename("HCC"),
            calculate_score(["Cirrhosis"]).rename("Cirrhosis"),
            calculate_score(["Healthy"]).rename("Healthy"),
            ss_all["label"],
        ],
        axis=1,
    )

    plot_df = plot_df[
        plot_df["label"].isin(
            [
                "cfDNA HCC and cirrhosis",
                "cfDNA cirrhosis only",
                "cfDNA healthy",
                "cfDNA healthy pooled",
                "Cirrhosis + HBV",
                "Cirrhosis + Other",
                "Cirrhosis + AATD",
                "HCC + EtOH",
                "HCC + HCV",
                "Solid Tissue Normal - Liver"
            ]
        )
    ]

    plot_df["class"] = plot_df["label"].apply(tissue_multi_class).values

    for x_var, y_var, color in [("HCC", "Cirrhosis", "#5192BF"), ("HCC", "Healthy", "#9A4973")]:
        plot_df_subset = plot_df[plot_df["class"].isin([x_var, y_var])].dropna(subset=[x_var, y_var])
        plot_df_subset["y_true"] = (plot_df_subset["class"] == x_var).astype(int).values

        #
        _, t, r, p = MLiquidBiopsyPlot.recall_precision_curve(plot_df_subset["y_true"], plot_df_subset["HCC"], color=color)
        plt.savefig(
            f"reports/PredMultiClass_Merged_score_scatter_{x_var}_{y_var}_recall_curve.pdf",
            bbox_inches="tight",
            dpi=600,
        )
        plt.close("all")

        #
        ax = GIPlot.gi_tissue_plot(
            x_var,
            y_var,
            plot_df=plot_df,
            hue="label",
            plot_reg=False,
            annot=False,
            pal=pal_label.to_dict(),
        )

        ax.axvline(
            t,
            lw=0.5,
            ls="--",
            c=color,
        )
        ax.text(t + 0.1, ax.get_ylim()[1] - 0.1, f"Recall={r:.2f}\nPrecision={p:.2f}", va="top", color=color, fontsize=4, zorder=3)

        ax.set_xlabel(f"HepatoDetect molecular score")
        ax.set_ylabel(f"{y_var} score")

        plt.savefig(
            f"reports/PredMultiClass_Merged_score_scatter_{x_var}_{y_var}.pdf",
            bbox_inches="tight",
            dpi=600,
        )
        plt.close("all")

    """
    CpG sites for validation
    """
    # top_features_cpgs = pd.read_csv(f"reports/PredMultiClass_Merged_top_features_cpg_sites.csv")

    for dtype in ["Small_Set", "Middle_Set", "Large_Set"]:
        # Select trained features
        if dtype in ["Small_Set"]:
            val_top_features = top_features_cpgs.query("count > 0.2")

        else:
            val_top_features = top_features_cpgs.copy()

        # Merge with independent sets of features
        val_features = pd.concat([
            pd.DataFrame(dict(cpg=list(val_top_features["cpgs"].unique()), db="TopFeatures")),
            MLiquidBiopsy.independent_features(methy_b),
        ], ignore_index=True)

        if dtype in ["Large_Set"]:
            val_features = val_features[
                val_features["db"].isin([
                    "Hlady et al.", "Villanueva et al.", "Cheng et al.",
                    "Xu et al. prognosis", "US-2019300965-A1", "US-2020263256-A1", "TopFeatures"
                ])
            ]

        else:
            val_features = val_features[
                val_features["db"].isin(["Hlady et al.", "Villanueva et al.", "TopFeatures"])
            ]

        # Add specific sets
        specific_clusters = ["chr17_Cluster184587"]
        specific_clusters_cpgs = list(set.union(*cluster_map_cpg[specific_clusters].values))

        val_features = pd.concat([
            pd.DataFrame(dict(cpg=specific_clusters_cpgs, db="TopFeatures")),
            val_features,
        ], ignore_index=True)

        # Remove duplicate CpG and annotate
        val_features = val_features.groupby("cpg").first().reset_index()
        val_features = val_features[val_features["cpg"].isin(methy_b.index)]

        val_features = pd.concat([
            val_features,
            pannot.loc[val_features["cpg"]][manifest_cols].reset_index(drop=True),
        ], axis=1)

        val_features["cluster"] = cluster_map.reindex(val_features["cpg"]).values
        val_features["cluster"] = val_features["cluster"].fillna(val_features["cpg"]).values
        val_features.to_csv(f"reports/PredMultiClass_Merged_validation_cpg_sites_{dtype}.csv", index=False)

        # Group by clusters
        group_func = dict(
            cpg=set, db=set, CHR_hg38="first", Start_hg38=min, End_hg38=max, Strand_hg38=set
        )

        val_features_by_region = val_features.groupby("cluster").agg(group_func)

        val_features_by_region["Strand_hg38"] = val_features_by_region["Strand_hg38"].apply(list)
        val_features_by_region = val_features_by_region.explode("Strand_hg38").reset_index()

        val_features_by_region["size"] = val_features_by_region.eval("End_hg38 - Start_hg38").astype(int).values
        val_features_by_region.to_csv(f"reports/PredMultiClass_Merged_validation_cpg_clusters_{dtype}.csv", index=False)

        LOG.info(f"{dtype}: CpGs={len(val_features)}; Clusters={len(val_features_by_region)}")
```
