## Supplementary File 1 for "DNA methylation fingerprint for the diagnosis and monitoring of hepatocellular carcinoma from tissue and liquid biopsies": ParseSamplesheet.py

```
import numpy as np
import pandas as pd
import seaborn as sns

colors = sns.color_palette("tab20b").as_hex() + sns.color_palette("tab20c").as_hex()

"""
Whole blood samples
"""

mnf_blood_ss = pd.read_csv(
    f"~/datasets/Methylation_450k_GSE40279/E-GEOD-40279.sdrf.txt", sep="\t"
)
mnf_blood_ss["Comment [Sample_source_name]"] = mnf_blood_ss[
    "Comment [Sample_source_name]"
].apply(lambda v: int(v[1:]))

mnf_blood_map = pd.read_csv(
    f"~/datasets/Methylation_450k_GSE40279/GSE40279_sample_key.txt.gz",
    sep="\t",
    header=None,
    index_col=1,
)
mnf_blood_ss["file"] = mnf_blood_map.loc[mnf_blood_ss["Comment [Sample_source_name]"]][
    2
].values

mnf_blood = mnf_blood_ss[
    [
        "FactorValue [AGE Y]",
        "FactorValue [SEX]",
        "FactorValue [ETHNICITY]",
        "Characteristics [organism part]",
        "file",
    ]
]
mnf_blood = mnf_blood.rename(
    columns={
        "FactorValue [AGE Y]": "age_at_index",
        "FactorValue [SEX]": "gender",
        "FactorValue [ETHNICITY]": "ethnicity",
        "Characteristics [organism part]": "sample_type",
        "file": "filename",
    }
)
mnf_blood["sample_type"] = mnf_blood["sample_type"].apply(lambda v: v.title())

mnf_blood["palette"] = colors[12]
mnf_blood["label"] = "Whole blood"

mnf_blood["dataset_id"] = "GSE40279"
mnf_blood["dataset_color"] = "#DA4453"


"""
TCGA samples
"""

mnf_tcga = (
    pd.concat(
        [
            pd.read_csv(
                f"~/datasets/Methylation_450k_TCGA_LIHC/gdc_manifest_20201013_071752.txt",
                sep="\t",
            ),
            pd.read_csv(
                f"~/datasets/Methylation_450k_TCGA_normals/gdc_manifest.2020-10-14.normals.txt",
                sep="\t",
            ),
        ],
        ignore_index=True,
    )
    .drop(columns=["md5", "size", "state"])
    .drop_duplicates()
    .set_index("id")
)
mnf_tcga["filename"] = mnf_tcga["filename"].apply(lambda v: "_".join(v.split("_")[:2]))

barcodes_tcga = pd.read_csv(
    f"~/datasets/Methylation_450k_TCGA_normals/TCGA_UUID_barcode.csv", index_col=0
)
mnf_tcga["barcode"] = barcodes_tcga.loc[mnf_tcga.index]

clinical_tcga = pd.read_csv(
    f"~/datasets/Methylation_450k_TCGA_normals/clinical.cases_selection.2020-10-26/clinical.tsv",
    sep="\t",
)
clinical_tcga = clinical_tcga[
    [
        "case_id",
        "case_submitter_id",
        "project_id",
        "age_at_index",
        "ajcc_pathologic_stage",
        "gender",
        "ethnicity",
        "primary_diagnosis",
        "tissue_or_organ_of_origin",
    ]
]
clinical_tcga = clinical_tcga.drop_duplicates().set_index("case_submitter_id")

sample_tcga = pd.read_csv(
    f"~/datasets/Methylation_450k_TCGA_normals/biospecimen.cases_selection.2020-10-26/sample.tsv",
    sep="\t",
    index_col="sample_submitter_id",
)

mnf_tcga = (
    pd.concat(
        [
            mnf_tcga,
            sample_tcga.reindex(mnf_tcga["barcode"].apply(lambda v: v[:16]))[
                ["sample_type"]
            ].set_index(mnf_tcga.index),
            clinical_tcga.reindex(
                mnf_tcga["barcode"].apply(lambda v: v[:12]).values
            ).set_index(mnf_tcga.index),
        ],
        axis=1,
    )
    .dropna()
    .reset_index(drop=True)
    .drop_duplicates()
)

mnf_tcga["primary_diagnosis"] = mnf_tcga["primary_diagnosis"].apply(
    lambda v: v.split(",")[0]
)
mnf_tcga["tissue_or_organ_of_origin"] = mnf_tcga["tissue_or_organ_of_origin"].apply(
    lambda v: v.split(",")[0]
)


def mnf_tcga_label(stype, torigin):
    if (str(stype) == "Primary Tumor") & (torigin == "Liver"):
        label, color = f"{stype} - {torigin}", colors[20]

    elif (str(stype) == "Recurrent Tumor") & (torigin == "Liver"):
        label, color = f"{stype} - {torigin}", colors[21]

    elif (str(stype) == "Solid Tissue Normal") & (torigin == "Liver"):
        label, color = f"{stype} - {torigin}", colors[28]

    elif (str(stype) == "Solid Tissue Normal") & (torigin not in ["Liver"]):
        label, color = f"{stype} - Other", colors[30]

    else:
        label, color = "Other", colors[39]

    return label, color


mnf_tcga["label"], mnf_tcga["palette"] = list(
    zip(
        *[
            mnf_tcga_label(s, t)
            for s, t in mnf_tcga[["sample_type", "tissue_or_organ_of_origin"]].values
        ]
    )
)

mnf_tcga["dataset_id"] = "TCGA"
mnf_tcga["dataset_color"] = "#4A89DC"

tcga_purity = pd.read_excel(f"~/datasets/Methylation_450k_TCGA_LIHC/41467_2015_BFncomms9971_MOESM1236_ESM.xlsx", index_col=0)
mnf_tcga["purity_CPE"] = tcga_purity.reindex(mnf_tcga["barcode"].apply(lambda v: v[:16]))["CPE"].values


"""
cfDNA
"""
mnf_cfdna = pd.read_csv(
    "~/datasets/Methylation_450k_GSE122126/filelist.txt", sep="\t"
).query("Type == 'IDAT'")
mnf_cfdna["gsm"] = mnf_cfdna["Name"].apply(lambda v: v.split("_")[0])
mnf_cfdna["filename"] = mnf_cfdna["Name"].apply(lambda v: v.split(".")[0][:-4])

mnf_cfdna_type = pd.read_csv(
    "~/datasets/Methylation_450k_GSE122126/sample_type_map.tsv", sep="\t", index_col=0
)["type"]
mnf_cfdna["sample_type"] = mnf_cfdna_type[mnf_cfdna["gsm"]].values


def mnf_Moss_cfdna(stype):
    if str(stype).startswith("hepatocytes_"):
        label, color = f"Hepatocytes", colors[8]

    elif str(stype).startswith("colon_epithelial_cells_"):
        label, color = f"Colon epithelial cells", colors[10]

    elif str(stype).startswith("pancreatic_"):
        label, color = f"Pancreatic cells", colors[11]

    elif str(stype).startswith("adipocytes_"):
        label, color = f"Adipocytes", colors[27]

    elif "_healthy_" in str(stype):
        label, color = f"cfDNA healthy", colors[28]

    elif "_pool" in str(stype):
        label, color = f"cfDNA healthy pooled", colors[29]

    elif str(stype).startswith("cfDNA_"):
        label, color = f"cfDNA", colors[4]

    elif not str(stype).startswith("cfDNA_"):
        label, color = f"Other cell types", colors[38]

    else:
        label, color = "Other", colors[39]

    return label, color


mnf_cfdna["label"], mnf_cfdna["palette"] = list(
    zip(*[mnf_Moss_cfdna(s) for s in mnf_cfdna["sample_type"]])
)

mnf_cfdna["dataset_id"] = "GSE122126"
mnf_cfdna["dataset_color"] = "#F6BB42"


"""
cfDNA HCC
"""
mnf_cfdna_hcc = pd.read_csv(
    "~/datasets/Methylation_450k_GSE129374/GSE129374_series_matrix.txt",
    sep="\t",
    index_col="Sample_title",
).T
mnf_cfdna_hcc = (
    mnf_cfdna_hcc["Sample_supplementary_file"]
    .iloc[:, 0]
    .apply(lambda v: v.split("/")[-1].replace("_Grn.idat.gz", ""))
    .reset_index()
)
mnf_cfdna_hcc = mnf_cfdna_hcc.rename(
    columns=dict(index="sample_type", Sample_supplementary_file="filename")
)


def mnf_hcc_cfdna(stype):
    if str(stype).endswith("_Cirrhosis_alone"):
        label, color = f"cfDNA cirrhosis only", colors[0]

    elif str(stype).endswith("_Cirrhosis_with_HCC"):
        label, color = f"cfDNA HCC and cirrhosis", colors[6]

    else:
        label, color = "Other", colors[39]

    return label, color


mnf_cfdna_hcc["label"], mnf_cfdna_hcc["palette"] = list(
    zip(*[mnf_hcc_cfdna(s) for s in mnf_cfdna_hcc["sample_type"]])
)

mnf_cfdna_hcc["dataset_id"] = "GSE129374"
mnf_cfdna_hcc["dataset_color"] = "#D770AD"


"""
Liver samples
"""
mnf_liver = pd.read_csv(
    "~/datasets/Methylation_450k_GSE60753/Liver_450k_iDaT_sample_sheet_02-04-20.csv"
)
mnf_liver["sample_type"] = mnf_liver["Disease"] + " + " + mnf_liver["Etiology"]
mnf_liver = mnf_liver.rename(columns=dict(Full_ID="filename"))


def mnf_liver_label(stype):
    if str(stype).startswith("Plated_hepatocyte"):
        label, color = f"Plated hepatocytes", colors[9]

    elif str(stype) in ["Normal + Normal", "Normal Liver + Normal"]:
        label, color = f"Liver normal", colors[29]

    elif str(stype) == "Cirrhosis + HBV":
        label, color = f"Cirrhosis + HBV", colors[16]

    elif str(stype) == "Cirrhosis + HCV":
        label, color = f"Cirrhosis + HCV", colors[17]

    elif str(stype) == "Cirrhosis + EtOH":
        label, color = f"Cirrhosis + EtOH", colors[18]

    elif str(stype).startswith("Cirrhosis + "):
        label, color = f"Cirrhosis + Other", colors[19]

    elif str(stype) == "HCC + HCV":
        label, color = f"HCC + HCV", colors[32]

    elif str(stype) == "HCC + EtOH":
        label, color = f"HCC + EtOH", colors[33]

    elif str(stype).endswith("HCC + "):
        label, color = f"HCC + Other", colors[35]

    elif str(stype) in [
        "Tumor + HCC cell line",
        "Tumor + cell line",
        "Tumor + cell lines",
    ]:
        label, color = f"HCC cell line", colors[24]

    elif str(stype) == "Tumor + High BMI":
        label, color = f"Tumor + High BMI", colors[25]

    else:
        label, color = "Other", colors[39]

    return label, color


mnf_liver["label"], mnf_liver["palette"] = list(
    zip(*[mnf_liver_label(s.split("-")[0]) for s in mnf_liver["sample_type"]])
)

mnf_liver["dataset_id"] = "GSE60753"
mnf_liver["dataset_color"] = "#37BC9B"


"""
Obese liver samples
"""
mnf_obese_liver = pd.read_csv(
    "~/datasets/Methylation_450k_GSE61446/GSE61446_series_matrix.txt",
    sep="\t",
    index_col="Sample_title",
).T
mnf_obese_liver["filename"] = (
    mnf_obese_liver["Sample_geo_accession"]
    + "_"
    + mnf_obese_liver["Sample_description_2"]
)
mnf_obese_liver = mnf_obese_liver.reset_index().rename(
    columns=dict(index="sample_type")
)
mnf_obese_liver["label"] = "Liver obese"
mnf_obese_liver["palette"] = colors[31]

mnf_obese_liver["dataset_id"] = "GSE61446"
mnf_obese_liver["dataset_color"] = "#E9573F"


"""
AATD liver samples
"""
mnf_aatd_liver = pd.read_csv(
    "~/datasets/Methylation_450k_GSE119100/GSE119100_series_matrix.txt",
    sep="\t",
    index_col="Sample_title",
).T
mnf_aatd_liver["filename"] = mnf_aatd_liver["Sample_supplementary_file_grn"].apply(
    lambda v: v.split("/")[-1].replace("_Grn.idat.gz", "")
)
mnf_aatd_liver = mnf_aatd_liver.reset_index().rename(columns=dict(index="sample_type"))


def mnf_aatd_label(stype):
    if "DNA from AATD patient" in str(stype):
        label, color = f"AATD", colors[22]

    elif "DNA from cirrhotic  AATD" in str(stype):
        label, color = f"Cirrhosis + AATD", colors[23]

    else:
        label, color = "Other", colors[39]

    return label, color


mnf_aatd_liver["label"], mnf_aatd_liver["palette"] = list(
    zip(*[mnf_aatd_label(s.split("-")[0]) for s in mnf_aatd_liver["sample_type"]])
)

mnf_aatd_liver["dataset_id"] = "GSE119100"
mnf_aatd_liver["dataset_color"] = "#967ADC"


"""
Merge
"""

mnf_cols = [
    "filename",
    "sample_type",
    "gender",
    "age_at_index",
    "ethnicity",
    "label",
    "palette",
    "barcode",
    "case_id",
    "project_id",
    "ajcc_pathologic_stage",
    "primary_diagnosis",
    "tissue_or_organ_of_origin",
    "purity_CPE",
    "dataset_id",
    "dataset_color",
]

mnf = pd.concat(
    [
        mnf_blood,
        mnf_tcga,
        mnf_cfdna,
        mnf_cfdna_hcc,
        mnf_liver,
        mnf_obese_liver,
        mnf_aatd_liver,
    ]
)[mnf_cols]
mnf = mnf[
    ~mnf["filename"].isin(
        [
            "5901393011_R01C02",
            "5901393027_R01C01",
            "5901393011_R05C01",

            "8784233035_R04C02",
            "8784233035_R05C02",
            "8784233056_R03C01",
            "8784233056_R04C01",
            "8942326007_R03C02",
            "8942326007_R04C02",
            "8942326028_R02C01",
        ]
    )
]
mnf = mnf.drop_duplicates()
mnf = mnf.replace("'--", np.nan)

mnf.to_csv("~/datasets/DNAMethylationPipeline_samplesheet.csv", index=False)
mnf.to_csv(
    "~/datasets/DNAMethylationPipeline_samplesheet.csv.gz",
    index=False,
    compression="gzip",
)
```
