## Supplementary File 1 for "DNA methylation fingerprint for the diagnosis and monitoring of hepatocellular carcinoma from tissue and liquid biopsies": SimulateCfDNA.py

```
import numpy as np
import pandas as pd
import seaborn as sns
import matplotlib.pyplot as plt
from umap import UMAP
from natsort import natsorted
from crispy.GIPlot import GIPlot
from matplotlib.lines import Line2D
from sklearn.preprocessing import StandardScaler
from LiquidBiopsy import LOG, manifest_label


class SimulateCfDNA:
    def __init__(
        self,
        beta_plasma,
        beta_hcc,
        std_plasma,
        std_hcc,
    ):
        self.beta_plasma = beta_plasma
        self.beta_hcc = beta_hcc

        self.std_plasma = std_plasma
        self.std_hcc = std_hcc

    @staticmethod
    def sample_error(beta_std):
        return pd.Series(
            np.random.normal(0, beta_std),
            index=beta_std.index,
        )

    def simulate_cfdna(self, weight_plasma, weight_hcc, add_error=True):
        index = set(self.beta_plasma.index).intersection(self.beta_hcc.index)

        # Simulate plasma cfDNA
        beta_plasma_ = self.beta_plasma.copy()

        if add_error:
            beta_plasma_ += self.sample_error(self.std_plasma / 2)

        beta_plasma_[beta_plasma_ > 1] = 1
        beta_plasma_[beta_plasma_ < 0] = 0

        # Simulate HCC cfDNA
        beta_hcc_ = self.beta_hcc.copy()

        if add_error:
            beta_hcc_ += self.sample_error(self.std_hcc / 2)

        beta_hcc_[beta_hcc_ > 1] = 1
        beta_hcc_[beta_hcc_ < 0] = 0

        # Weighted sum betas
        beta_ = weight_plasma * beta_plasma_[index] + weight_hcc * beta_hcc_[index]

        return beta_


if __name__ == "__main__":
    """
    Imports
    """
    # Methylation beta
    methy_b = pd.concat(
        [
            pd.read_csv(f"~/datasets/DNAMethylationPipeline_methy_b.csv"),
            pd.read_csv(f"~/datasets/DNAMethylationPipeline_methy_b_EPIC.csv"),
        ],
        axis=1,
    ).dropna()

    # Samplesheet
    ss = pd.read_csv(
        f"~/datasets/DNAMethylationPipeline_samplesheet.csv.gz", index_col=0
    )
    ss = pd.concat(
        [
            ss,
            pd.DataFrame(
                {
                    i: manifest_label(r["sample_type"], r["tissue_or_organ_of_origin"])
                    for i, r in ss.iterrows()
                }
            ).T,
        ],
        axis=1,
    )
    ss["stage"] = ss["ajcc_pathologic_stage"].apply(
        lambda v: v[:-1] if str(v) != "nan" and v.endswith(("A", "B", "C")) else v
    )

    # cfDNA types
    cfdna_types = pd.read_csv(
        f"~/datasets/Methylation_450k_GSE122126/cfdna_samples_classification.tsv",
        index_col=0,
        sep="\t",
    )

    cfdna_types = pd.Series(
        {
            ss.loc[i, "sample_type"]: cfdna_types.loc[i, "type"]
            if i in cfdna_types.index
            else c
            for i, c in ss[ss["label"].apply(lambda v: "cfDNA" in v)][
                "label"
            ].iteritems()
        }
    )

    """
    Healthy samples
    """
    healthy_samples = pd.read_csv(
        f"~/datasets/Methylation_450k_GSE122126/cfdna_samples_classification.tsv",
        index_col=0,
        sep="\t",
    ).query("type == 'healthy'")

    """
    HCC samples
    """
    hcc_samples = ss.query("label == 'Primary Tumor - Liver'")
    hcc_samples_choice = np.random.choice(
        hcc_samples.index, size=(10, 4), replace=False
    )

    """
    Simulate
    """
    concentrations = list(
        zip(np.arange(0.9, 1.0, 0.01), np.arange(0.01, 0.11, 0.01)[::-1])
    )

    methy_b_sim = dict()

    for i, hcc_sample in enumerate(hcc_samples_choice):
        hcc_sim = SimulateCfDNA(
            beta_plasma=methy_b[healthy_samples.index].mean(1),
            std_plasma=methy_b[healthy_samples.index].std(1),
            beta_hcc=methy_b[hcc_sample].mean(1),
            std_hcc=methy_b[hcc_sample].std(1),
        )

        for p_weight, hcc_weight in concentrations:
            methy_b_sim[
                f"simulated_{i+1}_cfDNA_HCC_{hcc_weight*100:.0f}%"
            ] = hcc_sim.simulate_cfdna(p_weight, hcc_weight, add_error=False)

    methy_b_sim = pd.DataFrame(methy_b_sim)
    methy_b_sim_samples = list(methy_b_sim)

    """
    Merge Simulated and Measured data
    """
    ss_merge = ss[
        ss["label"].isin(
            [
                "Primary Tumor - Liver",
                "Solid Tissue Normal - Liver",
                "hepatocytes",
                "Recurrent Tumor - Liver",
                "cfDNA Cirrhosis alone",
                "cfDNA Cirrhosis + HCC",
                "cfDNA",
                "cfDNA healthy",
                "cfDNA healthy pooled",
            ]
        )
    ]

    ss_sim = pd.concat(
        [
            ss_merge,
            pd.DataFrame(
                dict(sample_type=methy_b_sim_samples, tissue_or_organ_of_origin=np.nan),
                index=methy_b_sim_samples,
            ),
        ]
    )

    ss_sim = pd.DataFrame(
        {
            i: manifest_label(r["sample_type"], r["tissue_or_organ_of_origin"])
            for i, r in ss_sim.iterrows()
        }
    ).T
    ss_sim.to_csv("~/datasets/DNAMethylationPipeline_simulated_cfDNA_samplesheet.csv")

    methy_b_sim = pd.concat(
        [
            methy_b_sim,
            methy_b[ss_merge.index],
        ],
        axis=1,
    )
    methy_b_sim.to_csv("~/datasets/DNAMethylationPipeline_simulated_cfDNA_methy_b.csv")

    methy_b_sim_scaled = pd.DataFrame(
        StandardScaler().fit_transform(methy_b_sim.T),
        index=methy_b_sim.columns,
        columns=methy_b_sim.index,
    ).dropna()

    """
    UMAP
    """
    umap = UMAP()
    umap_dim = pd.DataFrame(
        umap.fit_transform(methy_b_sim_scaled),
        index=methy_b_sim_scaled.index,
        columns=["Dim1", "Dim2"],
    )

    plot_df = pd.concat([umap_dim, ss_sim], axis=1).dropna(
        subset=list(umap_dim.columns) + ["label"]
    )

    label_pal = ss_sim.groupby("label")["color"].first()

    plot_df["label"] = pd.Categorical(plot_df["label"], categories=natsorted(label_pal.index))

    ax = GIPlot.gi_tissue_plot(
        "Dim1",
        "Dim2",
        plot_df=plot_df.sort_values("label"),
        hue="label",
        plot_reg=False,
        annot=False,
        pal=label_pal,
    )

    ax.set_xlabel("UMAP Dimension 1")
    ax.set_ylabel("UMAP Dimension 2")

    plt.savefig(f"reports/SimulateCfDNA_umap.pdf", bbox_inches="tight", dpi=600)
    plt.close("all")

    """
    UMAP subset
    """
    TOP_CPGs = ["cg04645914", "cg06215569", "cg23663760", "cg13781744", "cg07610777"]

    methy_b_sim_subset = methy_b_sim.reindex(TOP_CPGs).dropna()
    methy_b_sim_subset_scaled = pd.DataFrame(
        StandardScaler().fit_transform(methy_b_sim_subset.T),
        index=methy_b_sim_subset.columns,
        columns=methy_b_sim_subset.index,
    ).dropna()

    umap_subset = UMAP()
    umap_subset_dim = pd.DataFrame(
        umap_subset.fit_transform(methy_b_sim_subset_scaled),
        index=methy_b_sim_subset_scaled.index,
        columns=["Dim1", "Dim2"],
    )

    plot_df = pd.concat([umap_subset_dim, ss_sim], axis=1).dropna(
        subset=list(umap_subset_dim.columns) + ["label"]
    )

    label_pal = ss_sim.groupby("label")["color"].first()

    plot_df["label"] = pd.Categorical(plot_df["label"], categories=natsorted(label_pal.index))

    ax = GIPlot.gi_tissue_plot(
        "Dim1",
        "Dim2",
        plot_df=plot_df.sort_values("label"),
        hue="label",
        plot_reg=False,
        annot=False,
        pal=label_pal,
    )

    for stype in ["cfDNA Cirrhosis alone", "cfDNA Cirrhosis + HCC"]:
        ax.scatter(
            plot_df.query(f"label == '{stype}'")["Dim1"],
            plot_df.query(f"label == '{stype}'")["Dim2"],
            c=label_pal[stype],
            marker="x",
            edgecolor="",
            s=10,
            alpha=0.8,
        )

    ax.set_xlabel("UMAP Dimension 1")
    ax.set_ylabel("UMAP Dimension 2")

    plt.savefig(f"reports/SimulateCfDNA_umap_subset.pdf", bbox_inches="tight", dpi=600)
    plt.close("all")

    """
    
    """
    plot_df = pd.concat([
        methy_b_sim.mean().rename("mean"),
        methy_b_sim.std().rename("std"),
        ss_sim,
    ], axis=1)

    fig, ax = plt.subplots(1, 1, figsize=(2.5, 2.5))

    ax.scatter(
        plot_df["mean"],
        plot_df["std"],
        c=plot_df["color"],
        marker="o",
        edgecolor="",
        s=5,
        alpha=0.8,
    )

    ax.legend(
        [Line2D([0], [0], color=label_pal[i], lw=4) for i in natsorted(label_pal.index)],
        natsorted(label_pal.index),
        frameon=False,
        loc="center left",
        bbox_to_anchor=(1, 0.5),
        prop={'size': 5}
    )

    ax.grid(axis="both", lw=0.1, color="#e1e1e1", zorder=0)

    ax.set_xlabel("Mean")
    ax.set_ylabel("Sandard deviation")

    plt.savefig(f"reports/SimulateCfDNA_mean_std.pdf", bbox_inches="tight", dpi=600)
    plt.close("all")
```
