## Supplementary File 1 for "DNA methylation fingerprint for the diagnosis and monitoring of hepatocellular carcinoma from tissue and liquid biopsies": TCGA450kReader.py

```
import pandas as pd

DDIR = "~/Dropbox (Ophiomics)/Ophiomics team folder/R&D/Datasets/TCGA DNA Methylation/"

data = pd.read_csv(
    f"{DDIR}/jhu-usc.edu_LIHC.HumanMethylation450.11.lvl-3.TCGA-MI-A75I-01A-11D-A32R-05.gdc_hg38.txt",
    sep="\t",
)

# Manifest
manifest = pd.read_csv(
    f"~/Projects/LiquidBiopsy/downloads/gdc_manifest.2020-10-12.txt", sep="\t"
)
manifest["filename"].apply(lambda v: v.split(".")[5].split("-")[3][:2]).value_counts()

# Manifest all
manifest_all = pd.read_csv(
    f"~/Projects/LiquidBiopsy/downloads/gdc_manifest.2020-10-12_all.txt", sep="\t"
)
manifest_all["filename"].apply(
    lambda v: v.split(".")[5].split("-")[3][:2]
).value_counts()
```
