## Supplementary File 1 for "DNA methylation fingerprint for the diagnosis and monitoring of hepatocellular carcinoma from tissue and liquid biopsies": TCGA_UUIDs.R

```
# Setup
setwd("~/Projects/LiquidBiopsy/")

# Imports
library(R.utils)
library(TCGAutils)
library(BiocFileCache)
library(rtracklayer)
library(MultiAssayExperiment)
library(RTCGAToolbox)
library(curatedTCGAData)

# Read manifest
manifest <- read.table(
  "downloads/Methylation_450k_TCGA_normals/gdc_manifest.2020-10-14.txt",
  header = T,
  stringsAsFactors = F,
)

# Convert
manifest_barcode <- UUIDtoBarcode(manifest[,'file_id'], legacy = T, from_type = 'file_id')

manifest_barcode_dict <- c()
for (n in names(manifest_barcode$associated_entities)) {
  if (length(unlist(manifest_barcode$associated_entities[n], use.names = F)) == 1) {
    manifest_barcode_dict[n] <- unlist(manifest_barcode$associated_entities[n], use.names = F)
  }
}
manifest_barcode_dict <- data.frame(manifest_barcode_dict)

write.csv(manifest_barcode_dict, 'downloads/Methylation_450k_TCGA_normals/TCGA_UUID_barcode.csv', quote = F)
```
