## Supplementary File 1 for "DNA methylation fingerprint for the diagnosis and monitoring of hepatocellular carcinoma from tissue and liquid biopsies": TCGANormalsFilterManifest.py

```
import pandas as pd

# TCGA UUID map
uuid = pd.read_csv(
    "downloads/Methylation_450k_TCGA_normals/TCGA_UUID_barcode.csv", index_col=0
)["manifest_barcode_dict"]

# Filter by normals
NORMALS = ["10", "11", "12", "13", "14"]
uuid = uuid[[c.split("-")[3][:-1] in NORMALS for c in uuid]]

# TCGA normals manifest
manifest = pd.read_csv(
    "downloads/Methylation_450k_TCGA_normals/gdc_manifest.2020-10-14.txt", sep="\t"
)

manifest_normals = manifest[manifest["file_id"].isin(uuid.index)]

manifest_normals.to_csv(
    "downloads/Methylation_450k_TCGA_normals/gdc_manifest.2020-10-14.normals.txt",
    index=False,
    sep="\t",
)
```
